# Regional probabilistic situational awareness and forecasting of COVID-19

**DOI:** 10.1101/2021.10.25.21265166

**Authors:** Solveig Engebretsen, Alfonso Diz-Lois Palomares, Gunnar Rø, Anja Bråthen Kristoffersen, Jonas Christoffer Lindstrøm, Kenth Engø-Monsen, Louis Yat Hin Chan, Ørjan Dale, Jørgen Eriksson Midtbø, Kristian Lindalen Stenerud, Francesco Di Ruscio, Richard White, Arnoldo Frigessi, Birgitte Freiesleben de Blasio

**Author notes:** Shared last authors (Arnoldo Frigessi, Birgitte Freiesleben de Blasio).

## Abstract

Mathematical models and statistical inference are fundamental for surveillance and control of the COVID-19 pandemic. Several aspects cause regional heterogeneity in disease spread. Individual behaviour, mobility, viral variants and transmission vary locally, temporally and with season, and interventions and vaccination are often implemented regionally. Therefore, we developed a new *regional* changepoint stochastic SEIR metapopulation model. The model is informed by real-time mobility estimates from mobile phone data, laboratory-confirmed cases, and hospitalisation incidence. To estimate locally and time-varying transmissibility, case detection probabilities, and missed imported cases, we present a new sequential Approximate Bayesian Computation method allowing inference in useful time, despite the high parametric dimension. We test our approach on Norway and find that three-week-ahead predictions are precise and well-calibrated, suitable for real-time surveillance.

**Significance:** We developed a *regional* infectious disease spread model focussing on operational usefulness in real time. The model is informed by near real-time mobile phone mobility data, laboratory-confirmed cases, and hospitalisation incidence. The model is used to estimate reproduction numbers and provide regional predictions of future hospital beds. Regional reproduction numbers are important due spatio-temporal heterogeneity due to for example local interventions. We assume different regional reproduction numbers for different periods of the epidemic. We propose a new calibration method to estimate the reproduction numbers and other parameters of the model, tailored to handle the increasingly high dimension of parameters over time. The model has been successfully used for local situational awareness and forecasting for the Norwegian health authorities during COVID-19.

## 1 Introduction

The COVID-19 pandemic has led to an unprecedented global crisis. Health authorities worldwide are continuously striving to effectively mitigate the disease spread, balancing the protection of health with social and economic costs. In most countries, the disease spread is characterised by significant geographical and temporal heterogeneity, requiring local tailoring of interventions. Therefore, time-sensitive information about the regional conditions becomes essential to management.

Mathematical modelling has been influential for preparedness planning and decision-making[1, 2, 3, 4, 5, 6]. Most of the published COVID-19 models are at national scale and when at regional level they do not account for inter-regional mobility[7, 8, 9, 10, 11].

We developed a real-time spatio-temporal SARS-CoV-2 metapopulation model used successfully in practice for assessment, monitoring, and short-term prediction, to inform regional and national policy decisions. The model exploits mobile phone mobility data and is calibrated using a new sequential Monte Carlo Approximate Bayesian Computation (SMC-ABC)[12] which we call the Split-SMC-ABC.

By including geography in our transmission model, we can provide surveillance indicators and predictions on a regional level which can take into account different interventions in different regions, a core task to guide local health authorities. Though regional estimates are valuable in local decision making, they pose a more difficult and high-dimensional estimation task. The proposed calibration method has been tailored to handle the increasing parameter dimension over time, so to provide timely estimates of key model-derived indicators, including reproduction numbers for situational awareness, and prediction of future hospital and ventilator beds. Importantly, we document predictive improvement of our regional model compared to a model with national reproduction numbers.

As an example, we implemented our framework for the spread of COVID-19 in Norway. The disease spread in Norway is characterised by continuous high circulation in Oslo and the densely populated areas surrounding the capital, and spatially shifting outbreaks in the remainder of the country. Before vaccination, the Norwegian mitigation strategy focussed on early detection, isolation, and municipality-based contact tracing, supported by national contact tracing teams, for rapid quarantining of contacts. Norway experienced its first confirmed SARS-CoV-2 cases in February 2020, followed by a rapid acceleration of cases. On 03/12/2020, the Norwegian government enforced a nationwide lockdown[13], succeeded by border closure[14] and internal travel restrictions[15]. In the following months, the measures were gradually removed. The epidemic resurged in the autumn and winter, leading to new national restrictions[16, 17, 18] and many locally targeted interventions[19, 20, 21].

Although our model is tailored to the management of the early COVID-19 epidemic, it remains applicable as a situational awareness tool during vaccination rollouts. The methodology can easily be applied to other countries and settings, and tailored to other infectious diseases where regional indicators need to be inferred as the epidemic unfolds.

## 2 Results

The estimated regional effective reproduction numbers are provided in table 1 for the early part of the Norwegian pandemic, for the rest see supplementary text section S3.7. We observe differences between regions, both in estimates and uncertainty. Estimates are higher and most certain for the largest counties Oslo and Viken, with most cases.

**Table 1:** Top: Estimated regional effective reproduction numbers (mean and 95% CI) in the early period. Middle: Estimated reduction of transmissibility at lock-down start and stop, population size and density[22]. Bottom: Estimated national effective reproduction numbers (mean and 95% CI).

| Region | R<sub>0</sub> until 03/14/2020 | R<sub>1</sub> from 03/15/2020 | R<sub>2</sub> from 04/20/2020 |  |
| --- | --- | --- | --- | --- |
| Viken | 3.92 (2.95-4.72) | 0.27 (0.09-0.47) | 0.84 (0.74-0.96) |  |
| Oslo | 5.11 (4.44-5.85) | 0.54 (0.41-0.71) | 1.04 (0.95-1.13) |  |
| Vestland | 3.28 (2.08-4.48) | 0.33 (0.05-0.55) | 0.51 (0.11-0.91) |  |
| Rogaland | 3.48 (2.28-4.43) | 0.2 (0.03-0.4) | 0.68 (0.1-1.15) |  |
| Trøndelag | 3.64 (1.85-5.53) | 0.62 (0.31-0.98) | 0.67 (0.32-0.91) |  |
| Vestfold og Telemark | 3.13 (1.53-4.84) | 0.25 (0.02-0.57) | 0.39 (0.07-0.69) |  |
| Innlandet | 3.73 (1.95-5.39) | 0.54 (0.2-0.84) | 0.49 (0.07-0.8) |  |
| Agder | 2.99 (1.68-4.04) | 0.32 (0.06-0.56) | 0.51 (0.08-0.96) |  |
| Møre og Romsdal | 3.41 (1.26-5.59) | 1.06 (0.84-1.29) | 0.42 (0.04-0.94) |  |
| Troms og Finnmark | 3.26 (1.85-4.47) | 0.25 (0.03-0.65) | 0.75 (0.17-1.27) |  |
| Nordland | 3.89 (1.44-6.24) | 0.38 (0.04-0.81) | 0.63 (0.17-1.08) |  |
| Region | Reduction *R*<sub>1</sub> | Reduction *R*<sub>2</sub> | Population | Population density (1/m<sup>2</sup>) |
| Viken | 0.93 (0.84-0.98) | 0.79 (0.67-0.84) | 1 241 165 | 55 |
| Oslo | 0.89 (0.84-0.93) | 0.8 (0.75-0.84) | 693 494 | 1 628 |
| Vestland | 0.9 (0.74-0.99) | 0.84 (0.56-0.98) | 636 531 | 20 |
| Rogaland | 0.94 (0.82-0.99) | 0.80 (0.5-0.98) | 479 892 | 56 |
| Trøndelag | 0.83 (0.47-0.94) | 0.82 (0.51-0.94) | 468 702 | 12 |
| Vestfold og Telemark | 0.92 (0.63-0.99) | 0.88 (0.55-0.99) | 419 396 | 26 |
| Innlandet | 0.86 (0.57-0.96) | 0.87 (0.59-0.99) | 371 385 | 8 |
| Agder | 0.89 (0.67-0.99) | 0.83 (0.43-0.98) | 307 231 | 21 |
| Møre og Romsdal | 0.69 ((-0.02)-0.85) | 0.88 (0.25-0.99) | 265 238 | 19 |
| Troms og Finnmark | 0.92 (0.65-0.99) | 0.77 (0.31-0.96) | 243 311 | 3 |
| Nordland | 0.9 (0.44-0.99) | 0.84 (0.25-0.97) | 241 235 | 7 |
| National reproduction numbers |  |  |  |  |
| R | Period | R | Period |  |
| 3.24 (2.47-3.98) | From 02/17/2020 | 0.49 (0.41-0.58) | From 03/15/2020 |  |
| 0.66 (0.29-1.1) | From 04/20/2020 | 0.65 (0.12-1.09) | From 05/11/2020 |  |
| 0.95 (0.14-1.63) | From 07/01/2020 | 1.09 (0.73-1.36) | From 08/01/2020 |  |
| 0.9 (0.73-1.1) | From 09/01/2020 | 1.28 (1.09-1.47) | From 10/01/2020 |  |
| 1.23 (1.04-1.5) | From 10/26/2020 | 0.81 (0.75-0.87) | From 11/05/2020 |  |
| 1.06 (1.02-1.12) | From 12/01/2020 | 0.6 (0.5-0.71) | From 01/04/2021 |  |
| 0.8 (0.65-0.93) | From 01/22/2021 | 1.5 (1.39-1.64) | From 02/08/2021 |  |
| 1.08 (1.01-1.14) | From 03/02/2021 |  |  |  |

The effect of the lockdown on reproduction numbers was significantly different between regions, with reductions ranging from 69% to 94% compared to before lockdown. After reopening schools and kindergartens, the reduction moved to between 77% and 88%. In Oslo, the reduction due to the lockdown is estimated to 89% (95% CI 84%-93%), and 80% (75%-84%) when lockdown was eased.

The fit to the daily hospitalisation and laboratory-confirmed case incidence is provided in figures 1 and 2 respectively, for some counties. The uncertainty varies and naturally is larger for counties with fewer cases (e.g., Nordland).

**Figure 1:**
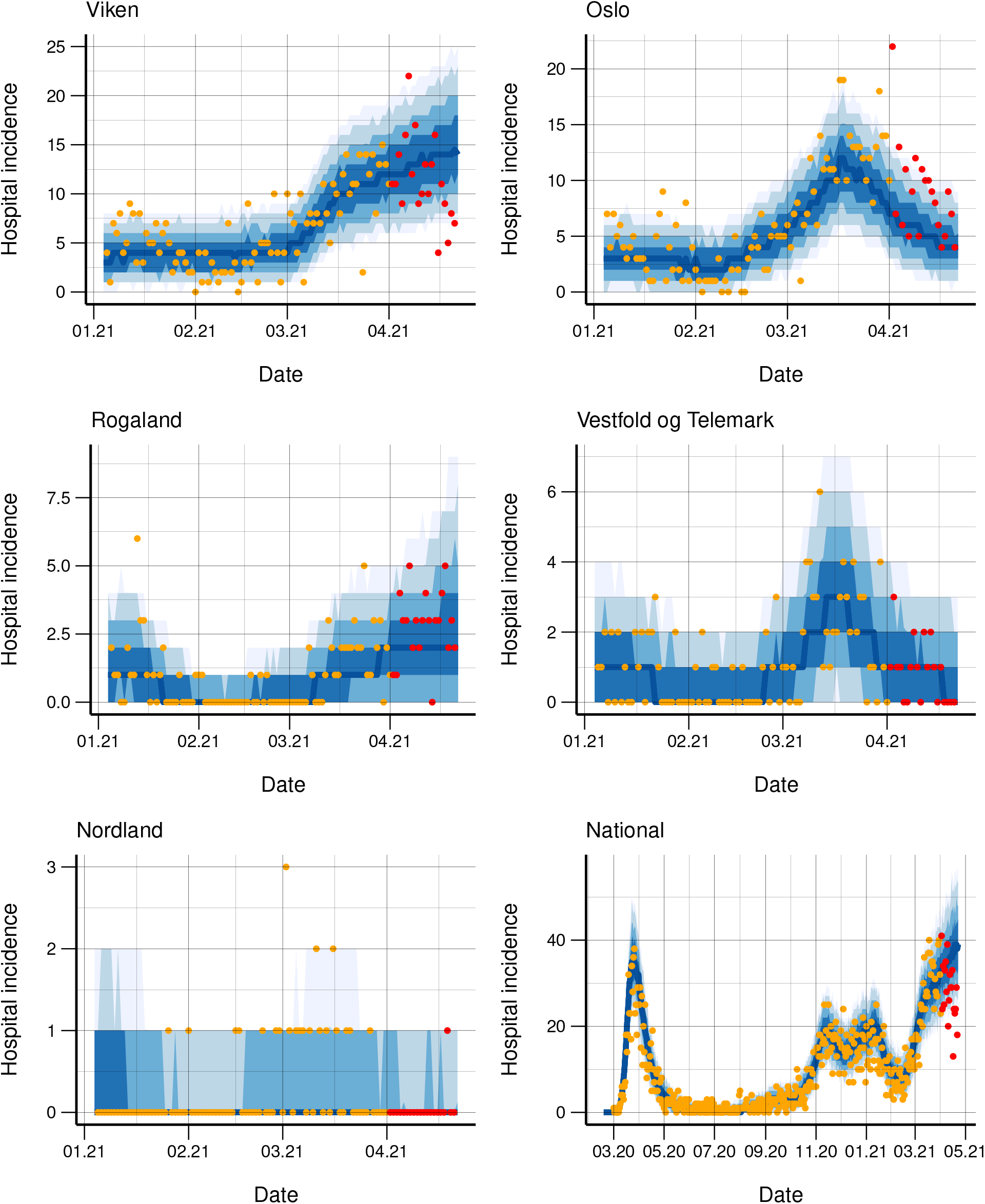
Observed daily (orange dots) and simulated (blue bands) hospitalisation incidence for five counties and nationally. 3-week-ahead predictions (blue bands) from 01/04/2021 are included together with actual data (red dots), which are not used in the calibration.

**Figure 2:**
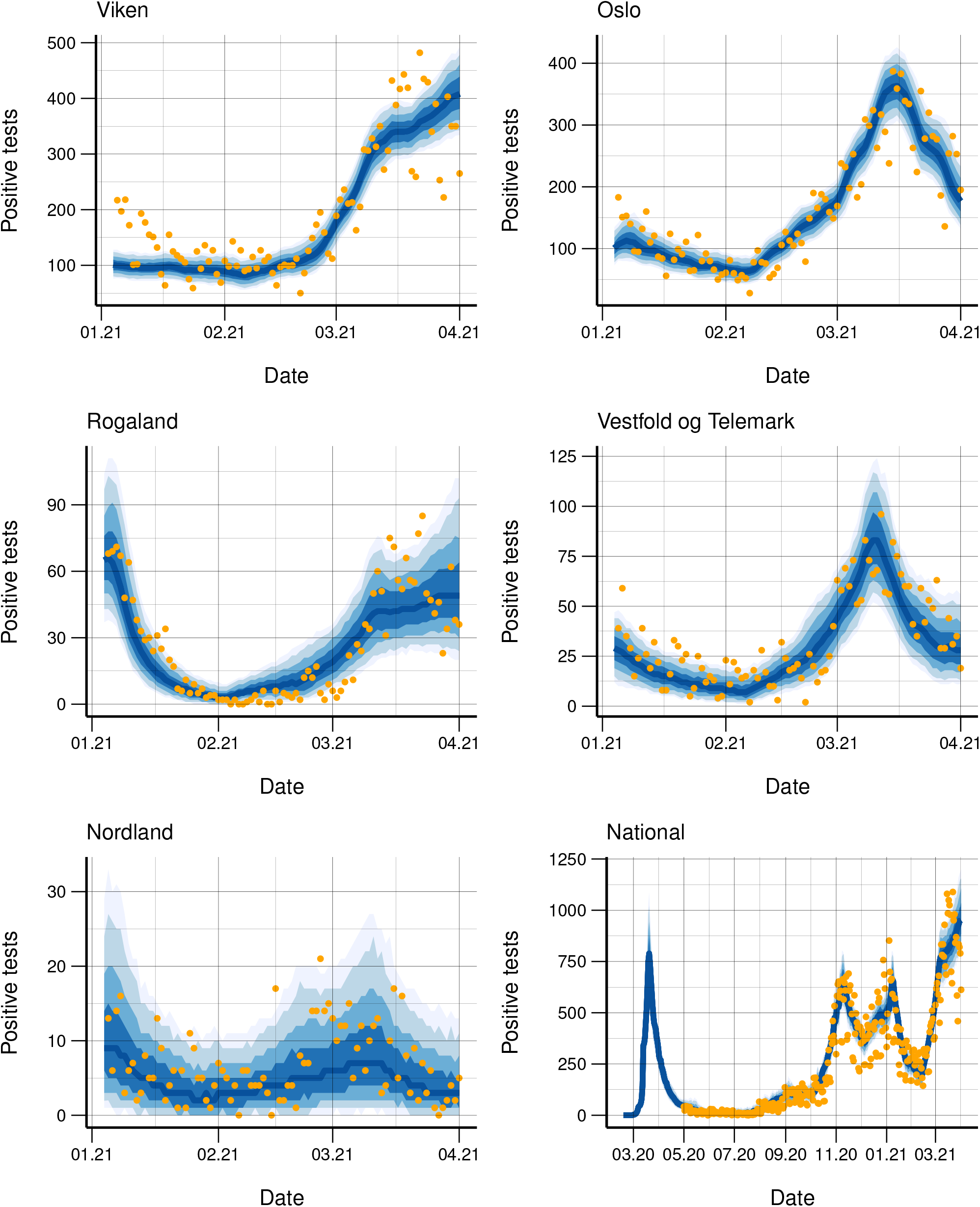
Observed daily (orange dots) and simulated (blue bands) laboratory-confirmed cases for five counties and nationally.

### 2.1 Regional predictions

Regional predictions of hospitalisations have been pivotal for preparedness and capacity planning. Importantly, our predictions assume no changes in the transmissibility in the short-term, hence showing what would happen if policies and behaviour remained unchanged. However, our predictions often triggered new policies and behavioural changes, which in turn can influence reproduction numbers. Therefore, we produce only three-weeks-ahead predictions.

In figure 1, we show three-weeks-ahead predictions of hospitalisations, using data until 04/01/2021. As additional interventions were implemented in Norway in late March 2021, our predictions overestimate actual data in county Viken and nationally. This is due to the delay between transmission and laboratory-confirmed test and/or hospitalisation: the changes were not yet visible in the data used for calibration. In the supplementary text (figures S8-S33), we present predictions for other dates, where the predictions are better because intervention policies remained unchanged.

When the main interest is predicting hospitalisations, it is unclear whether it is best to use both test and hospitalisation data or only the latter. On one side, using all available data should reduce uncertainty, and test data contain more information about recent changes because of the shorter delay. But if the two data sources are not appropriately coherent, the hospital predictions may be substantially biased. We compared hospital predictions when calibrating to both test and hospital data, to when calibrating only to the latter. Results (see supplementary text figures S12-S17) showed that point predictions were more accurate when only calibrating to the hospital data. There was a systematic underestimation of the predicted hospitalisations when also using the test data. However, prediction uncertainty is larger when using only the hospitalisation data.

### 2.2 The national picture

To present a national picture, we calibrate the model assuming the same transmissibility in all regions (table 1). In Norway, the estimated national reduction in transmissibility after the lockdown in March 2020 was 85% (78%, 89%), to a reproduction number significantly below 1. After reopening schools, we estimate a reduction of 79% (65%, 91%) compared to before interventions. During late spring 2020, many restrictions were lifted, but the estimated mean/median effective reproduction number stayed below 1 until 08/01/2020, when the borders, universities and schools reopened. During autumn 2020, the national reproduction number increased. Many restrictions were again implemented on 11/05/2020, and we estimate a reduction of 75% (69%, 80%) compared to *R*_0_. For the restrictions on 01/04/2021, we estimate a reduction of 81% (76%, 85%) compared to *R*_0_. On 03/02/2021, Oslo implemented several restrictions. Nationally, we estimate an effect of these interventions of 66% (57%, 73%) compared to *R*_0_.

### 2.3 Policy compliance and reduced mobility

The mobility data are indicators for compliance with mobility reducing policies, like travel restrictions and teleworking. Figure 3 shows the daily number of trips out from each of Norway’s counties. There was a clear reduction in mobility during the March 2020 lockdown of approximately 50%. Thereafter, mobility increased steadily until the summer. In July, geographic differences in mobility correspond to summer vacations. Later in 2020, Norway experienced high, regionally varying mobility, with peaks around vacations. Late 2020 and early 2021, mobility decreases again during the second and third waves. The systematic mobility drop on weekends indicates commuting to work even though teleworking was recommended.

**Figure 3:**
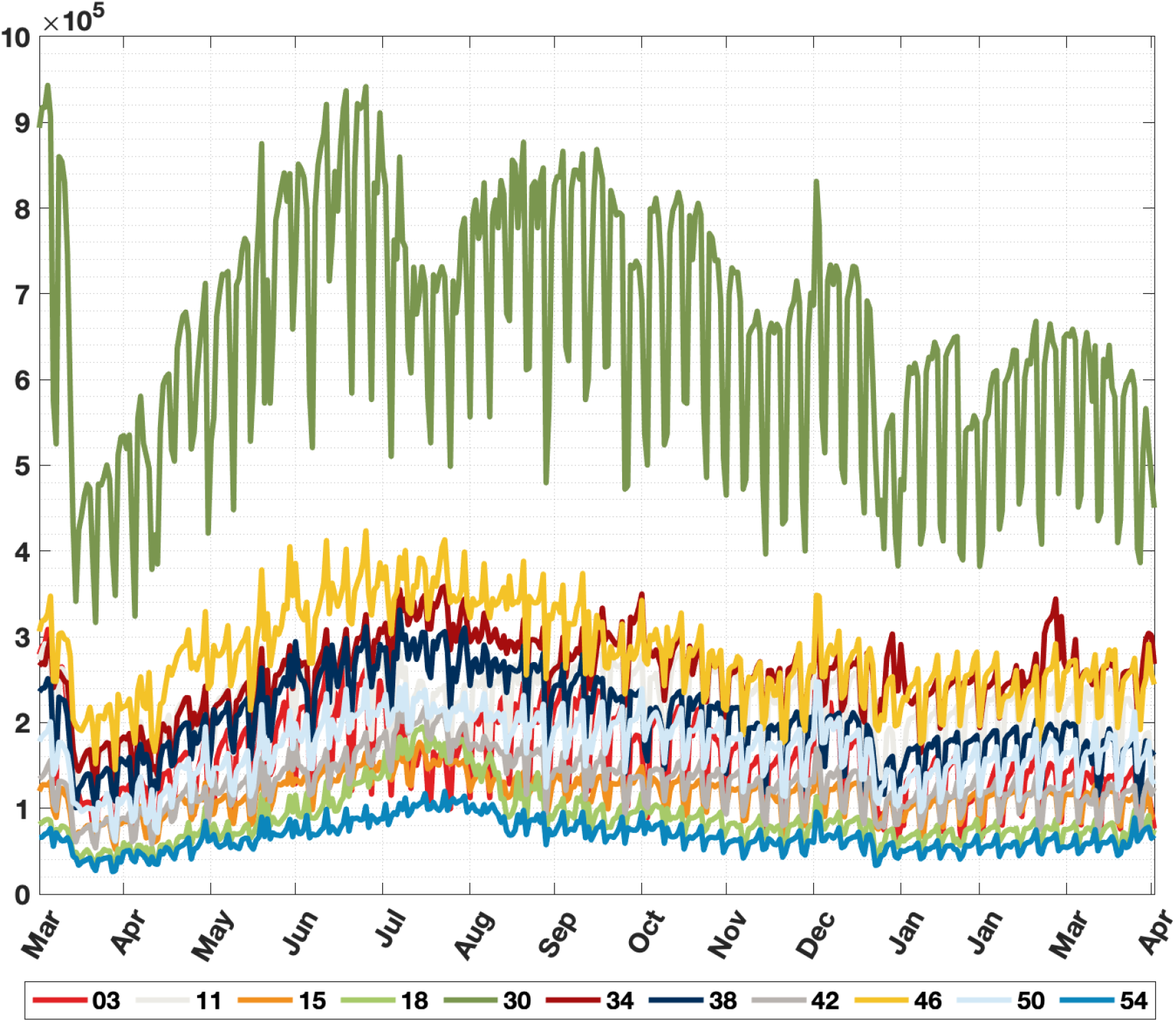
Daily time series of aggregated outgoing mobility from Norway’s counties: Oslo (03), Rogaland (11), Møre og Romsdal (15), Nordland (18), Viken (30), Innlandet (34), Vestfold og Telemark (38), Agder (42), Vestland (46), Trøndelag (50), Troms og Finmark (54).

For some counties (e.g., Troms og Finnmark), mobility has recovered to the levels prior to the first lockdown, while this is not the case for others (Oslo and Viken).

### 2.4 Half of the imported cases were discovered

In the beginning of an epidemic and whenever the disease level is low, importation of the virus into the regions is decisive. In Norway, many of the early imported cases were identified, as testing of everyone returning from known risk areas was recommended[23]. However, an unknown number of imported cases escaped control. We estimated that only 49% (33%, 84%) of the imported cases were notified and registered.

### 2.5 Predictive improvement with regional transmissibility

We investigate the predictive performance of the proposed model with regionally varying reproduction numbers for Norway and compare it to the model using a nationally constant reproduction number. We predict the weekly hospitalisation incidence and the number of new confirmed symptomatic cases region-wise for three weeks ahead following 04/01/2021, 03/01/2021, 11/01/2020, 10/01/2020, and 09/01/2020. We calibrate using data up to the prediction date. To quantify the quality of predictions and compare models, we calculate a multivariate energy-score for the region-wise predictions and a continuous ranked probability score (CRPS) for the aggregated national predictions[24], using the R-package scoringRules[25]. Proper scoring rules allow to study the quality of probabilistic forecasts by simultaneously considering the overlap between the predictions and observations and the width of the forecast distribution. Lower scores indicate better predictive performance. We also present the percentage of weeks per region when the observed hospital incidence is within the 95% prediction interval (PI).

Results, averaged over the five periods, are in figure 4, see also supplementary text table S3. The model with regional reproduction numbers clearly outperforms the national model on predicting hospital incidence and confirmed cases at the regional level, shown by the regional energy scores. The national model does slightly better on the aggregated national CRPS score for shorter predictions of 1 week, but the regional model performs better at national level for predictions two and three weeks ahead. The coverage of the regional 50% and 95% PI is good for the hospital admissions, but low for the test data. This is probably since we use a 7-days moving average in the calibration of the test data, to avoid estimating additional day-of-the-week effects. In Norway, short term predictions of hospital admissions have been an important public health tool, while predictions of the number of confirmed cases have mainly been used for model checking and validation.

**Figure 4:**
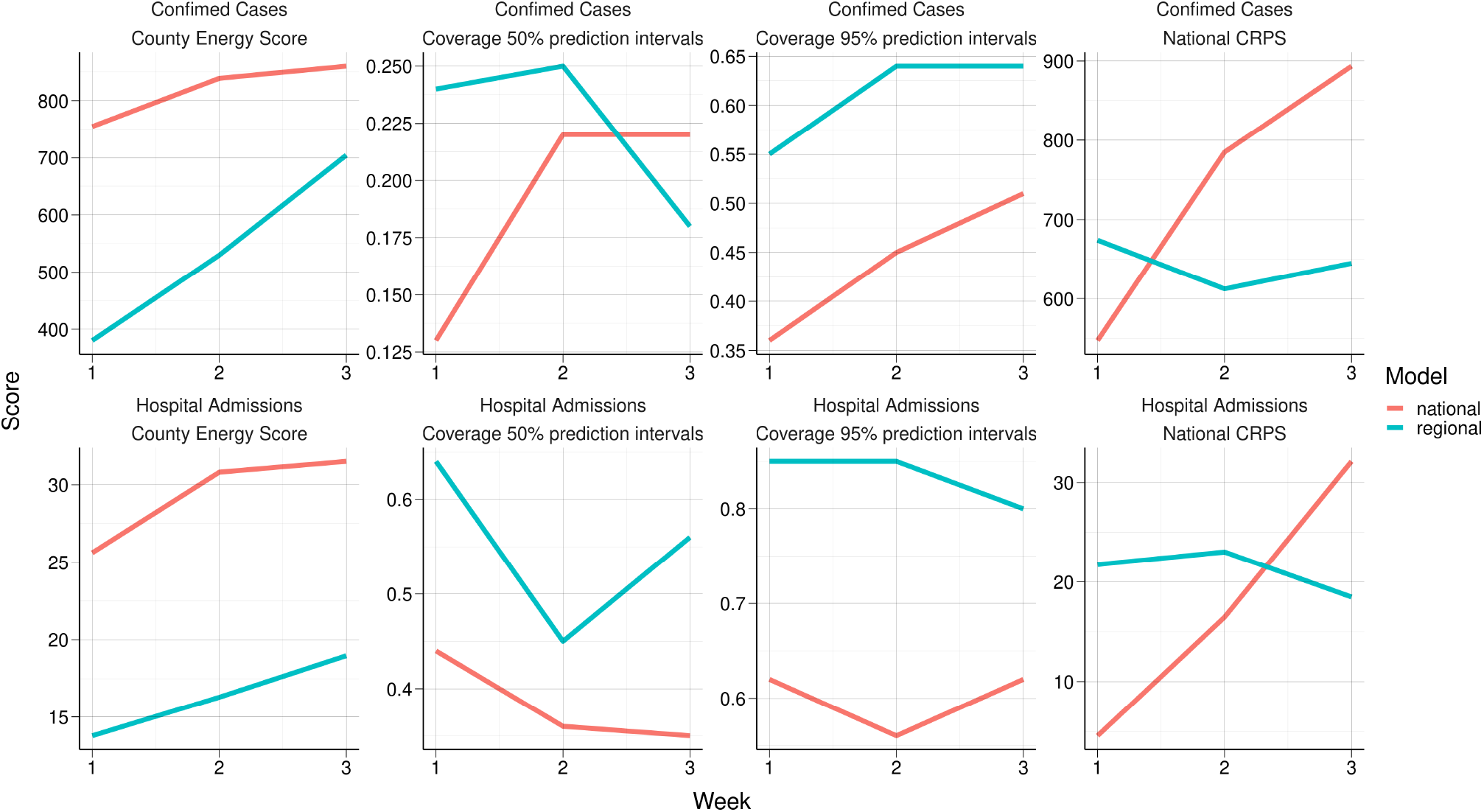
Average county energy score, national CRPS and county level coverage of 50 and 95% PI for the regional and national model.

## 3 Discussion

We presented a new calibration framework and regional model for situational awareness and surveillance of a pandemic. During a pandemic like the COVID-19, timely information about the effects of most recent interventions is crucial for operational policy decisions. We show how it is possible to use both the laboratory-confirmed cases and the hospitalisation data to inform our regional model and show the importance of the integration of both data sources. Hospital incidence data are reliable, but suffer of a time delay. The case data are less delayed and contain information on the most recent days; however, repeated changes in testing criteria, capacity, and technologies, make case data difficult to de-bias, as needed for inference. We use the total number of tests performed as a surrogate for testing capacity and suggest using case data only under stable testing conditions.

Multiple data sources are often inconsistent, because of unstable and noisy measurements. Inconsistency is known to be challenging[26] and it results in compromising parameter estimates between the data sources. While this was useful to make estimation of reproduction numbers more precise, we showed that when predicting hospital occupancy, calibrating to only hospital data resulted in less bias, but considerably higher variance.

Inference for many parameters is necessary in a regional model and has been an insurmountable challenge for useful-computational-time operations. Our new calibration method consists of series of calibrations each with less parameters, by splitting the calibration data into different periods and moving chronologically. Once the past parameters have been estimated, they are kept fixed to the posterior distribution so that one can use all the computational time in operation to estimate the most recent reproduction numbers.

A key feature of our model is the lack of age structure in the disease dynamics, which would require many more parameters. Instead, we suggest a data-driven approach, using the age distribution of the positive test cases to describe the age-profile of transmission. In this way we can account for behavioural changes in the population, avoiding doubtful a priori assumptions about mixing patterns between age groups.

Similarly, our approach copes with vaccination in an innovative way. We have not included additional compartments for vaccinated individuals in the metapopulation model. That would have required multiple assumptions about the timevarying effects of the different vaccines. Instead, we estimate the effective reproduction number after vaccination directly from the data. Hence, the estimated effective reproduction number automatically captures the population which is immune due to vaccines. In Norway, vaccination has followed age groups. Since we use the age profile of the test data to calculate hospitalisation risks, the effect of the vaccination is visible as a decay in hospitalisation risks for the vaccinated age groups and in this way enter model. By 04/01/2021, approximately 5.4% and 7.5% of the Norwegian population had received their second and first vaccination dose, respectively.

We tested our model in the case of Norway, where many essential aspects are present: incompletely observed importation; time-varying testing capacity and test-targeting strategies; focus on regional capacity limits of the health system; nationwide and local interventions; uncertain changes in population behaviour and adherence; spatial heterogeneity of the epidemic with clusters in the largest cities. Nationally, we estimated a reduction of 85% in transmissibility due to the first lockdown, similar to the UK (estimated reduction 75%[27]), Germany (79%[28]) and all Europe (81% (range 75% to 87%)[29]). We estimate larger reduction effects in the larger regions which also had the most cases, compared to the reduction in the national average. Similar results were found for London (81%[27]) compared to the whole UK.

Though we found that the early interventions successfully contained the epidemic, we are not able to discern how the various measures contributed. Interestingly, we found that the reopening of kindergartens and schools did not increase transmissibility significantly. Similar findings have been reported in China, where the increase in intra-city movement after the lockdown was negatively correlated with transmissibility[30]. We also found that the transmission rate did not grow significantly when mobility was almost back to normal. Hence, mobility appeared not to drive infectious contacts, possibly because of hygienic measures and maintaining distances.

The regional model performed better at predicting hospitalisations than the model assuming the same transmissibility nationally. Estimating regional reproduction numbers is possible and improves local hospital planning.

We did not estimate changepoints from data but chose them manually. The model performance depends on their placements. Failure to add a changepoint when there is a significant change in transmission, makes the model perform badly because more parameters are needed to cope with non-stationarity. Methods for data-driven changepoint detection[28] can be very useful.

For Norway, we used a single nationwide detection probability for the laboratory-confirmed cases, reflecting the national testing criteria. Testing practice and thus detection probabilities may differ between regions. It is possible to estimate separate detection probabilities per region, but with a significant computational cost. In addition, testing criteria have varied in time, independently of the number of tests performed, affecting the consistency between test and hospital data.

The three-weeks-ahead predictions could be improved by incorporating estimated effects of planned interventions during this period. For this purpose, data on the effect of different restrictions (both isolated and in combination) on regional transmissibility are necessary. Such data are not yet available and is a theme for further research.

## 4 Material and methods

### 4.1 Case data

We utilise different COVID-19 data sources. We have access to anonymised individual-level hospital incidence data from the Norwegian Intensive Care and Pandemic Registry. This data set includes all patients admitted within 14 days of a positive laboratory test or diagnosed with COVID-19 in the discharge report for all hospitalised individuals in Norway until 04/01/2021. From MSIS, we also have access to all the positive and negative COVID-19 tests performed from 04/01/2020 until 04/01/2021 in Norway, and for each tested case we have information about whether or not the individual was infected abroad. Additional details are provided in the supplementary text section S2.1.

### 4.2 Mobility data

We estimate mobility between counties of Norway by mobile phone data for the whole epidemic period until 04/01/2021. Every day is divided into six-hour intervals, during which the total number of individuals transitioning between counties is counted. Individual movements are not tracked over time. When predicting the future, we regularise the most recent movement matrix to ensure conservation of the number of people per county and the rare depletion of inhabitants in smaller counties. We use all subscribers to Telenor Norway, the largest mobile network operator. Additional details are given in supplementary text section S2.2.

### 4.3 Infectious disease spread model

For situational awareness and forecasting of the COVID-19 epidemic, we have developed a stochastic spatial metapopulation SEIR model[31, 32]. The model consists of three layers: (i) the total population, distributed in regions, with a region-specific age profile; (ii) a mathematical transmission model per region; (iii) an inter-regional mobility model informed by mobile phone data.

Multiple studies indicate the importance of presymptomatic transmission[5, 33, 34] and asymptomatic infection[35] of SARS-CoV-2. We include six compartments in the model: susceptible, exposed, presymptomatic, infectious with symptoms, infectious asymptomatic, and recovered. Details are provided in supplementary text section S2.4. For each region, we assume that transmissibility is a step-function, constant between changepoints, corresponding to changes in regional regulations. We calculate the expression for the basic reproduction number of this model, *R*_0_, as the largest eigenvalue of the next generation matrix, see supplementary text section S3.8. The effective reproduction number is obtained by multiplying the eigenvalue with the average proportion susceptible during the constant periods between changepoints.

#### 4.3.1 Changepoint specification

For Norway, we estimate the first changepoint nationwide between 03/12/2020-03/16/2020 which is when the first national lockdown was implemented. The best fit was for 03/15/2020 (see supplementary text section S3.8). The second changepoint was set on 04/20/2020, when the kindergartens reopened nationwide. After that, the regional changepoints were set when there were changes in regional policies. In addition, we include further changepoints to capture other changes in behaviour or viral properties. The results presented for Norway are calibrated to data up to 04/01/2021, though the model in this paper, has been in constant use for the Norwegian health authorities at least until 10/01/2021[36].

#### 4.3.2 Importation

We seed the epidemic with infected cases imported from abroad, in their residence county. As we expect that some imported cases go undetected, we import an additional random, Poisson distributed number of cases for each observed imported case, with mean estimated from the data during calibration. For Norway in this paper we calibrate this amplification factor between 02/17/2020 and 04/01/2021.

### 4.4 Calibration

To estimate the model parameters, we calibrate to region-specific daily hospital incidence and laboratory-confirmed cases. Using multiple data sources in inference is useful, as each component might carry unique information, intrinsically or due to measurement error[26, 37]. For these reasons however, inconsistency between data sources can appear, challenging inference.

We use age-specific hospitalisation risks, the age structure of each region, and the age distribution of the laboratory-confirmed cases, to simulate hospitalisations. The time between symptom onset and hospitalisation is estimated from registry data on all hospitalised cases. This leads to a stochastic time shift between transmission and hospitalisation of approximately 14 days, see supplementary text section S2.5.4 for details.

We model the laboratory-confirmed cases as a binomial process of the daily incidence. The case detection probability by testing depends on the total number of tests, as a proxy for the time-varying effort to detect cases. As our model only estimates transmission within a country, we exclude the cases imported from abroad. For Norway, we choose not to use the laboratory-confirmed cases before 05/01/2020, as the test criteria and capacity were significantly different early in the epidemic.

Inference was based on a new version of SMC-ABC (split-SMC-ABC, see supplementary text section S2.8), resulting in samples from the approximate joint posterior distribution of the parameters. ABC is known to fail to scale in the number of parameters[38]. For Norway, after a year, there are almost 100 parameters, far beyond what is feasible for SMC-ABC, which then fails to converge in useful time. To our knowledge, our new split-SMC-ABC is the first method able to estimate so many parameters in a stochastic model, in time useful for operational regional surveillance, exploiting that correlations of transmissibility become less important over time. It has turned out to be an important instrument in decision making in Norway[36]. To simulate the future, we propagate posterior samples into the future, and obtain credibility intervals (CI) of the forecasts.

The split-SMC-ABC is based on temporal splits of the data and calibration. We first perform a calibration of the early part of the epidemic using data until a predefined split point. The results are then probabilistically propagated further, after the split, such that in operation we only calibrate the post-split parameters, to the most recent data. Multiple splits can be introduced. Data overlap between splits, to reduce discontinuities in inference and to cope with time delays between data and transmission.

Often, regions are demographically and epidemiologically different. When all the regional parameters are calibrated simultaneously, the more populated regions with more cases are typically estimated better. In the last post-split calibration, we therefore first perform a separate calibration for each region, assuming a common transmissibility for all other regions. The results for each region are then used to inform prior distributions in a final simultaneous calibration of all the parameters. This final simultaneous calibration is necessary to handle the spatial dependence.

## Data Availability

Non-sensitive Norwegian COVID-19 case data are available at: https://github.com/folkehelseinstituttet/surveillance_data. Individual requests for the regional COVID-19 sentive case data should be made to the Norwegian Institute of Public Health, Registry Department. Requests for access to mobility data should be directed to Telenor Research on. The infectious disease spread model is publicly available on GitHub in the R-package spread, and can be found on https://github.com/folkehelseinstituttet/spread.

## Acknowledgements

This project was funded by the Norwegian Research Council project number 312721. SE and AF acknowledge partial funding from the Norwegian Research Council centre BigInsight project 237718. AF acknowledges support from Nordic-MathCovid funded by Nordforsk. The authors are grateful to Geir Storvik, Magne Aldrin, Gry Marysol Grøneng and Pia Karoline Abel-Zur Wiesch Genannt Hülshoff for discussions on models during the COVID-19 pandemic in Norway. We are grateful to Norwegian Institute of Public Health, Beredskapsregisteret for covid-19 for gathering the Norwegian COVID-19 data, making it available for use.

