## Supplementary Information (SI) for "Regional probabilistic situational awareness and forecasting of COVID-19"

### Supporting information appendix for: Regional probabilistic situational awareness and forecasting of COVID-19

October 18, 2021

### S1 Supplementary backgroud

#### S1.1 COVID-19 epidemic in Norway

In December 2019, a cluster of pneumonia cases with unknown origin, later identified as caused by the SARS-CoV-2 virus, was observed in Wuhan, China. By the end of January 2020, the virus had reached all provinces of mainland China[1]. Northern Italy emerged as the first country outside China experiencing a major outbreak of COVID-19, with 1689 confirmed cases as of March 2 2020[2]. Most European countries rapidly followed.

In late February 2020, Norway experienced its first confirmed COVID-19 case. The early Norwegian strategy aimed to prevent local spread through early detection and isolation of cases, contact tracing, and quarantining of people returning from travel to high-risk areas[3]. Frequent hand-washing and self-isolation when showing symptoms of respiratory infection were encouraged. Norway experienced many imported cases, primarily citizens returning from winter holidays in Italy and Austria.

By March 12 2020, there were more than 500 confirmed cases[4], both importations and local transmission events. On that date, the Norwegian government enforced a nationwide lockdown, mandating the closure of all kindergartens, schools, and educational institutions, imposing of teleworking for nonessential workplaces, closure of public places, cancellation of public events and businesses with a high risk of transmission[5]. Later that week, borders were closed to foreign nationals without a residence permit in Norway[6], and local travel to holiday property was prohibited[7].

The extensive regulations led to a reduction in the number of reported cases and hospitalisations in April 2020. From April 20 2020, many of the interventions were scaled back; kindergartens and primary schools reopened, followed by other schools two weeks later. In May 2020, many closed businesses could resume, and public gatherings with up to 50 people were allowed. The situation remained stable and under control during the whole summer in Norway. After summer, there was an increase in cases, in particular in the capital Oslo. On November 5 2020, national contact reducing interventions were implemented[8]: people were recommended to stay at home as much as possible, keeping two meters distance to anyone in risk groups, avoid unnecessary domestic travel, and alcohol serving after midnight was banned. On January 4 2021, further contact reducing interventions were implemented. Norwegians were asked not to invite others home. All indoor sports and cultural events were discouraged. All teaching at universities had to be digital, and serving of alcohol was banned[9]. Further national restrictions followed on March 25 2021[10]. Over the course of the pandemic, regionally varying transmissibility led to numerous local restrictions in Norway, for example in Oslo[11, 12] and Bergen[13]. On March 2 2021, Oslo implemented many local restrictions. All restaurants and nonessential shops were closed. Outdoor events were banned. Upper secondary schools were closed. Additional restrictions were implemented on March 17 2021, including that it was no longer legal to have more than two guests in the house. Lower secondary schools were closed and partial digital teaching was introduced in primary schools[11].

An overview of counties in Norway is provided in figure S1.

### S2 Materials and Methods

#### S2.1 Case data

This study utilises different COVID-19 data sources. Beredskapsregisteret for COVID-19 (Beredt C19) gathered all the Norwegian COVID-19 data and made it available for use. We obtained anonymised individual-level hospital incidence data from the Norwegian Intensive Care and Pandemic Registry (NIPaR). This data set includes all patients admitted within 14 days of a positive laboratory test or diagnosed with COVID-19 in the discharge report. The data set contains age,

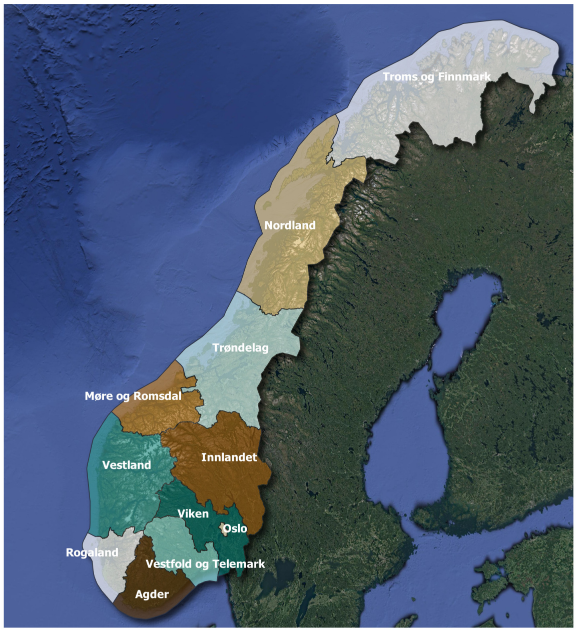


Figure S1: Overview of Norway's 11 counties.

residence region, date of hospitalisation, date of entering mechanical respirator, and discharge dates from both mechanical respirator and hospital. NIPaR was merged with data from the Norwegian Surveillance System for Communicable Diseases (MSIS), which comprises data on all notifiable infectious diseases in Norway, including COVID-19. The MSIS data include age, whether the patient was infected abroad or in Norway, date of positive test, and date of symptom onset for all COVID-19 cases. We used the data both to calibrate the transmissibility in the model and to estimate the hospitalisation parameters, see Section S2.5.3. By April 12 2021, NIPaR included 4.248 hospitalised individuals, where 735 needed mechanical respirator treatment, while MSIS contained 103.461 positive test results.

We obtained data from MSIS on the number of negative SARS-CoV-2 tests by region from April 1 2020, when the reporting commenced. In this data set, negative tests within seven days of a previous negative test were excluded. By April 12 2021, the data contained 3.426.045 negative test results. We used the negative test data to estimate the probability of detecting a case.

#### S2.2 Real-time mobile phone mobility data

The geographical spread of COVID-19 is governed by movements of infectious individuals. We have no precise data on mobility of individuals between municipalities in real-time. We used mobile phone mobility data from Telenor Norway as a surrogate.

Every mobile phone is continuously connected to a cell tower and switches from one tower to another as it moves between their areas of connectivity. The geographic location of the connected cell tower provides an approximate location of the phone, and therefore of its owner. The cell tower network also connects to every device and handset connected to the mobile operator's network. To focus on people movements, we have filtered out devices (IoT/M2M) that are not likely to be carried by an individual. For every six-hour time window, each individual subscriber of the mobile operator (in our case Telenor Norway), is assigned to the municipality most recently visited in the last two hours of the six-hour time window. An individual has moved from municipality $i$ to municipality $j$ if they were in $i$ during the last 2 hours of one six-hours period and in $j$ in the final two hours of the next six-hour period. The population travel patterns are then approximated by counting how many subscribers transitioned between any pairs of the 356 municipalities in Norway, during two consecutive six-hour periods. This represents a mobility/origin-destination matrix. Each mobility matrix contains aggregated data of anonymous counts of people travelling. There are no individual identifiers in the data. The counts in the generated mobility matrices are further up-scaled by a factor representing the overall Telenor Norway market share, estimated at 47.5% in 2019[14]. As a final privacy-preserving precaution, we censored counts below 20 in the mobility matrices to prevent potential re-identification of individuals or small groups of people. The structure of the mobility data for Norway is the following:

```
Date,    Time, From,    To, Count
20200224, 12, 1101, 1101,  8960
20200224, 12, 1101, 1103,   239
20200224, 12, 1101, 1108,   194
20200224, 12, 1101, 1111,    91
```

Specifically, the date format is YYYYMMDD; the time window is one of (00,06,12,18) designating the terminating hour in the arrival municipality; the To/From code identifies each municipality using the official Norwegian four-digits numbering system.

The time series of mobility matrices covers the period from the first confirmed case in Norway up to today and is updated daily. Hence, the model is informed by real-time mobility patterns and has been so since the first confirmed case in Norway, and we utilise actual mobility data when simulating the past. In this way, alterations in the mobility patterns as a response to interventions, seasonality, or other causes, are automatically incorporated in our model predictions.

The model uses a regularised version of the mobility matrix of the most recent non-holiday weekday when predicting future events. First, the four mobility matrices for the given weekday are averaged. Then an optimisation is performed to obtain the closest matrix that fulfils the population conservation constraint (see Section S2.3). For the predictions in this paper, we used regularised mobility data from the following dates:

| Prediction date | Regularisation date |
|---|---|
| 09/01/2020 | 09/01/2020 |
| 10/01/2020 | 10/01/2020 |
| 11/01/2020 | 10/30/2020 |
| 03/01/2021 | 03/01/2021 |
| 04/01/2021 | 03/25/2021 |

While the mobile phone mobility data is measured on the municipality level, we aggregate the data to the county level in the simulations.

We assume that the mobile phone mobility data are representative of the whole Norwegian population. In reality, there are likely sources of bias in the mobility data, for example, age bias (very young and old individuals less often possess a mobile phone and are thus not well captured). However, Telenor Norway has a substantial market share in Norway and is well distributed over all age groups.

The SEIR model operates with six-hour intervals, during which mobility between municipalities is frozen so that numerical solutions of the differential equations can be computed. Of course, our construction misses movements that happen during the four non-monitored hours. We have chosen to use six hours as the temporal resolution, in order to capture commuting and not miss long-distance trips. With a shorter time interval, we would better capture short-time/short-distance movements, compared to a longer time interval. Vice versa, with a longer time window, we would measure long distance/long time travel better. We believe that the short-duration movements are less critical in the spread of the epidemic. Moreover, the finer the time resolution, the more noise and randomness, as there are fewer individuals moving. Hence, the data become more sensitive to the exact location and density of the cell towers. In addition, a more refined time scale is also computationally more expensive. The reason why we do not monitor all locations visited by each mobile phone, and instead only record the last visited location during the last two of the six hours of each period, is computational: the full history of each visited cell tower is extremely large and it is computationally hard to process such complete data to generate matrices which are more frequent. However, this should not be critical to the workings of the model, since during the night movements are limited, and trips of short duration are less important in the spread of the epidemic.

#### S2.3 Population conservation

When we use the calibrated model for prediction, we need to consider inconsistencies in the total in- and outgoing flow for each region. Reasons for these inconsistencies are multiple: Customers turn off/on their mobile phone, issues with network coverage rendering the handset losing connection with the network, customers churning between different mobile operator's networks, etc. However, if there is an imbalance in the number of people entering and leaving a region, we will for long-term predictions risk that the population size in some regions grow out of control. In contrast, other regions may, in the worst case, end up empty. Therefore, to preserve population at the region level, we estimate a regularised mobility matrix, based on mobility data from the most recent weekday. The regularised mobility matrix is the closest to the observed mobility matrix, measured by mean squared error per entry, which satisfies the conservation constraint that total incoming flow equals total outgoing flow for each region. The obtained matrix entries are then rounded to the nearest integer.

Specifically, let $X_{ij}^t$ be the element of the mobility matrix denoting the number of people moving from location $i$ to location $j$ between time $t$ and time $t+1$. The conservation of people for each region requires the following equality:

$$X_{ii}^t + \sum_{j \neq i} X_{ji}^t = X_{ii}^{t+1} + \sum_{j \neq i} X_{ij}^{t+1}. \tag{S1}$$

Conserving the number of people in a location basically requires a balance in people not moving, people travelling out from the site and people travelling to it. In terms of the mobility matrices, this means the following: For a given location $i$ and time $t$, all incoming transitions are summed up (column-wise), and this must equal the sum of outgoing transitions at time $t+1$ (row-wise).

Assume now that we want to simulate ahead in time, using only one constant mobility matrix for all times $t$. When we use the same matrix all the time, then equation (S1) can be rewritten as (where we drop $t$); $\forall i$

$$\sum_{k:k\neq i} X_{i,k} = \sum_{k:k\neq i} X_{k,i}. \tag{S2}$$

This means that the matrix row-sums are equal to the column-sums. A symmetric matrix trivially fulfils this requirement. However, this is not the only possibility, and skewed mobility matrices are important due to commuting patterns. We want to find the closest matrix that satisfies (S2), and do this by solving the following Quadratic Programming problem:

$$\text{argmin}_Z \sum_{i,j:i\neq j} (X_{ij} - Z_{ij})^2, \tag{S3}$$

subject to the linear constraints

$$\forall i \sum_{k:k\neq i} Z_{ik} = \sum_{k:k\neq i} Z_{ki}. \tag{S4}$$

#### S2.4 Regional transmission model

A regional metapopulation model is used to simulate the spread of COVID-19 in both space and time. The model is an extension of the SEIR metapopulation model[15]. The model consists of three layers: (i) the population structure in each region, based on census data (in the case of Norway from Statistics Norway[16]), (ii) a dynamic network of movements describing the travel patterns between regions, and (iii) a local transmission model within each region.

The population is initialised according to data on population sizes in the regions (for Norway in January 2020[16]). Each individual in the model then has a designated home location. To model the disease spread, the individuals first mix in their current location for six hours. Then, the individuals travel according to the related mobility matrices, and then mix again in their new location for six hours. When we implement the travel from location $i$ to $j$, we preferentially send back the individuals with home location $j$ who are visitors in location $i$. If more people travel from $i$ to $j$ according to the mobility matrices than those from $j$ currently in $i$, we send individuals who belong to $i$ and are currently present in location $i$. If this is still not enough, we send other visitors in $i$, who belong to other regions, to $j$. In this way, the virus can spread from one region to another, for example by an individual travelling from a susceptible location to an infected location, acquiring the infection, and then travelling back. We preferentially send individuals back to their home location, keeping track of where they come from, in order not to overestimate the spread[17]. In Norway, local mobility and commuting are of primary importance. The local rules which we here adopted can be changed in other situations and countries.

###### S2.4.1 Local stochastic epidemiological model

For each time interval, we assume homogeneous mixing within each region. We assume that the individuals can be in one out of six states: susceptible ($S$), exposed and not infectious ($E_1$), presymptomatic and infectious ($E_2$), infectious symptomatic ($I$), infectious asymptomatic ($I_a$), or recovered ($R$). The additional compartments have been included to better fit the epidemiology of COVID-19 transmission. Multiple studies indicate the importance of presymptomatic transmission[18, 19, 20] and asymptomatic infection[21]. We implement the transitions between the states for individuals currently in location $i$ by stochastic difference equations:

$$S^i(t + \delta t) = S^i(t) - X_1(t), \tag{S5}$$

$$E_1^i(t + \delta t) = E_1^i(t) + X_1(t) - X_2(t), \tag{S6}$$

$$E_2^i(t + \delta t) = E_2^i(t) + X_3(t) - X_4(t), \tag{S7}$$

$$I^i(t + \delta t) = I^i(t) + X_4(t) - X_5(t), \tag{S8}$$

$$I_a^i(t + \delta t) = I_a^i(t) + X_2(t) - X_3(t) - X_6(t). \tag{S9}$$

The stochastic transitions between the states are

$$X_1(t) \sim \text{Binom}(S^i(t), \beta_t^i \delta t / N_t^i \cdot (I^i(t) + r_{I_a} I_a^i(t) + r_{E_2} E_2^i(t))),$$

$$X_2(t) \sim \text{Binom}(E_1^i(t), \lambda_1 \delta t),$$

$$X_3(t) \sim \text{Binom}(X_2(t), (1 - p_a)\delta t),$$

$$X_4(t) \sim \text{Binom}(E_2^i(t), \lambda_2 \delta t),$$

$$X_5(t) \sim \text{Binom}(I^i(t), \gamma \delta t),$$

$$X_6(t) \sim \text{Binom}(I_a^i(t), \gamma \delta t).$$

Here, $N_t^i$ is the number of individuals present at time $t$ in the region $i$, $\beta_t^i$ is the probability of transmission upon a contact times the contact rate, $r_{I_a}$ is the relative infectiousness of the asymptomatic, $r_{E_2}$ is the relative infectiousness of the presymptomatic, $1/\lambda_1$ is the latent period, $p_a$ is the probability of being an asymptomatic carrier, $1/\lambda_2$ is the presymptomatic period, and $1/\gamma$ is the infectious period and asymptomatic infectious period, assumed equal. We have one set of equations for each region $i$. The per-compartment counts $S^i$, $E_1^i$, $E_2^i$, $I^i$, $I_a^i$, and $R^i$ are the number of individuals in each compartment who are currently present in the region. For the transmission parameter $\beta_t^i$, we have a subscript denoting time-dependence, as we assume it to be a step-function. It also has a superscript for region, as we allow different transmissibility for each region.

We do not provide the equation for $R^i(t)$, the number recovered, as we assume constant population sizes, so that $N_t^i = S^i(t)+E_1^i(t)+E_2^i(t)+I^i(t)+I_a^i(t)+R^i(t)$ for each region.

By calculating the largest eigenvalue for the next generation matrix of the corresponding deterministic system of differential equations, we get an equation for the basic reproductive number $R_0$[22] of our epidemiological model, given by

$$R_0 = \beta_0 \cdot \left((1-p_a)/\gamma + p_a r_{I_a}/\gamma + (1-p_a) r_{E_2}/\lambda_2\right). \qquad \text{(S10)}$$

We calculate the effective reproduction numbers by multiplying the estimated reproduction number (given by the estimated $\beta_t^i$ instead of $\beta_0$ in equation S10) by the estimated mean proportion susceptible in the corresponding period. We chose to put our prior distributions on the reproduction numbers instead of the $\beta_t^i$, as we have more intuition about their size. Then we reverse equation S10 to calculate the $\beta_t^i$ from the reproduction numbers.

#### S2.5 Models for admission to hospital and intensive care

Based on the estimated incidence in each region obtained from the spatial metapopulation model, we model the number of individuals admitted to the hospital. In the model assuming a common, national transmissibility, we use a binomial distribution with hospitalisation probabilities provided in table S1. In the model with region-specific transmissibilities, we instead use a 28-days moving average of the daily estimated hospitalisation risk, as explained in section S2.5.2. We use a moving average because the daily hospital admission data are prone to randomness and noise. This means that in the model with a national transmissibility, we assume hospitalisation risks constant every calendar month, while when we model transmissibility regionally we assume smoothed daily hospitalisation risks. It would be straightforward to also include daily moving averages in the national hospitalisation model. When simulating, once an individual is selected to be sent to the hospital, we generate the delay from onset of symptoms to hospitalisation and the length of stay in hospital from two negative binomial distributions. For Norway, we estimated these parameters from individual-level registry data, as explained in the following sections.

##### S2.5.1 Hospitalisation risk

To calculate the hospitalisation probability conditional on infection per age group, we start with the estimated hospitalisation probabilities in[23]. Remember that our compartmental model does not have age compartments, because these would make the model computationally intractable. On the other hand, the probability of requiring hospitalisation is highly dependent on age. Given the demographic age profile of a region, we can compute the regional probability of hospital admission as a mixture of the probabilities per age group in[23], each multiplied with the proportion of the age group in the region. Compared to the French population under study in [23], Norway has a practice of not admitting elderly in nursing homes with COVID-19 to hospitals. Therefore, we adjusted the hospitalisation risk for the elderly by the percentage of nursing home residents. Finally, we corrected the hospital admission probabilities to account for age. The disease burden does not just follow the age demography, and we need to quantify the proportion of infected individuals per age group. To this end, we used the age distribution of the positive cases as a proxy for the age distribution of COVID-19 infections. Test data is a commonly available data source. For Norway, we obtained test data from the MSIS registry; see details below. To account for a changing age profile of infected throughout the pandemic, we used monthly estimates from May 1 2020 in the national model, reported in table S1. In the regional model we use daily estimates. In this paper, we predict hospitalisations using the latest test data before the prediction period to mimic data availability in a real-life situation.

##### S2.5.2 Regional hospitalisation risks

The age profile in the positive test data varies between regions and changes over time. We estimated the daily hospitalisation proportion of positive tests in each age, regionally and nationally, over a moving window of the 28 previous days. We use the observed proportion of hospitalised cases in each age group, but we add a small number in both the number of hospitalisations and number of positive tests in each age group in order to avoid dividing by zero. Hence, a pseudo count of 0.001 was added to the number of individuals hospitalised in each age group

Table S1: Hospitalisation probabilities when infected, adjusted for age, as estimated from test data, national values.

| | Until 2020-05-01 | Until 2020-06-01 | Until 2020-07-01 | Until 2021-08-01 |
|---|---|---|---|---|
| 0-9 years | 0.0001 | 0.0005 | 0.001 | 0.00045 |
| 10 - 19 years | 0.0004 | 0.001 | 0.001 | 0.001 |
| 20 - 29 years | 0.006 | 0.007 | 0.0099 | 0.009 |
| 30 - 39 years | 0.013 | 0.019 | 0.0091 | 0.017 |
| 40 - 49 years | 0.018 | 0.015 | 0.015 | 0.016 |
| 50 - 59 years | 0.043 | 0.032 | 0.016 | 0.032 |
| 60 - 69 years | 0.059 | 0.031 | 0.025 | 0.039 |
| 70 - 79 years | 0.086 | 0.048 | 0.038 | 0.032 |
| 80+ years | 0.319 | 0.134 | 0.134 | 0.029 |
| | Until 2020-09-01 | Until 2020-10-01 | Until 2020-11-01 | Until 2020-12-01 |
| 0-9 years | 0.0004 | 0.0004 | 0.0003 | 0.0006 |
| 10 - 19 years | 0.001 | 0.001 | 0.001 | 0.0015 |
| 20 - 29 years | 0.014 | 0.011 | 0.01 | 0.0072 |
| 30 - 39 years | 0.013 | 0.015 | 0.016 | 0.014 |
| 40 - 49 years | 0.013 | 0.014 | 0.017 | 0.016 |
| 50 - 59 years | 0.025 | 0.024 | 0.027 | 0.028 |
| 60 - 69 years | 0.015 | 0.030 | 0.031 | 0.036 |
| 70 - 79 years | 0.029 | 0.033 | 0.032 | 0.037 |
| 80+ years | 0.040 | 0.091 | 0.067 | 0.12 |
| | Until 2021-01-01 | Until 2021-02-01 | Until 2021-03-01 | From 2021-03-01 |
| 0-9 years | 0.0006 | 0.0007 | 0.0014 | 0.0018 |
| 10 - 19 years | 0.001 | 0.0013 | 0.0021 | 0.0026 |
| 20 - 29 years | 0.007 | 0.0087 | 0.01 | 0.011 |
| 30 - 39 years | 0.014 | 0.016 | 0.022 | 0.022 |
| 40 - 49 years | 0.016 | 0.018 | 0.026 | 0.025 |
| 50 - 59 years | 0.03 | 0.032 | 0.039 | 0.038 |
| 60 - 69 years | 0.038 | 0.032 | 0.041 | 0.04 |
| 70 - 79 years | 0.049 | 0.033 | 0.031 | 0.037 |
| 80+ years | 0.14 | 0.14 | 0.064 | 0.043 |

before dividing by the total number of individuals testing positive plus the number of age groups times 0.001. The regional proportions were adjusted using national data in those regions when few tested positive in a given period. This was done by calculating a weighted fraction; the regional fraction got the weight equal to the total number of positive cases during that period, while the national fraction got the weight of 50. Then this adjusted fraction of positive tests by age and region was used together with the age-specific probabilities as described in section S2.5.1 to get a hospitalisation risk per age group and region.

##### S2.5.3 Length of stay in hospital and time to hospital admission

The length of stay (LOS) in hospital for COVID-19 patients may change over time. We therefore allowed changepoints in the hospitalisation parameters. In addition, there may be regional differences, and we thus also included the possibility for separate parameters for certain regions. These periods will here be called time-space periods. The parameters for the time-space periods were estimated separately.

The aim was to estimate distributions for the time from symptom onset to hospitalisation, the LOS for patients with and without ventilator treatment, and for the proportion of hospitalised cases needing ventilator treatment. The LOS for patients needing ventilator treatment was divided into three periods: before, during and after ventilator treatment to obtain estimates of the number of required ventilators per day. For patients with more than one COVID-19 hospitalisation episode, the LOS was set equal to the sum of all admission periods.

We used a proportional hazard Cox model to estimate the parameters for each time-space period, assuming right censoring in time. The survival curves were estimated with time-space periods as an explanatory variable, using the *survfit* function in the R-package *survival*. Specifically, one minus the cumulative distribution function of the negative binomial distributions was fitted to each survival curve by minimising the sum of squared errors, using the function *optim* in R.

To estimate the hospitalisation parameters we used individual-level data from the Norwegian Surveillance System for Communicable Diseases (MSIS) and the Norwegian Intensive Care and Pandemic Registry (NIPaR)[24] as presented in Section S2.1. We include three changepoints, dividing the data into four periods – before June 2020, June 2020-December 2020, January 2021-February 2021, and March 2020-April 12, 2021.

Except for the Oslo region, there were no significant differences in the regional COVID-19 hospitalisation parameters. The hospitals in Oslo increased the use of mechanical ventilators from January 2021. Therefore, we obtained separate estimates for Oslo in the last two time periods. When estimating the time spent under ventilator treatment, we merged the periods January 2021-February 2021 and March 2021-April 12, 2021, to reduce the level of censoring. In a separate project, we are trying to find the causes of these changes, see also [25]). Figures S2, S3, S4, S5 and S6 show for each time-space period a histogram of time to event together with an estimated negative binomial distribution fitting the data in red. The parameters of the negative binomial distribution (mu and size) together with the number of patients (N) in the data are shown in the upper right corner of each panel.

###### S2.5.4 Time from symptom onset to hospitalisation

When estimating time from symptom onset to the first hospitalisation, patients with symptom onset date equal to the COVID-19 test date were disregarded. In Norway, this registration practice is common in situations where symptom onset is unknown. We assumed that these cases were missing at random and hence did not introduce bias. All patients included in this analysis were hospitalised, so no right censoring was needed for this calculation. Figure S2 shows the real data and the fitted distribution in red.

###### S2.5.5 Time from hospitalisation to discharge

The time from hospitalisation to discharge was estimated separately for two groups, those who required ventilator treatment and those who did not. Right censoring occurred in the last time-space period from March 2021; 15 and 65 patients from Oslo and the rest of Norway, respectively, were still in hospital at the end of that period, out of a total of 476 patients from Oslo and 960 from the rest of Norway. The number of patients included in each time-space period is shown in figure S3 together with the real data and the fitted distribution in red.

For those needing mechanical ventilator treatment, we considered three time intervals: time from admission to start of ventilator treatment (figure S4), time on ventilator (figure S5) and time to discharge after ventilator treatment (figure S6). Both the estimation of the distribution of the time on ventilator (figure S5) and time after ventilator treatment (figure S6) included censoring events in the last

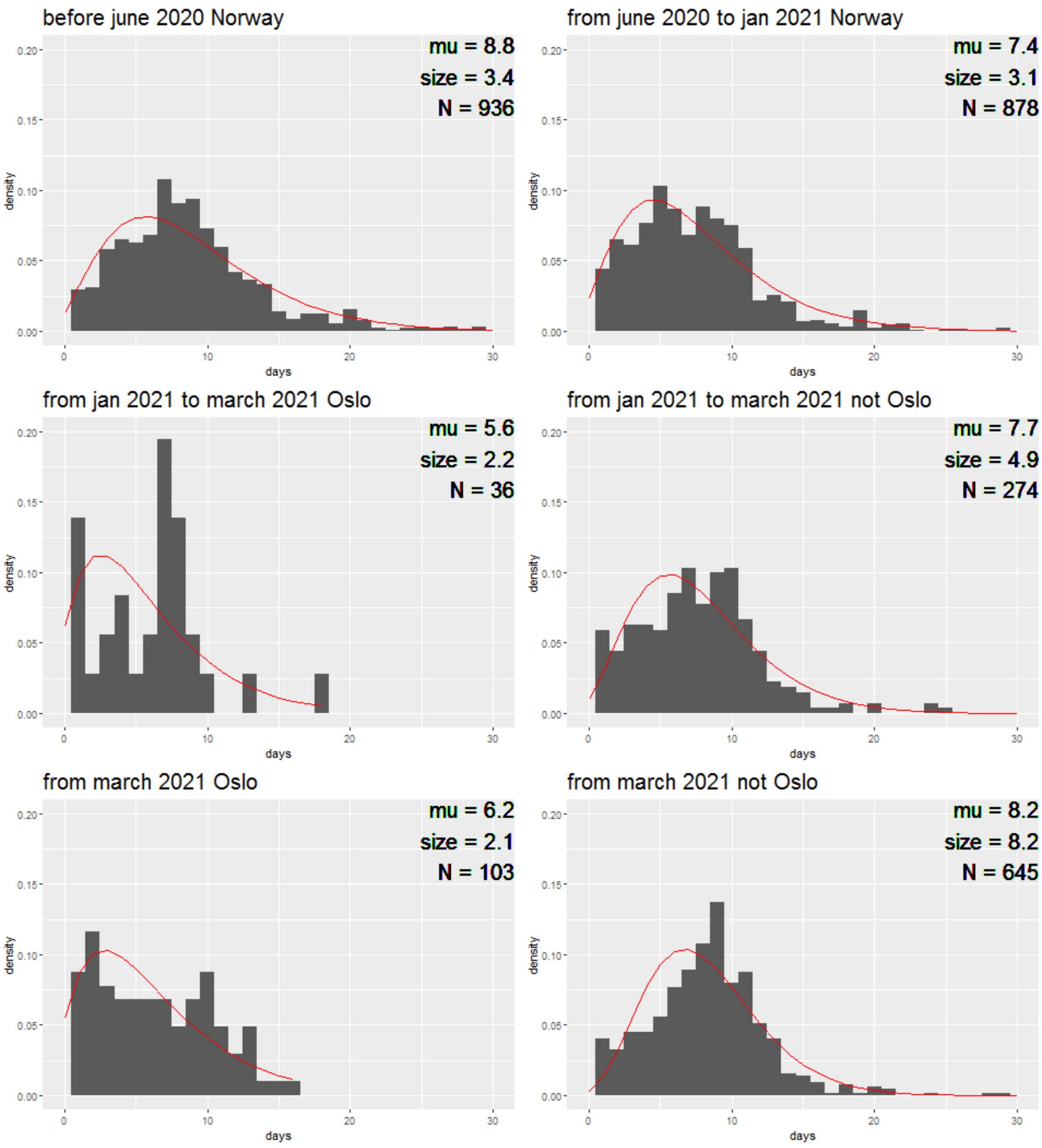


Figure S2: Histogram of time from symptom onset to hospitalisation for different time-space periods. Red line shows the fitted negative binomial distribution described with parameters (mu and size) in the upper right corner. N is the number of observations in each histogram.

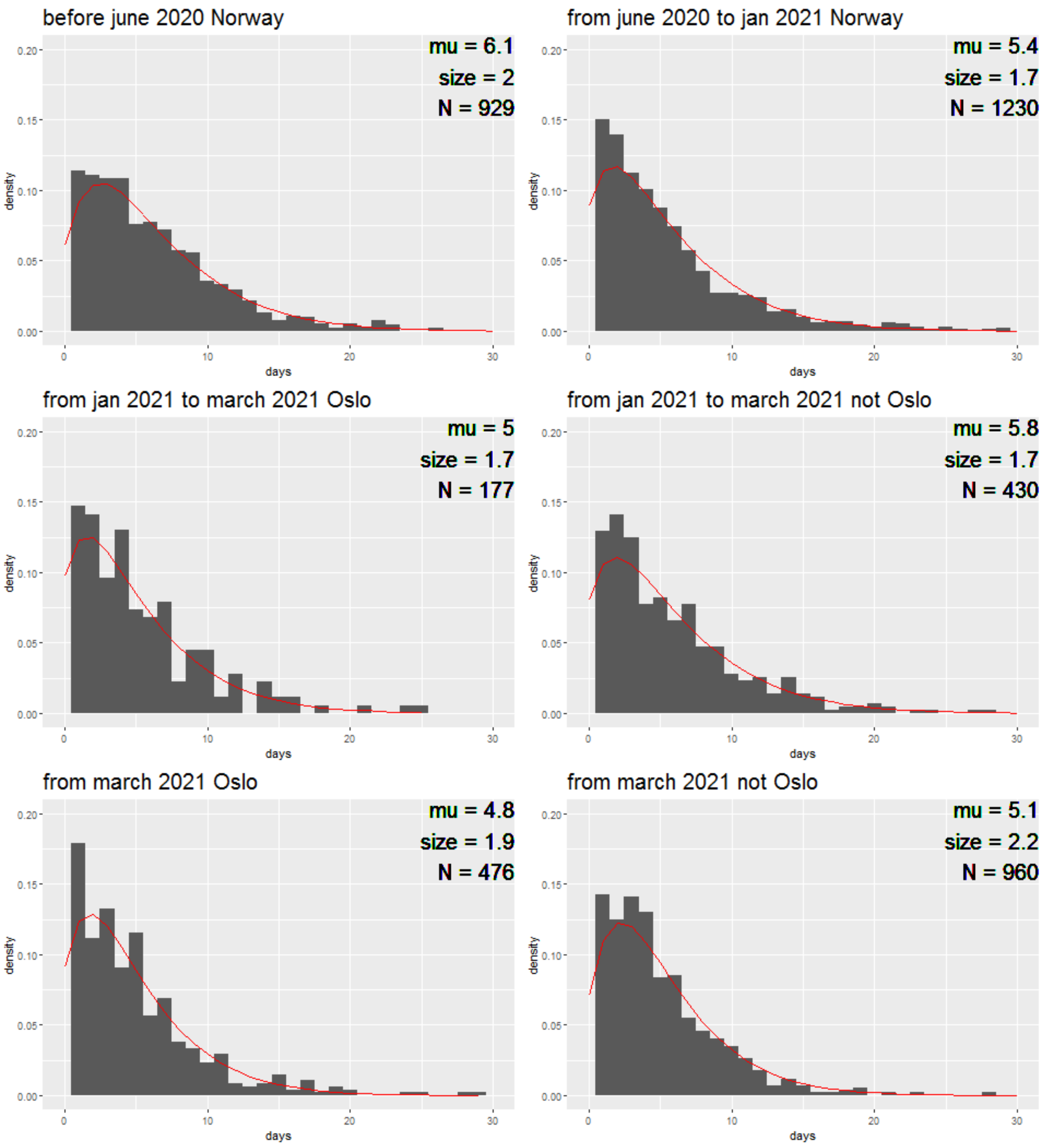


Figure S3: Time from hospitalisation to discharge without respirator treatment for different time and space periods. Red line shows the fitted negative binomial distribution described with parameters (mu and size) in the upper right corner. N is the number of observations in each histogram.

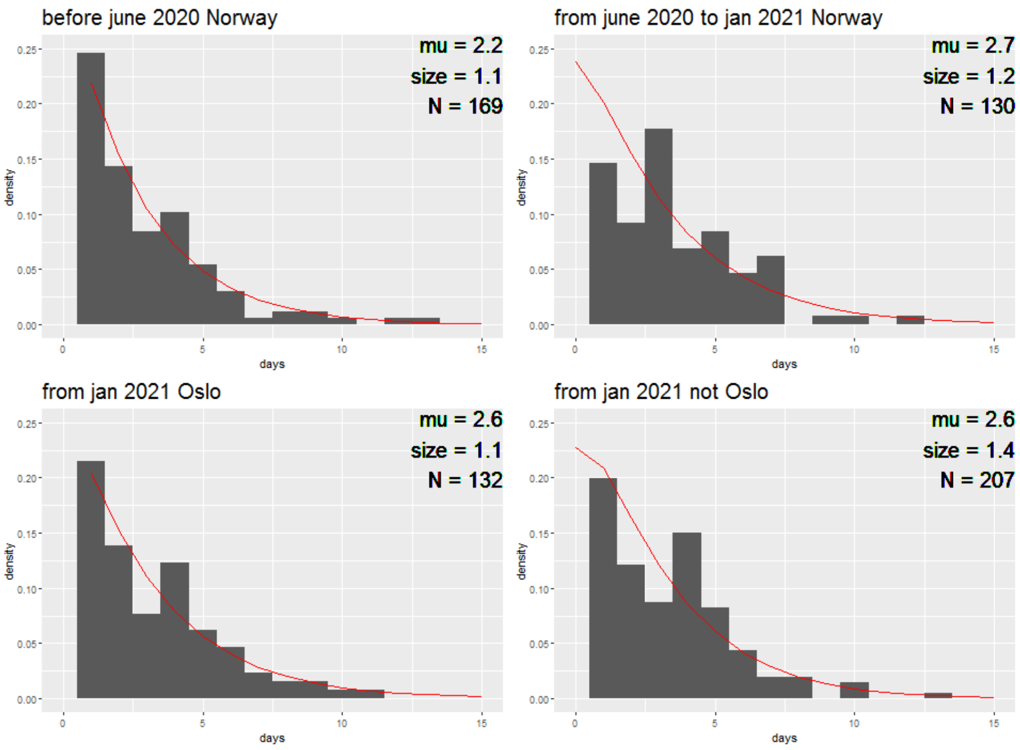


Figure S4: Time from hospitalisation to ventilator treatment for different time and space periods. Red line shows the fitted negative binomial distribution described with parameters (mu and size) in the upper right corner. N is the number of observations in each histogram.

period from January 2021. 13 patients from Oslo and 23 patients from the rest of Norway were censored when estimating time on ventilator, out of a total of 132 patients from Oslo and 207 patients from the rest of Norway. For the time after ventilator treatment, the censored patients were 17 and 28 for the same periods, out of a total of 120 for Oslo and 170 for the rest of Norway.

The proportion of patients needing ventilator treatment has changed over time. From the beginning until June 2020, 0.149 (163 out of 1092) of patients re-

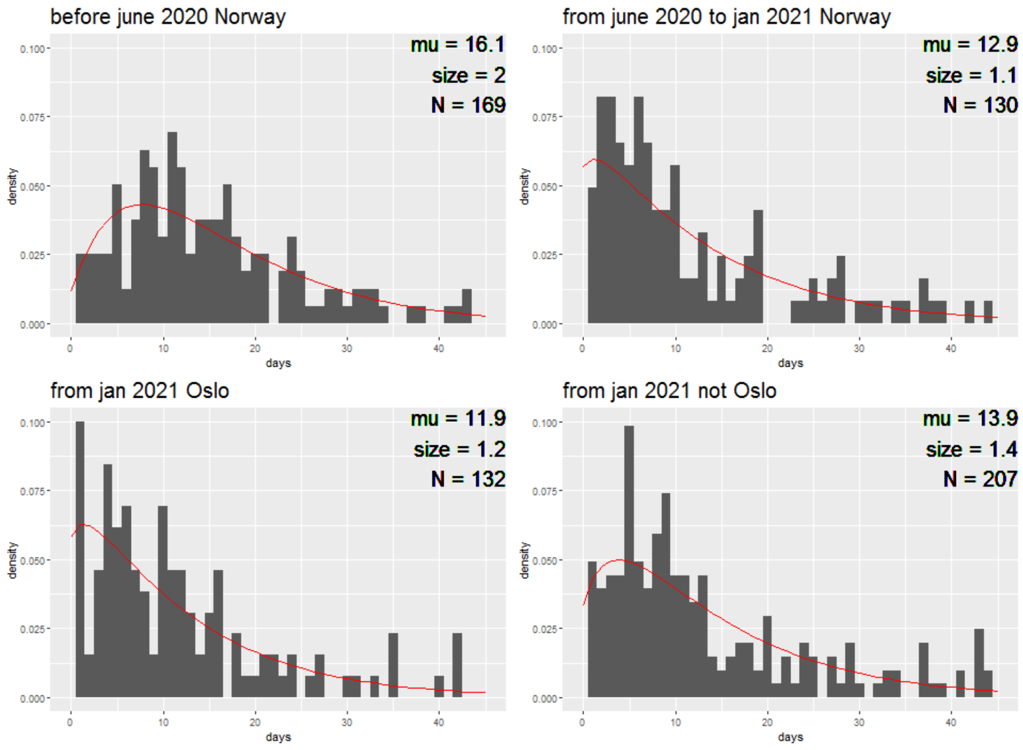


Figure S5: Time on ventilator treatment for different time and space periods. Red line shows the fitted negative binomial distribution described with parameters (mu and size) in the upper right corner. N is the number of observations in each histogram.

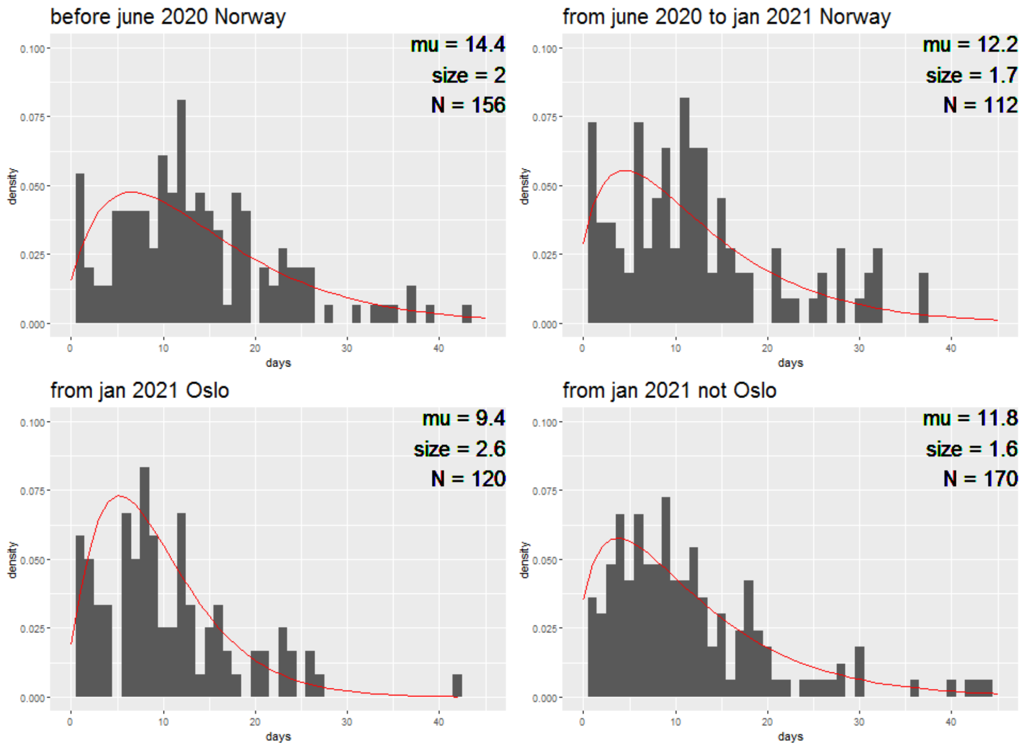


Figure S6: Time after ventilator treatment for different time and space periods. Red line shows the fitted negative binomial distribution described with parameters (mu and size) in the upper right corner. N is the number of observations in each histogram.

quired ventilator treatment. Between June 2020-December 2020 the proportion decreased to 0.098 (133 out of 1363). In the last period, January 2021-April 12 2021, 0.156 (121 out of 774) of patients in Oslo required ventilators, in the rest of Norway the proportion was 0.121 (191 out of 1581).

###### S2.5.6 Increased hospitalisation risk due to the alpha variant

Viral variants sometimes impact the hospitalisation risk. In these cases, the estimates in[23] need to be updated. This was the case of the alpha variant (also called the B.1.1.7 variant). See also [26] for evidence of increased transmissibility of the alpha variant in Norway compared to the previously circulating Norwegian variants. The two first detected cases of the alpha variant imported to Norway occurred on December 27 2020, for two individuals returning from the United Kingdom [27]. Since then, Norway experienced an increasing proportion of cases attributed to the alpha variant. As the alpha variant is also associated with more severe disease and a higher risk of hospitalisation[28, 29] compared to previously circulating strains, we need to adjust for a gradually increasing hospitalisation risk from January 2021 in Norway. For this purpose, we assumed a logistic increase in hospitalisation risk between January 1 2021 and March 31 2021, assuming a total increase of 60%, resulting in the formula

$$\frac{1000\exp(-0.073t) + 1.6 \cdot 10\exp(0.073t)}{1000\exp(-0.073t) + 10\exp(0.073t)}.$$

The slope in the formula was estimated from the test data. In the model with national transmissibility, we assume an increase of 9% in January, 50% in February and 60% from March 2021 corresponding to the monthly values of the logistic increase assumed regionally, as the national model assumes monthly hospitalisation risks instead of daily hospitalisation risks.

#### S2.6 Laboratory-confirmed cases

To calibrate to the observed number of laboratory-confirmed cases, we simulate how many cases are detected by testing (positive test cases). We assume that the number of positively tested cases can be modelled as a binomial process of the simulated daily total incidence of symptomatic and asymptomatic cases, and with success probability $\pi_t$. We also assume a delay $d$ between the day of test and the day of infection. In order to make the hospitalisation data and the test data consistent, we need to capture the changes in test criteria, and we choose to model this in the detection probability $\pi_t$, as

$$\pi_t = \exp\left(\pi_0 + \pi_1 \cdot k_t\right)/(1 + \exp(\pi_0 + \pi_1 \cdot k_t)),$$

where $k_t$ is a 7-days backwards moving average of the total number of tests performed (with both positive and negative results), and $\pi_0$ and $\pi_1$ are two parameters that we estimate, assuming positivity of $\pi_1$.

In Norway, the testing criteria and capacity have changed significantly since early in the epidemic. Therefore, we only calibrate to the test data from May 1 2020. The resulting estimated detection probability is provided in figure S7. We see that it has been increasing since May 2020.

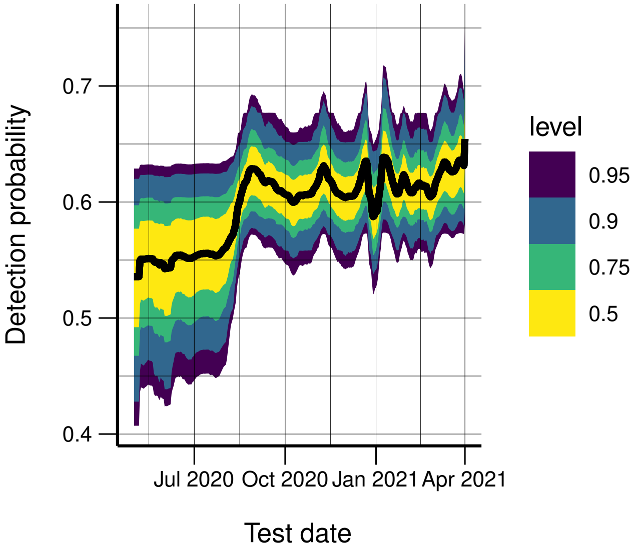


Figure S7: Estimated probability to detect a case by testing.

#### S2.7 Parameter assumptions and uncertainties

A lot is uncertain about the natural history of SARS-CoV-2. Hence, there are many unknown parameters in the model that our method estimates (or calibrates) from data. We calibrate the amplification factor and the transmission parameters $\beta_t^i$, which are directly related to the reproductive numbers. We also calibrate the parameters $\pi_0$ and $\pi_1$ in the model for the detection of positive tests, and the delay $d$ between transmission and testing. As described above, we estimate the hospitalisation parameters from individual-level data on hospitalised cases.

The remaining parameters related to the natural history of COVID-19 are assigned fixed values or distributions based on values found in the literature. We base the natural history parameters on estimates from[18], with an adjustment of the presymptomatic period based on[20]. Specifically, we assume a latent period exponentially distributed with expected duration of three days, a presymptomatic period exponentially distributed with expected duration of two days, an infectious period (time in $I$ and $I_a$) exponentially distributed with expected duration of five days, a probability of asymptomatic infection of 40%, a relative infectiousness of presymptomatic of 1.25 compared to the infectiousness in $I$ which is 1, and a relative infectiousness of 0.1 of the asymptomatic cases. Our assumptions are also in agreement with[30], who found an incubation period for COVID-19 of 5.1 days. All the parameter choices and values are provided in table S2.

Table S2: Parameter assumptions

| Assumptions | Mean | Distribution | Reference |
|---|---|---|---|
| **Model parameters** | | | |
| Exposed period ($1/\lambda_1$) | 3 days | Exponential | [18] |
| Presymptomatic period ($1/\lambda_2$) | 2 days | Exponential | [18] |
| Symptomatic infectious period ($1/\gamma$) | 5 days | Exponential | [18] |
| Asymptomatic, infectious period ($1/\gamma$) | 5 days | Exponential | [18] |
| Infectiousness asympt. ($r_{I_a}$) | 0.1 | Fixed | [18] |
| Infectiousness presymp. ($r_{E_2}$) | 1.25 | Fixed | [18] |
| Probability asymptomatic ($p_a$) | 0.4 | Fixed | [18] |
| **Healthcare** | | | |
| % symptomatic and asymptomatic infections requiring hospitalisation: | | Fixed | [23] corrected for: proportion of elderly living in elderly homes in Norway (last two age groups), and age-profile in laboratory-confirmed cases since May 1 2020. Corrected values available in table S1. |
| 0-9 years | 0.1% | | |
| 10 - 19 years | 0.1% | | |
| 20 - 29 years | 0.5% | | |
| 30 - 39 years | 1.1% | | |
| 40 - 49 years | 1.4% | | |
| 50 - 59 years | 2.9% | | |
| 60 - 69 years | 5.8% | | |
| 70 - 79 years | 9.3% | | |
| 80+ years | 22.3% | | |
| Overall hospitalisation risk | 2.26% | Fixed | [23] (adapted to Norwegian demography) |

#### S2.8 Calibration

In the model for Norway with a national transmissibility, we estimate the transmissibility parameters $\beta_t$, the parameters $\pi_0$, $\pi_1$, in the expression for the detection probability, the delay $d$ between transmission and test, and the amplification factor for the imported cases. As of April 1 2021, this results in 19 nationally estimated parameters because of 14 changepoints in the transmission. In the model with regionally varying transmission parameters $\beta_t^i$, we in addition calibrate these region-specific transmissibility parameters. For Norway, as of April 1 2021, we use in total 94 regional transmissibility parameters. Because the regional calibration is a much more difficult estimation problem than the national calibration, we first estimate the amplification factor, $\pi_0$, $\pi_1$, and $d$ from the national calibration and use the obtained posterior distributions of these parameters as priors for the same parameters in the regional calibration.

We developed a new sequential Monte Carlo version of Approximate Bayesian Computation (SMC-ABC) to estimate the parameters of our model. The idea of ABC is to identify parameters that produce simulated data that are close to the observed data, measured in some distance. We propose to use the least squares between the simulated regional hospital incidence and the observed regional hospital incidence, summed over all the regions. Similarly, we calculate the sum of the least squares distance between the observed and simulated test incidence by region.

This, however, is not sufficient to give good results in all regions when there is heterogeneity in the level of transmission between the various regions, as is often the case. For example, more populous and more densely populated regions tend to experience more cases than smaller and less urban regions. Therefore, in the regional calibration, we compute a vector of multiple distances, one for the sum of the larger/more densely populated regions and one with the sum of the distances for all other regions. In this way, as all components of this vector are minimised in the ABC, we get a good overall fit, not only in the regions with most cases, which would have the greatest weight in a sum. In SMC-ABC[31], the parameters are calibrated in subsequent rounds, where the distances decrease in each round. We thus use four separate tolerances for the distances, ensuring that they all reduce in each round (hospitalisation incidence in the largest regions, hospitalisation incidence for the rest of the regions, laboratory-confirmed cases in the largest regions, and laboratory-confirmed cases in the rest of the regions). In Norway, infection rates have been consistently high in the densely populated region of Oslo and the neighbouring populous Viken region, and we therefore use a separate tolerance measure for these two regions combined.

In the first round, we start with predefined thresholds for the distances, and a prior for the parameters. Candidate parameter values are drawn from the prior, and we simulate the hospitalisation and positive test time series for each region using these candidate parameters. We accept the candidate parameters if all the distances are below the prespecified thresholds. For all subsequent rounds, each threshold is chosen as the 0.80-quantile of the distances for the accepted parameters in the previous round. Candidate parameters are now sampled from the set of chosen parameters from the last round using importance weights. To add further exploration, these candidate parameters are perturbed with a multivariate normal distribution with covariance equal to the empirical covariance matrix of the parameters retained in the previous round. We obtain 1000 parameter sets in each round when we calibrate the regional parameters, 200 when we calibrate the national parameters. We summarise the algorithm in S1, see also [15]. In the algorithm, the observed hospital incidence data are denoted by $\mathbf{H}$ and the observed positive tests as $\mathbf{T}$. The corresponding simulated hospital incidence is denoted by $\mathbf{H}'$ and the simulated positive tests as $\mathbf{T}'$, and $\bar{\cdot}^7$ denotes a 7-days backwards moving average. Subscript $OV$ denotes that the error is calculated for Oslo and Viken (the largest regions), while $-OV$ means all counties except Oslo and Viken. The algorithm is almost the same for the national calibration, except that we calculate the errors for the simulations aggregated over regions and compare it to the nationally aggregated data. Hence there are only two thresholds for the national algorithm, one for the hospitalisation incidence and one for the test data.

For the test data, we choose to use a 7-days backwards moving average for the covariate $k_t$, and to calculate the distance between the observed number of positive tests and the simulated ones. This is done to take into account potential day-of-the-week-effects. This allows for less day-to-day variance than using the daily data directly.

As our prior distribution, we assume an independent normal prior for all the variables except the delay between transmission and a positive test, which is assumed to be a uniform integer between 0 and 4. For the reproduction numbers, we use the same prior mean and variance for all the regions. For Norway, the mean

**Algorithm S1** ABC-SMC

**Initialise:**

Set a starting value $\epsilon_0^{hOV}$, $\epsilon_0^{h-OV}$, $\epsilon_0^{tOV}$, $\epsilon_0^{t-OV}$ for the tolerances for the hospitalisation data in the largest regions and for the rest of the regions, and test data in the largest regions, and the rest of the regions, respectively, $r = 1$ and $\mathbf{w}^1 = (1, \dots, 1)$.

set $i = 0$.
**while** $i < 1000$ **do**
  **if** $r = 1$ **then**
    sample parameters $\theta_i$ from the prior $\pi$.
  **else**
    Sample $\theta^p$ from $\theta^{r-1}$ with weights $\mathbf{w}^{r-1}$.
    Propose $\theta_i$ from a normal distribution $N$ centred at $\theta^p$ with variance equal to the empirical variance from the previous round.
    If $\pi(\theta_i) = 0$, sample a new $\theta_p$.
  **end if**

Run the model with $\theta_i$, providing the simulated number of hospitalised cases $\mathbf{H}'$, and positive tested $\mathbf{T}'$.

**if** $f^h(\mathbf{H_{OV}}', \mathbf{H_{OV}}) = ||\mathbf{H_{OV}}' - \mathbf{H_{OV}}||_2 < \epsilon_r^{hOV}$ and $f^h(\mathbf{H_{-OV}}', \mathbf{H_{-OV}}) = ||\mathbf{H_{-OV}}' - \mathbf{H_{-OV}}||_2 < \epsilon_r^{h-ov}$, $f^t(\mathbf{T_{OV}}', \mathbf{T_{OV}}) = ||\bar{\mathbf{T}}_{\mathbf{OV}}'^{7} - \bar{\mathbf{T}}_{\mathbf{OV}}^{7}||_2 < \epsilon_r^{tOV}$ and $f^t(\mathbf{T_{-OV}}', \mathbf{T_{-OV}}) = ||\bar{\mathbf{T}}'^{7}_{\mathbf{-OV}} - \bar{\mathbf{T}}_{\mathbf{-OV}}^{7}||_2 < \epsilon_r^{t-OV}$, **then**

set $\theta_i^r = \theta_i$, and calculate weights $\mathbf{w}_i^r$ for the parameters, as

$$\mathbf{w}_i^r = \begin{cases} 1, & \text{if } r = 0, \\ \frac{\pi(\theta_i)}{\sum_{j=1}^{n} w_j^{r-1} N(\theta_i | \theta_j^{r-1})}, & \text{otherwise,} \end{cases}$$

Increment $i = i + 1$.
  **end if**
**end while**
normalise the weights as $\mathbf{w}_i^r = \frac{w_i^r}{\sum_j w_j^r}$.
set $\epsilon_{r+1}^{hOV}$ to the 80th percentile of $f^h(\mathbf{H_{OV}}', \mathbf{H_{OV}})$ for the accepted parameters.
set $\epsilon_{r+1}^{h-OV}$ to the 80th percentile of $f^h(\mathbf{H_{-OV}}', \mathbf{H_{-OV}})$ for the accepted parameters.
set $\epsilon_{r+1}^{tOV}$ to the 80th percentile of $f^t(\mathbf{T_{OV}}', \mathbf{T_{OV}})$ for the accepted parameters.
set $\epsilon_{r+1}^{t-OV}$ to the 80th percentile of $f^t(\mathbf{T_{-OV}}', \mathbf{T_{-OV}})$ for the accepted parameters.
Increment $r = r + 1$.

is assumed to be 3.7 with variance 0.4 for $R_0$, and 1.0 with variance 0.25 for the other reproduction numbers. For the amplification factor, we assume a mean of 1.3 and a variance of 0.25. For $\pi_0$ and $\pi_1$, we assume mean 0 and variance 4.0 and $25 \cdot 10^{-9}$, respectively. We truncate the normal distribution at 0 for the reproduction numbers, amplification factor, and $\pi_1$. The priors for $R_0$ and the amplification factor are based on early assessments on hospital prevalence data for Norway. The priors for the other transmissibility parameters have been centred at 1 as a neutral value, with a rather large variance to cover the range of reasonable values.

###### S2.8.1 Split-SMC-ABC

The regional calibration is challenging, as it is a very high-dimensional calibration problem. With the standard SMC-ABC, we were not able to obtain convergence in useful time for our almost 100 parameters. Therefore, we developed a new version of the SMC-ABC to produce reliable and timely results, which we call the Split-SMC-ABC.

The simple idea is to treat the past estimated reproduction numbers as fixed, and use all the computational time to estimate the most recent reproduction numbers, the key parameters for situational awareness. However, parameters that enter the earlier part of the epidemic model are also important, as the more recent reproduction numbers (or equivalently $\beta_t^i$) depend on them. For example, a higher-than-average reproduction number in one period is typically followed by a lower-than-average reproduction number in the following period, to compensate for the high number of cases produced in the first period. In addition, the whole history is necessary for estimating the total cumulative number of infections and hence the immunity of the population. Therefore, since the present parameters depend on the past parameters, we cannot treat the two calibration problems (past and present) as independent. Instead, we run the past until convergence and use the obtained posterior distributions when calibrating the more recent period. In addition, we need to handle the transition period between the past period and the more recent period. Here, we exemplify the algorithm with the dates used in the simulations in this paper, but the approach is, of course, general. We focus on the split into two parts, which we call batches. Note that this operation can be repeated multiple times, generating multiple batches, by following the same procedure.

We start with the first batch calibration, obtained by calibrating to all the data up to August 15, 2020. From this calibration, we obtain 1000 posterior samples of transmissibility parameters up to August 1 2020, $\pi_0$, $\pi_1$, $d$, and the amplification factor. We also have the 1000 corresponding epidemic time trajectories of $S^i$, $E_1^i$, $E_2^i$, $I^i$, $I_a^i$, and $R^i$ in each region $i$. The first batch calibration contained a changepoint on August 1 2020, in all regions. We denote this timepoint, August 1 2020, by $t_{cal}$, which is the first date of the data we wish to calibrate to in the second batch. Hence, all the parameters prior to $t_{cal}$ are assumed to follow the posterior distributions estimated in the first batch calibration. We also fix the time-independent parameters $\pi_0$, $\pi_1$, $d$, and the amplification factor to follow their posterior distribution from the first batch. In the second batch calibration, we calibrate all transmissibility parameters entering the model after $t_{cal}$.

Note that the first batch calibration was obtained by calibrating to data up to

$t_{cal} + 15$. This is important due to the posterior dependence between consecutive transmissibility parameters. As the observed hospitalisations and laboratory-confirmed cases on $t_{cal}$ correspond to transmission events that occurred a certain number of days before, we start the simulations on the day $t_{cal} - 15$. 15 days cover approximately 90% of the expected hospitalisations in the simulations (which are more delayed than the positive tests). Hence, we keep the 1000 calibrated trajectories (along with the corresponding calibrated reproduction numbers) from the first batch calibration, up to the day $t_{cal} - 15$. The simulations in the second batch are started on the exact states ($S^i$, $E_1^i$, $E_2^i$, $I^i$, $I_a^i$, and $R^i$ in each region $i$) that were obtained in the first batch calibration, on the date $t_{cal} - 15$. When continuing one of the 1000 past trajectories from $t_{cal} - 15$ to $t_{cal}$, the corresponding most recent reproduction number is used.

In the second batch, we calibrate only parameters from $t_{cal}$, using all the available data from $t_{cal}$. In the first round $r = 1$, we first sample uniformly which of the 1000 trajectories produced in the first batch we should continue (with the corresponding amplification factor, $\pi_0$, $\pi_1$, $d$, and reproduction number for the first 15 days for that trajectory). We continue to run with different samples of the past trajectories and the new parameters from the second batch, until we have 1000 accepted parameter sets with errors lower than the prespecified thresholds for the test data and the hospital data from $t_{cal}$ until the latest data point.

For all subsequent rounds, $r > 1$, we first sample which past trajectory we should run. The sampling probabilities $S(p)$ for each past trajectory $p$ are calculated as

$$S(p) = (1/1000 + N_{r-1}(p))/1001,$$

where $N_{r-1}(p)$ is the number of times trajectory $p$ was chosen in the previous round $r-1$, then normalised to sum to 1. Hence all past trajectories have non-zero probability of being chosen. The probability of selecting a specific trajectory increases linearly with how frequently it was accepted in the previous round. We divide by 1001 to ensure that the probabilities sum to 1. Then we sample from the 1000 accepted parameter sets for the second batch from the accepted parameter sets of the previous round in the usual way, as described in Algorithm S1.

Using the split method, we lose some of the dependence structure between the past and the most recent reproduction numbers. We limit this effect by letting the last, fixed reproduction number be informed by future data in a period of 15 days after $t_{cal}$. As mentioned, this has motivation and works well in practice in the case of Norway. Other lengths of the overlap can be tested in other cases. From a theoretical point of view, it is difficult to quantify how much stochastic dependence is lost in this way.

For Norway, we have two additional temporal split points in the calibration where we consider the past fixed, resulting in four different batches. One before August 2020, one between August 2020 and November 5 2020, one between November 5 2020 and January 4 2021, and one after January 4 2021.

##### S2.8.2 Regionally separate prior calibrations

We noticed that this calibration method, when ran for a reasonable number of rounds, produced parameter estimates which typically led to good fits for the most populated regions and/or regions with more cases. In comparison, the fit was not particularly good for the regions with fewer cases.

As already mentioned, we introduced a vector of calibration distances in the SMC-ABC, with one distance measure for the cases in large regions and one combining the rest of the regions, by summing their distance measures. To ensure a good fit in all regions, we would have preferred to introduce one distance measure for each region. However, it is not feasible to work with such a high-dimensional error measure, as the probability of sampling a parameter vector that improves the fit in all regions simultaneously is low. Hence, such a high dimensional error measure would result in convergence issues. Therefore, we propose a two-step version of the split-SMC-ABC for the final batch.

We first perform one separate calibration for each region. This is done using the same disease spread model and setup as previously described, but assuming a common, national transmissibility for all the other regions, except the one of interest. We then take the separately calibrated transmissibility parameters for each region and use their posterior distribution as prior distributions in the regional calibration which simultaneously calibrates the transmissibility for all the regions. This last step is necessary to learn the needed spatial correlations, as the cases in the different regions are dependent due to the mobility between the regions. The algorithm for the separate calibrations for each region is the same as the algorithm provided in Algorithm S1, except that the error is separated into the region of interest and the rest of the regions, instead of the largest regions and the rest of the regions.

### S3 Supplementary results

#### S3.1 Validation results

The validation scores are provided in table S3.

Table S3: Validation results comparing the regional and national model.

| | National | | | Regional | | |
|---|---|---|---|---|---|---|
| | Week 1 | Week 2 | Week 3 | Week 1 | Week 2 | Week 3 |
| **Hospital** | | | | | | |
| region Energy Score | 25.62 | 30.81 | 31.51 | 13.78 | 16.29 | 18.97 |
| National CRPS | 4.52 | 16.46 | 32.12 | 21.75 | 23.03 | 18.48 |
| Coverage 50% prediction interval | 0.44 | 0.36 | 0.35 | 0.64 | 0.45 | 0.56 |
| Coverage 95% prediction interval | 0.62 | 0.56 | 0.62 | 0.85 | 0.85 | 0.80 |
| **Confirmed Cases** | | | | | | |
| region Energy Score | 754.81 | 839.09 | 860.36 | 380.28 | 529.86 | 704.24 |
| National CRPS | 547.20 | 784.84 | 893.63 | 674.01 | 612.28 | 644.91 |
| Coverage 50% prediction interval | 0.13 | 0.22 | 0.22 | 0.24 | 0.25 | 0.18 |
| Coverage 95% prediction interval | 0.36 | 0.45 | 0.51 | 0.55 | 0.64 | 0.64 |

#### S3.2 Fit to test and hospitalisation data

When estimating the transmissibility, we choose to use both the hospitalisation data from the beginning of the epidemic and the test data from May 2020, as both contain information. The fit to the regional hospitalisation data until April 2021, along with three-weeks ahead predictions, is provided in figures S8-S9, for the model allowing region-specific transmissibilities. The fit to the test data is provided in figures S10-S11. Note that the test data are more volatile than the hospitalisation incidence due to flare-ups of local outbreaks that become controlled. For this reason, test data are not well captured by the constant step-function in the transmissibility unless the dates for the changepoints are tailored to the dates of the outbreaks (see for example Agder).

However, they do contain information on the transmissibility, in particular for the more recent days. We note that for the first days of prediction, our hospital incidence predictions seem to cover the real data that are later observed, but that we start overpredicting over time. This is due to local restrictions implemented in Oslo on March 17 2021, and national restrictions implemented on March 25 2021, which have reduction effects that we have not captured with our most recently estimated effective reproduction number, which is active from March 2 2021. This is particularly clear in the fit to the largest region in Norway, Viken, and in the national aggregate.

Note that we do not capture all the uncertainty in the test data. This is likely because we calibrate to a 7-days-backwards moving average of the daily test data. Moreover, the binomial assumption could be too simple, and a beta-binomial distribution would allow more variance, at the cost of an additional parameter.

#### S3.3 Fit to data up to March 2021

We include also the fit to the test data and hospitalisation data for the calibration up to March 2021. Here, no changepoints later than February 8 2021 were used.

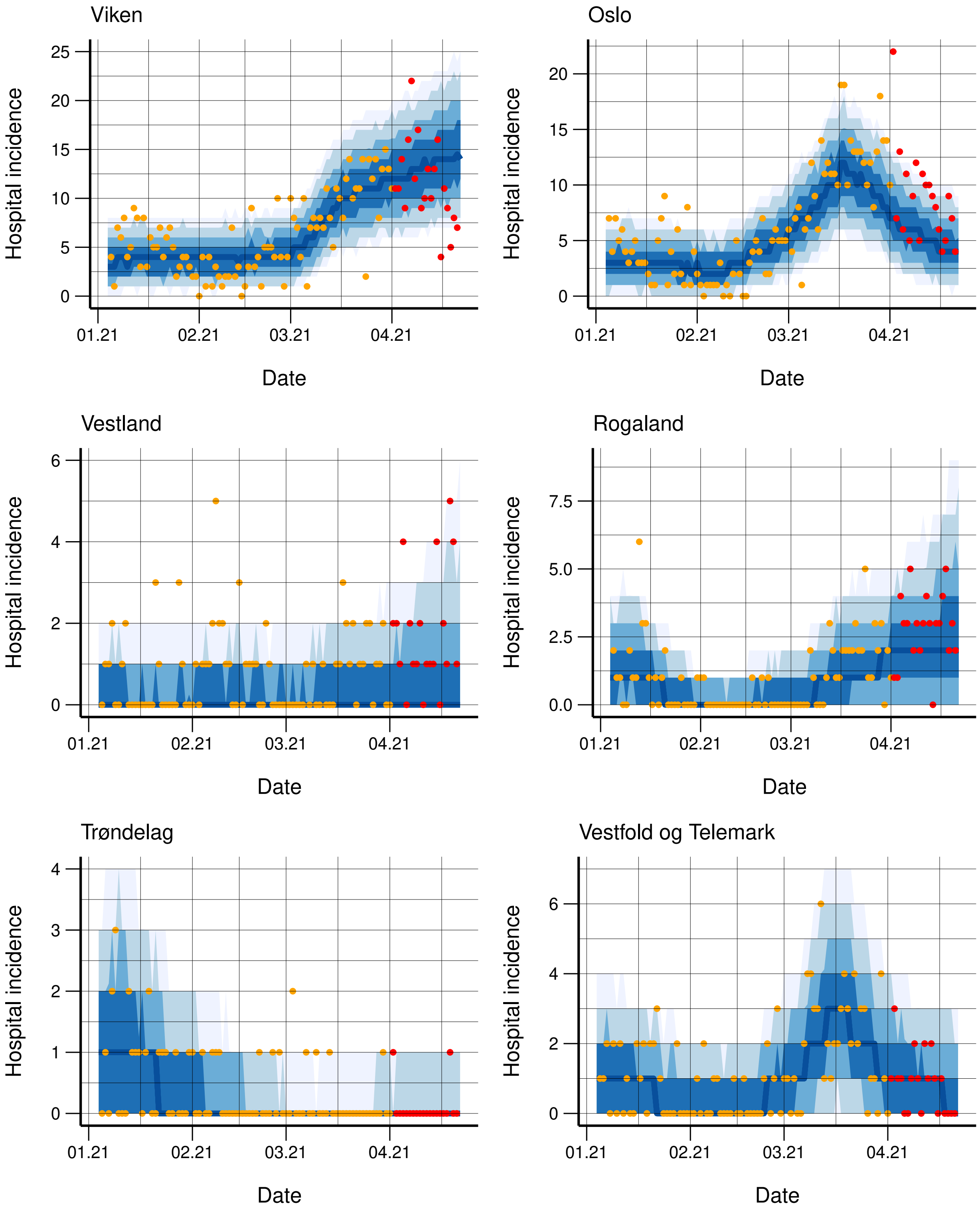


Figure S8: Observed (orange dots) and simulated hospitalisation incidence for each region, along with 95%, 90%, 75%, and 50% credibility intervals. The regions are ordered according to population, with the capital Oslo as second. The incidence for the next three weeks not used in the calibration is provided in red.

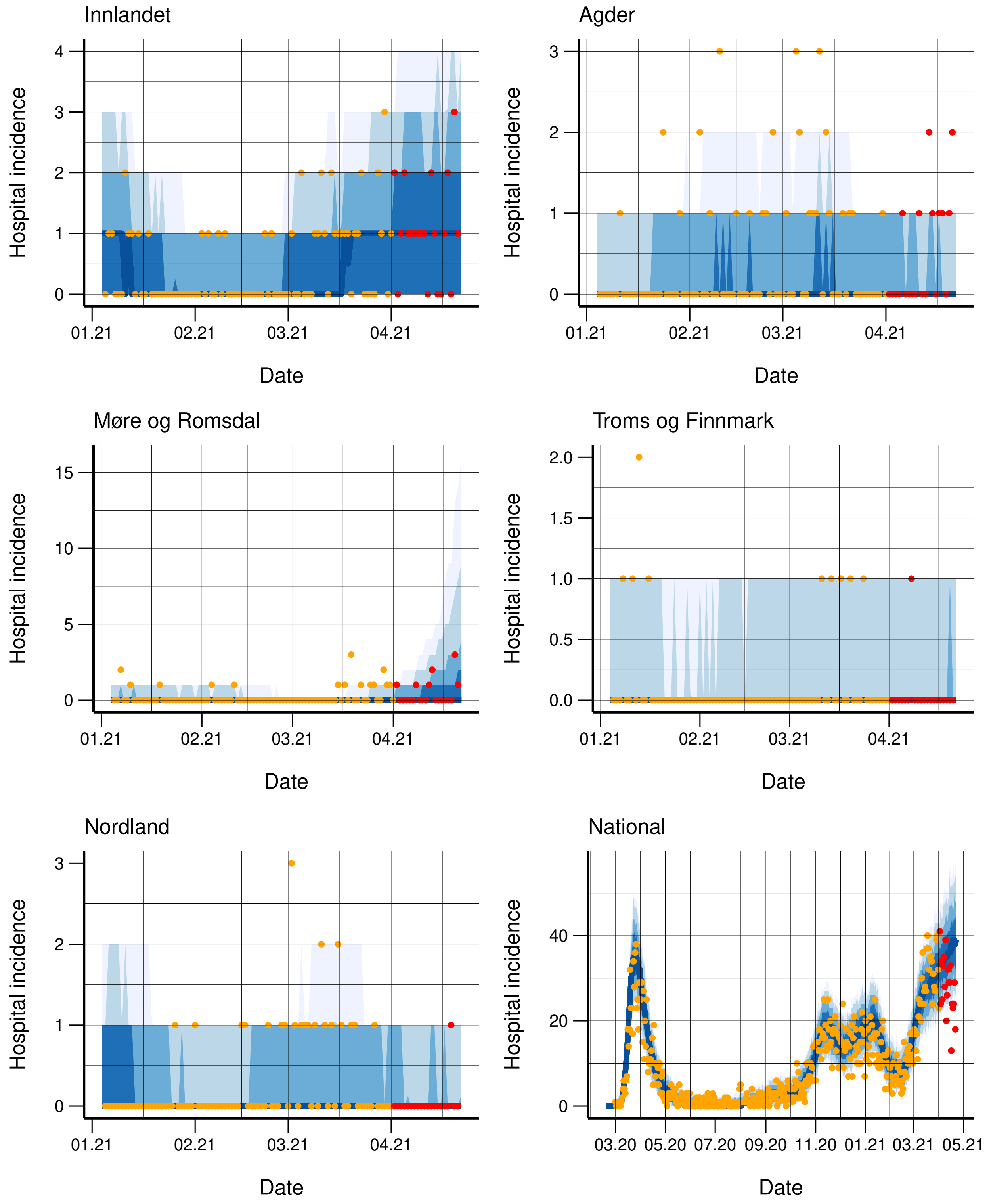


Figure S9: Observed (orange dots) and simulated hospitalisation incidence for each region, along with 95%, 90%, 75%, and 50% credibility intervals. The regions are ordered according to population. In the lower right panel, we have the national fit for the model assuming the same transmissibility in all counties. The incidence for the next three weeks not used in the calibration is provided in red.

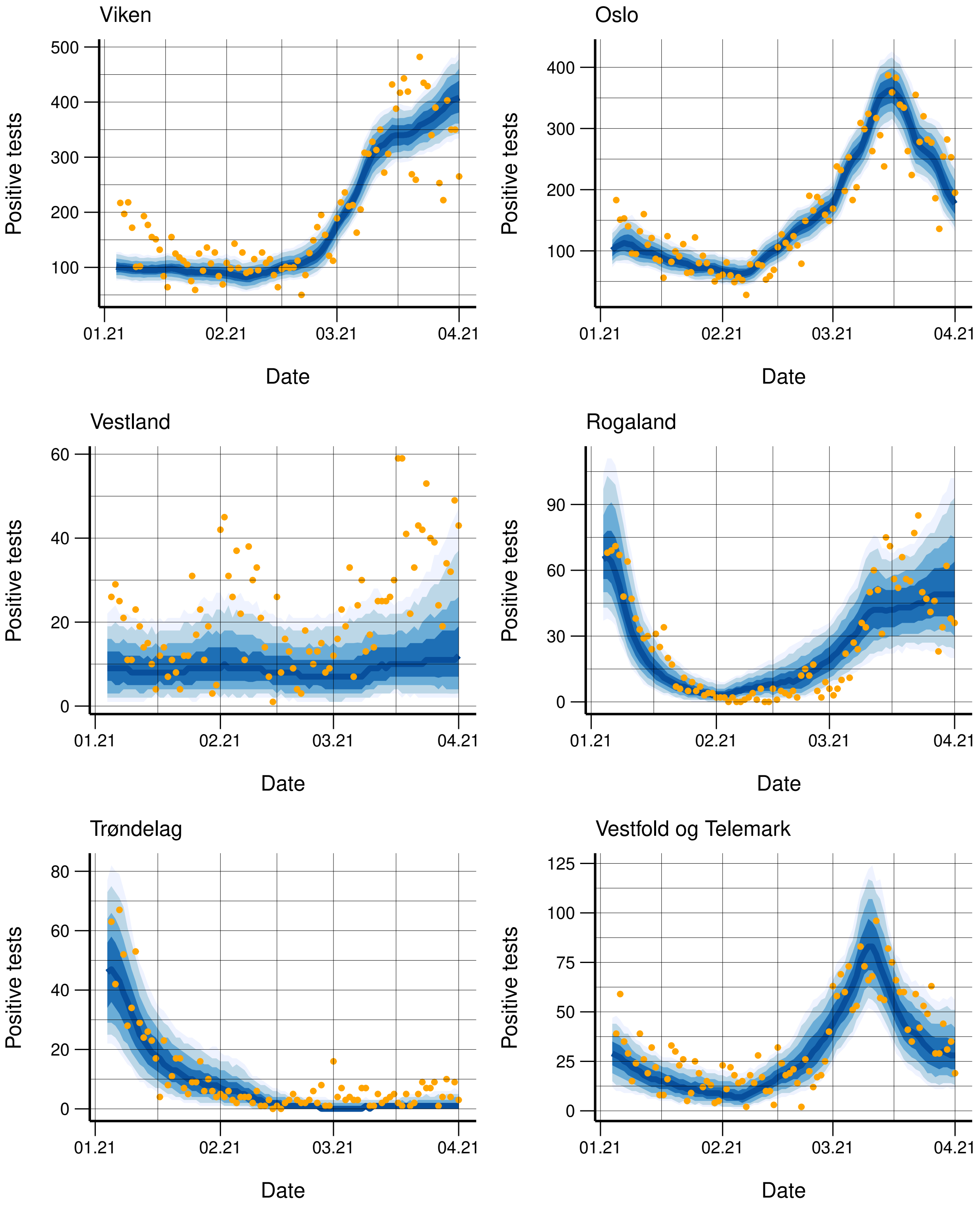


Figure S10: Observed (orange dots) and simulated positive tests for each region, along with 95%, 90%, 75%, and 50% credibility intervals. The regions are ordered according to population, with the capital Oslo as second.

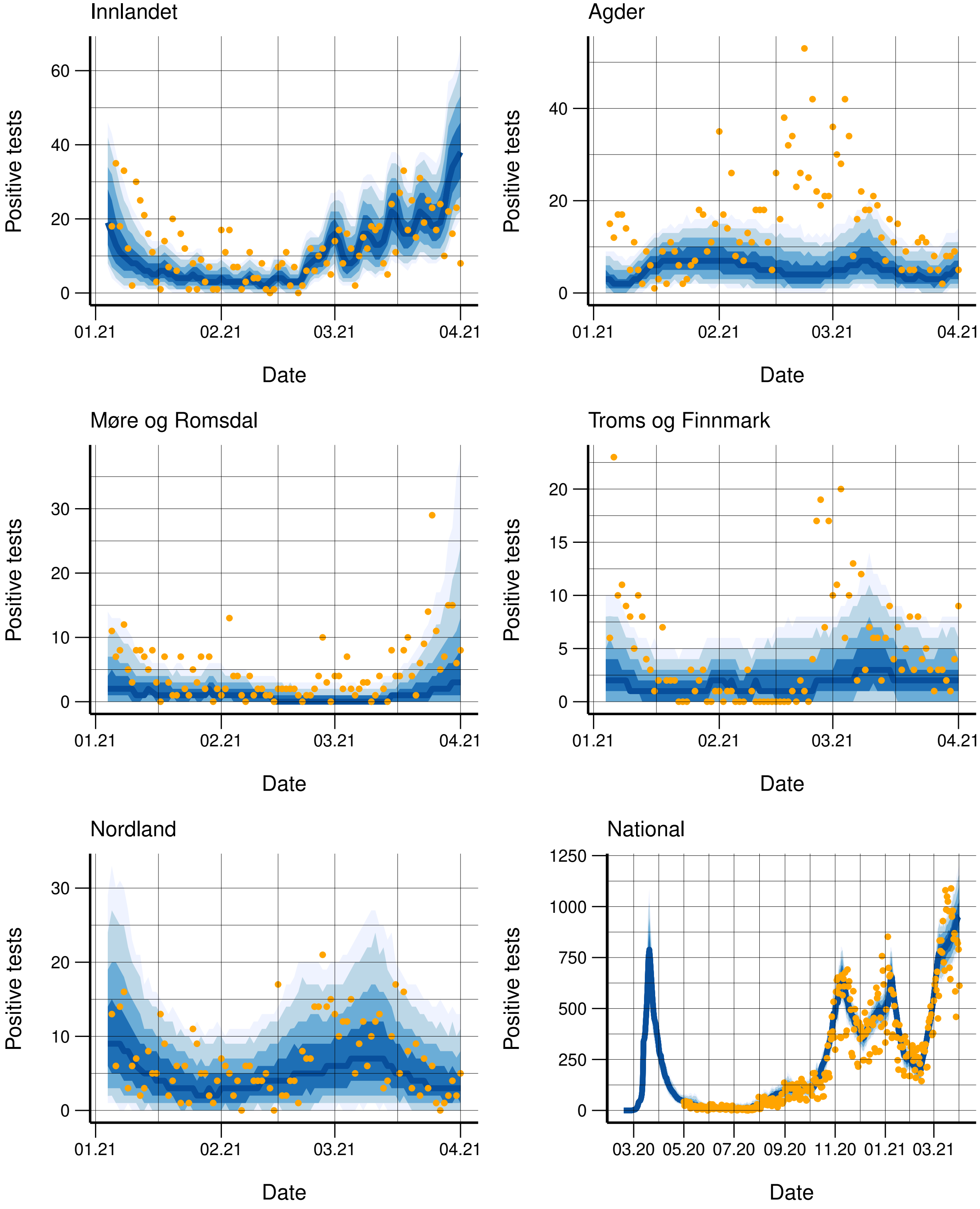


Figure S11: Observed (orange dots) and simulated positive tests for each region, along with 95%, 90%, 75%, and 50% credibility intervals. The regions are ordered according to population. In the lower right panel, we have the national fit for the model assuming the same transmissibility in all counties.

The fit to the regional hospitalisation data until March 1, along with three-weeks ahead predictions, is provided in figures S12-S13, for the model allowing region-specific transmissibilities. The fit to the test data is provided in figures S14-S15. The predictions of future hospitalisations are better here than for April 1 2021, as the intervention measures implemented later in March take more time before the effect shows up in the data. We note however that we are underestimating the hospitalisation a bit, in particular in Viken.

We repeat the calibration up to March 1 2021 using only hospitalisation data. The fit to the hospital incidence is provided in figures S16-S17. If we consider Viken, we note that the hospitalisation predictions are better in this setting when only calibrating to the hospitalisation data, than when we calibrate to both the hospitalisation data and the test data, when we underpredicted the hospitalisation incidence (figures S12-S13). However, the variance is also larger.

#### S3.4 Fit to data up to November 1 2020

We include also the fit to the test data and hospitalisation data for the calibration up to November 1 2020. Here, no changepoints later than October 1 2020 were used. The fit to the regional hospitalisation data until November 1 2020, along with three-weeks ahead predictions, is provided in figures S18-S19, for the model allowing region-specific transmissibilities. The fit to the test data is provided in figures S20-S21. We note that for the future hospitalisations, we are underestimating the cases in Oslo.

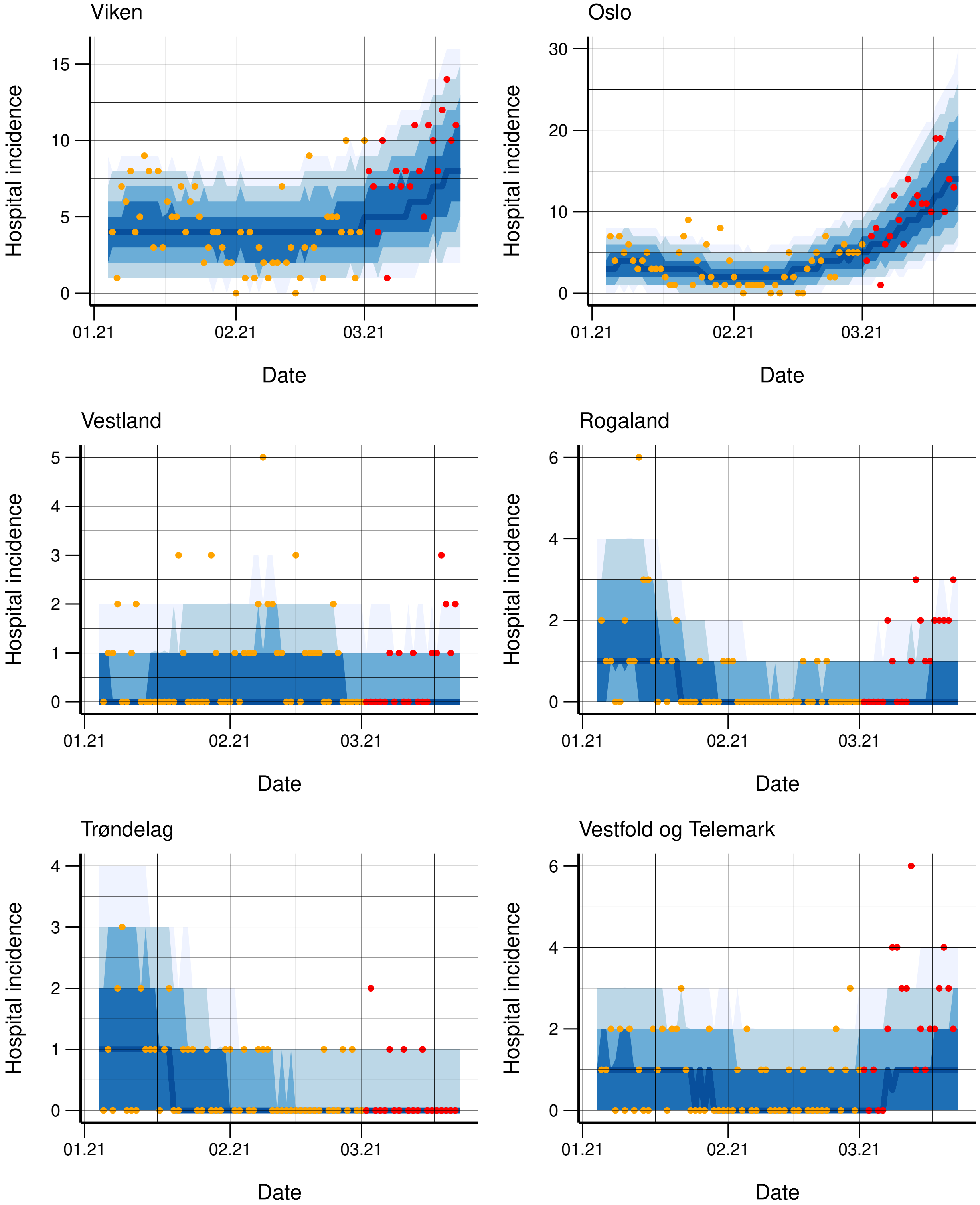


Figure S12: Observed (orange dots) and simulated hospitalisation incidence for each region, along with 95%, 90%, 75%, and 50% credibility intervals. The regions are ordered according to population, with the capital Oslo as second. The incidence for the next three weeks not used in the calibration is provided in red.

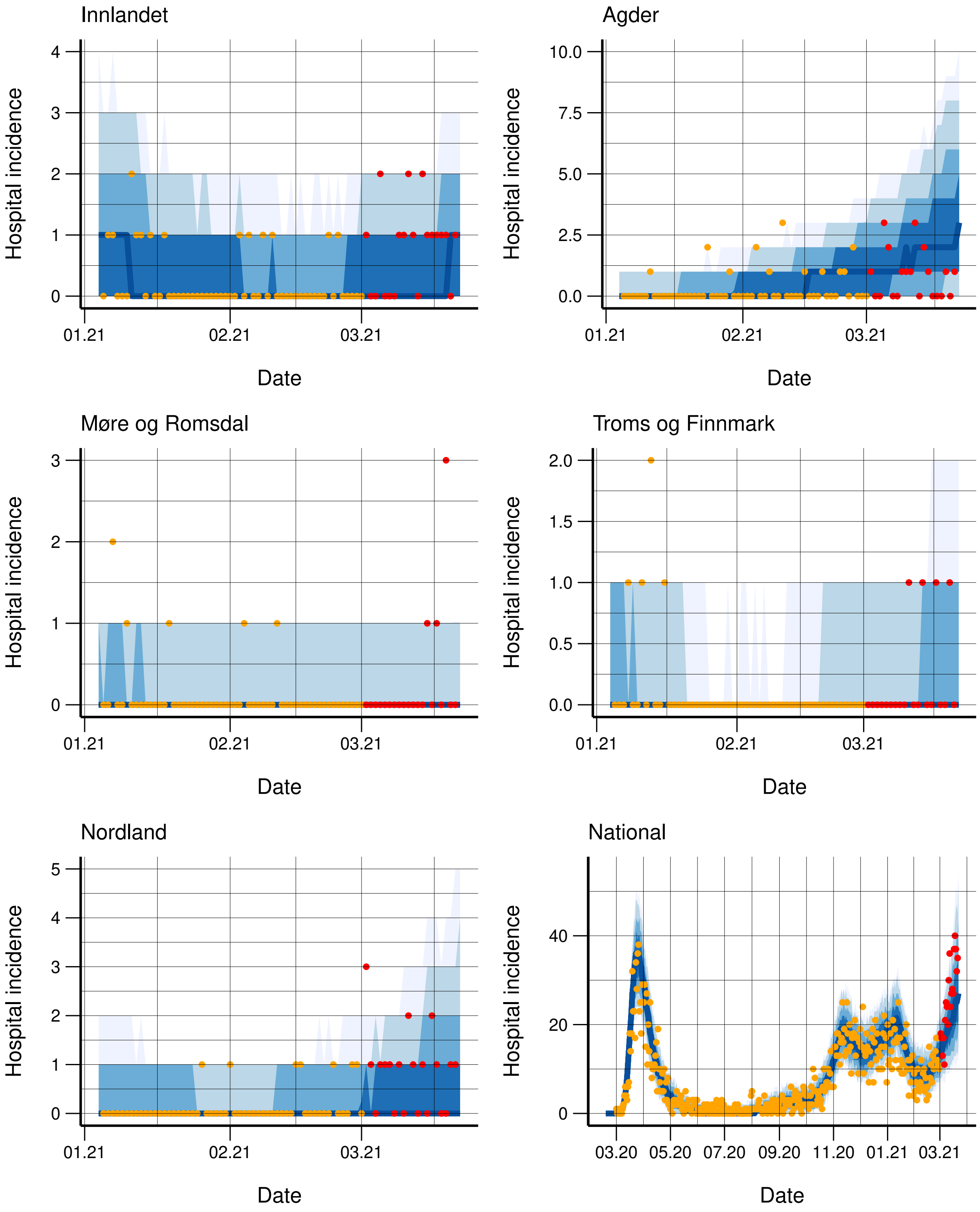


Figure S13: Observed (orange dots) and simulated hospitalisation incidence for each region, along with 95%, 90%, 75%, and 50% credibility intervals. The regions are ordered according to population. In the lower right panel, we have the national fit for the model assuming the same transmissibility in all counties. The incidence for the next three weeks not used in the calibration is provided in red.

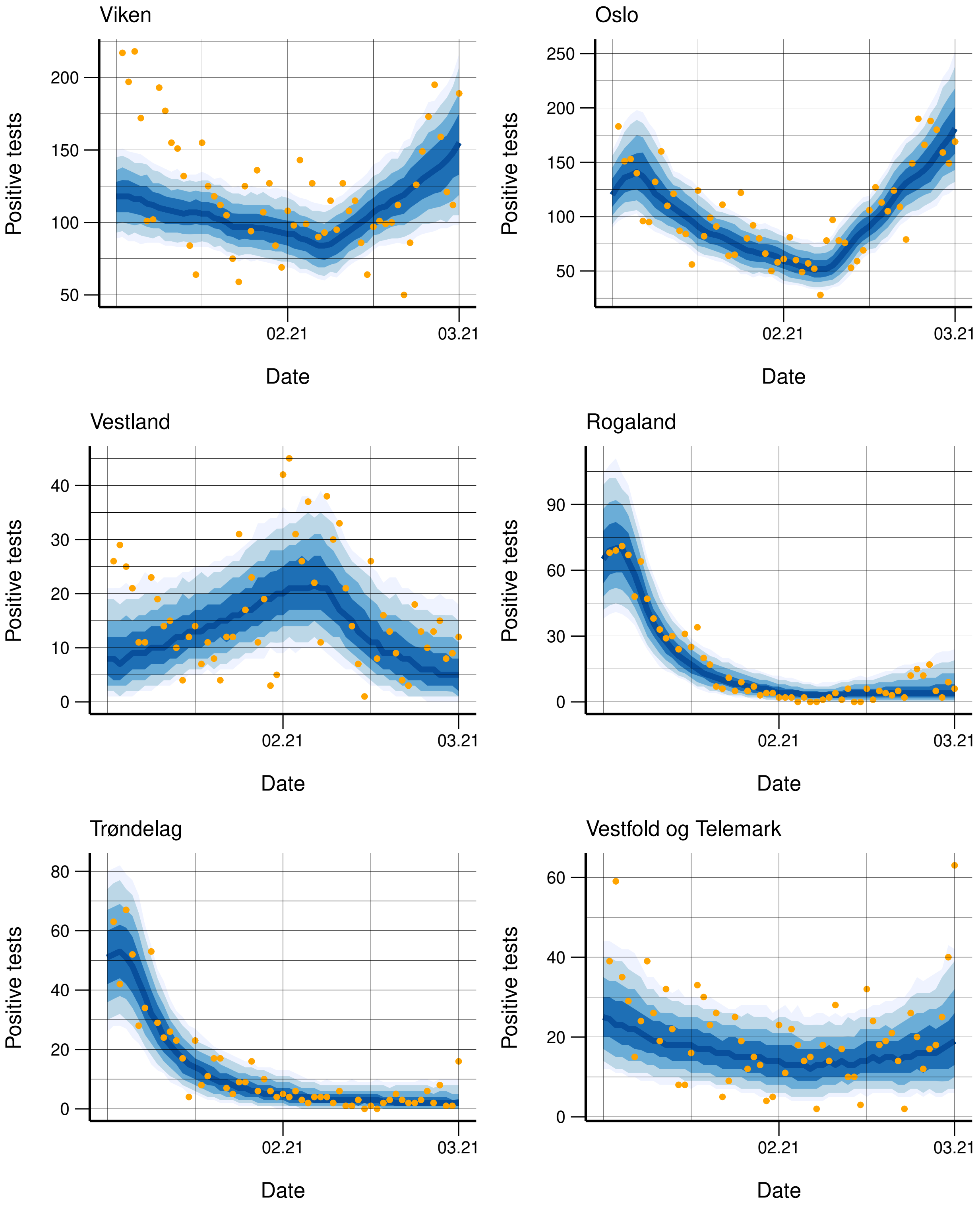


Figure S14: Observed (orange dots) and simulated positive tests for each region, along with 95%, 90%, 75%, and 50% credibility intervals. The regions are ordered according to population, with the capital Oslo as second.

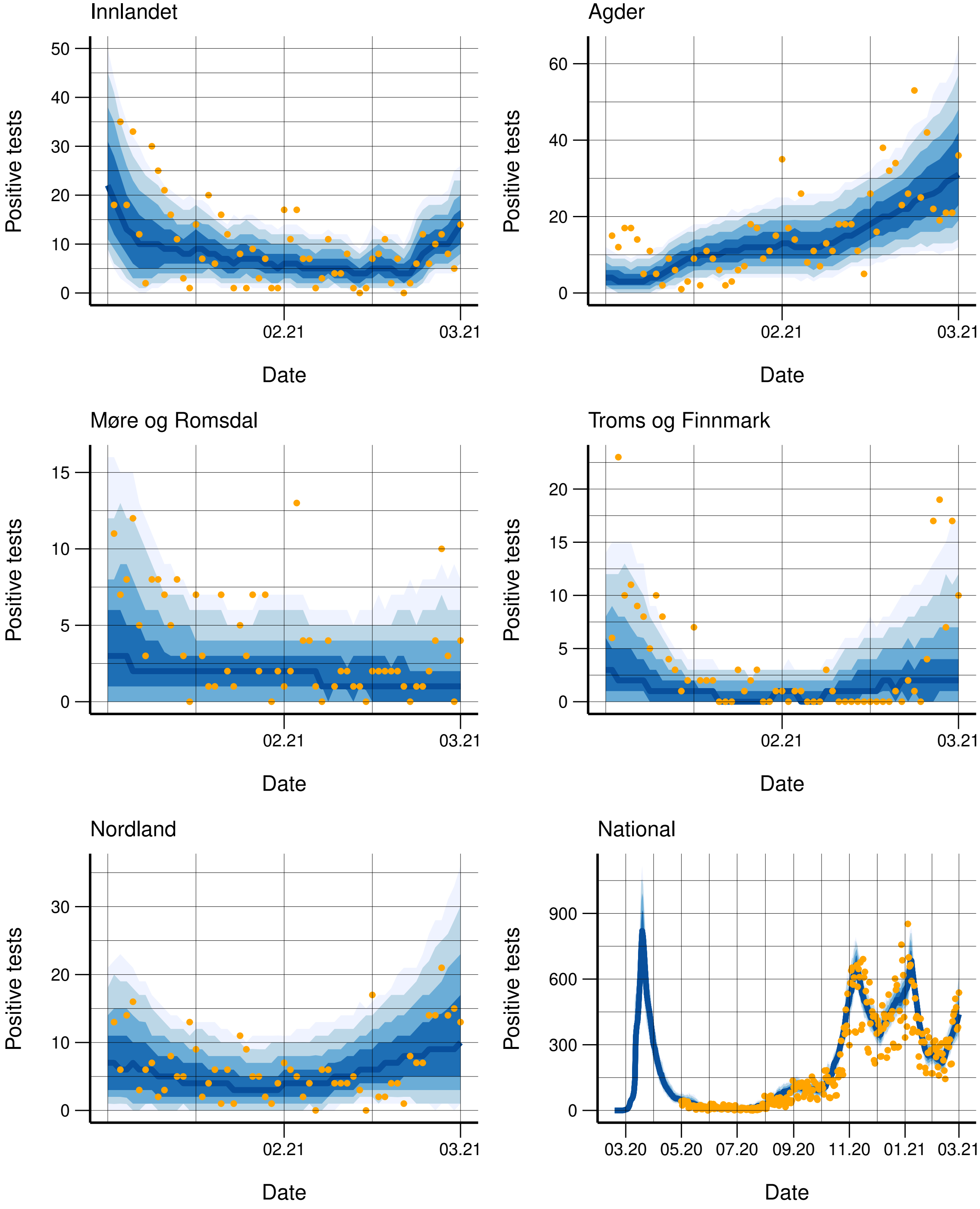


Figure S15: Observed (orange dots) and simulated positive tests for each region, along with 95%, 90%, 75%, and 50% credibility intervals. The regions are ordered according to population. In the lower right panel, we have the national fit for the model assuming the same transmissibility in all counties.

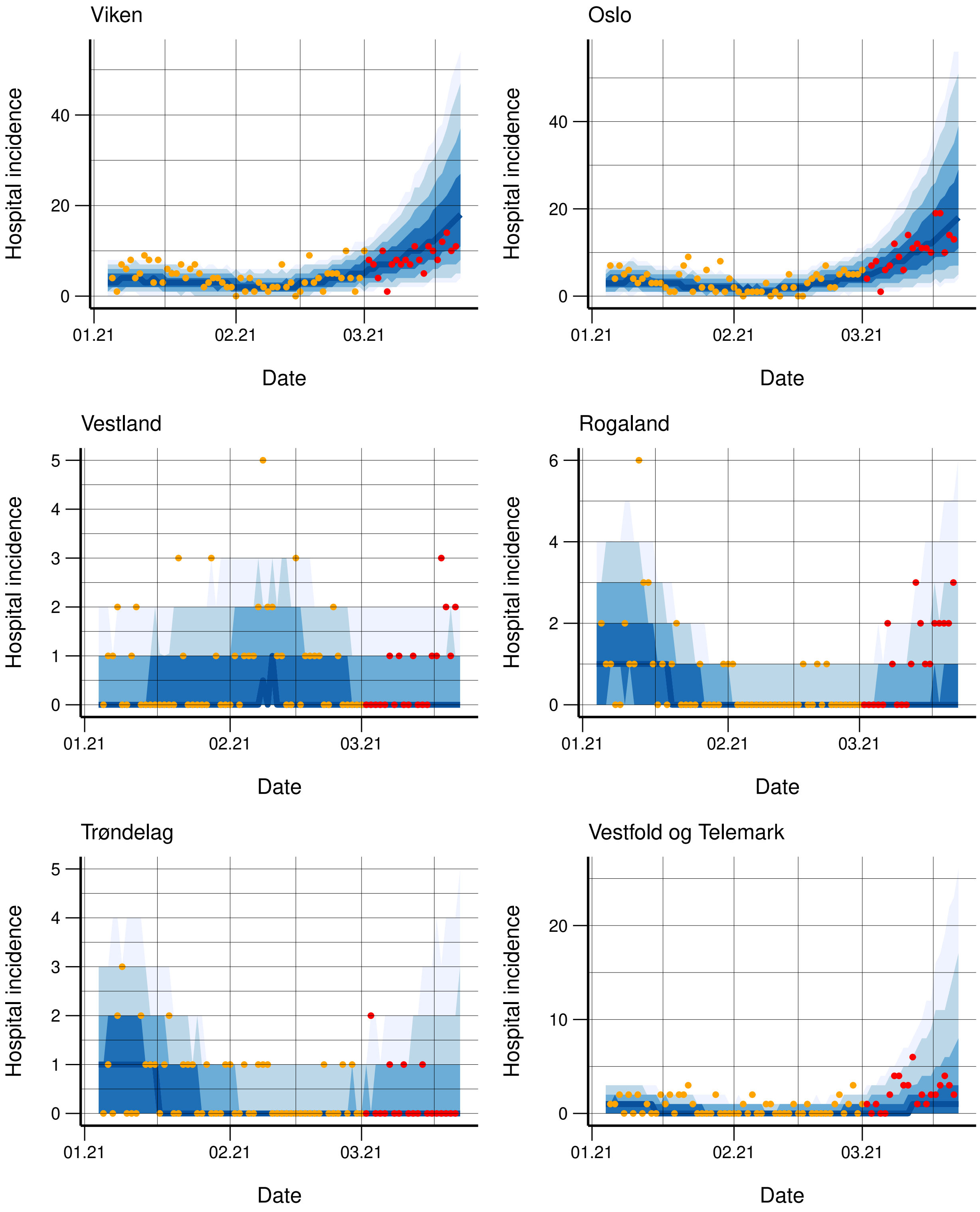


Figure S16: Observed (orange dots) and simulated hospitalisation incidence for each region, along with 95%, 90%, 75%, and 50% credibility intervals. The regions are ordered according to population, with the capital Oslo as second. The incidence for the next three weeks not used in the calibration is provided in red.

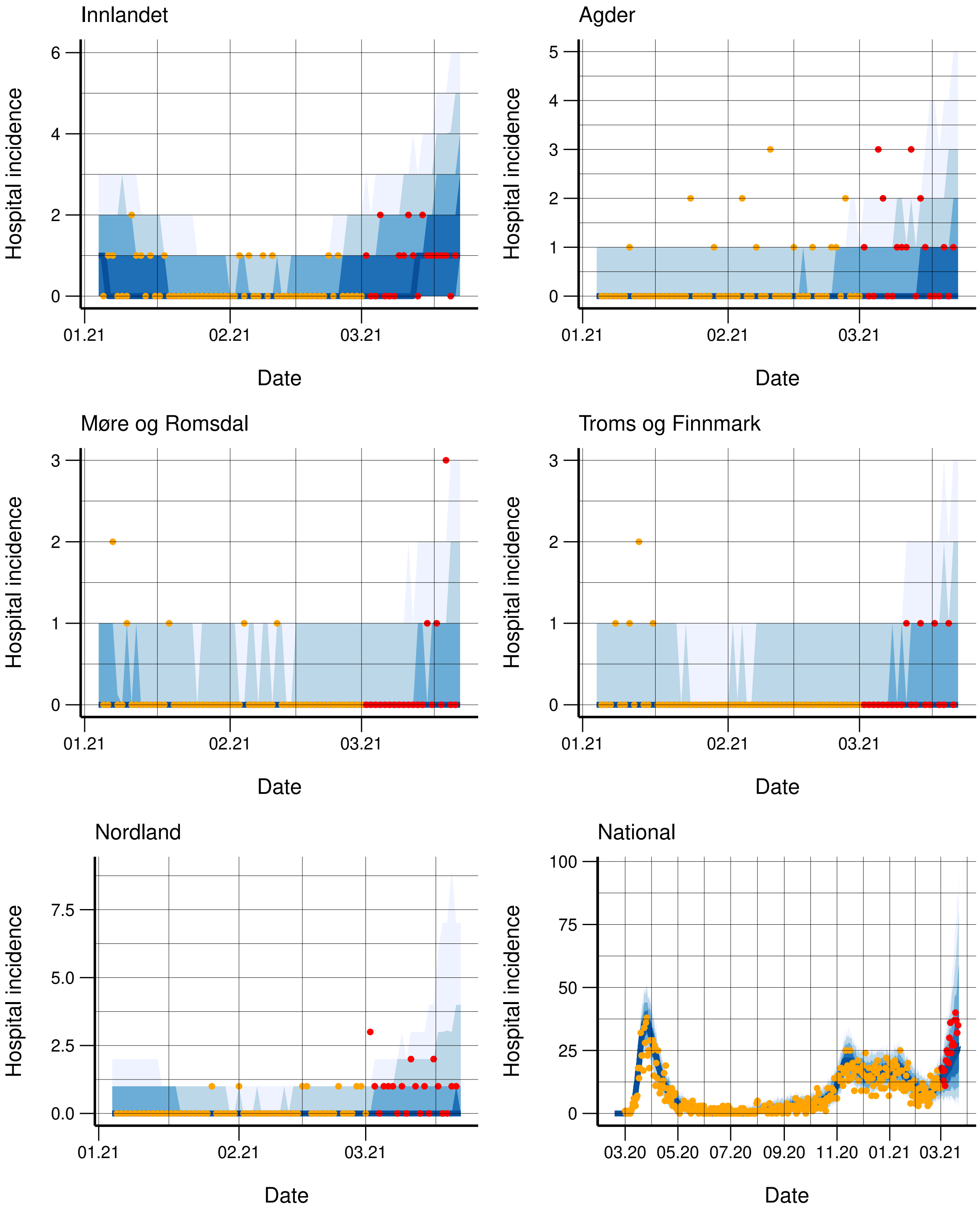


Figure S17: Observed (orange dots) and simulated hospitalisation incidence for each region, along with 95%, 90%, 75%, and 50% credibility intervals. The regions are ordered according to population. In the lower right panel, we have the national fit for the model assuming the same transmissibility in all counties. The incidence for the next three weeks not used in the calibration is provided in red.

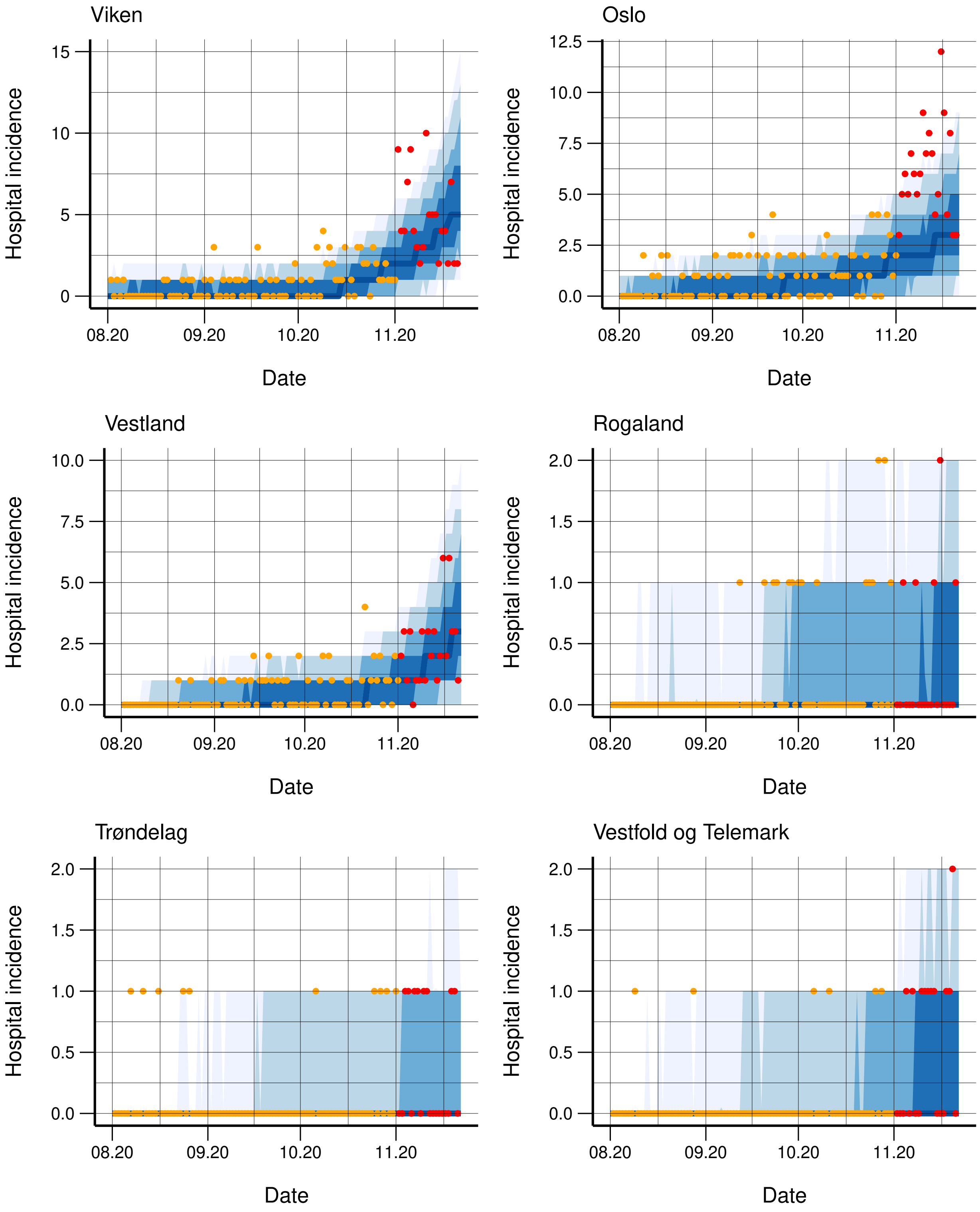


Figure S18: Observed (orange dots) and simulated hospitalisation incidence for each region, along with 95%, 90%, 75%, and 50% credibility intervals. The regions are ordered according to population, with the capital Oslo as second. The incidence for the next three weeks not used in the calibration is provided in red.

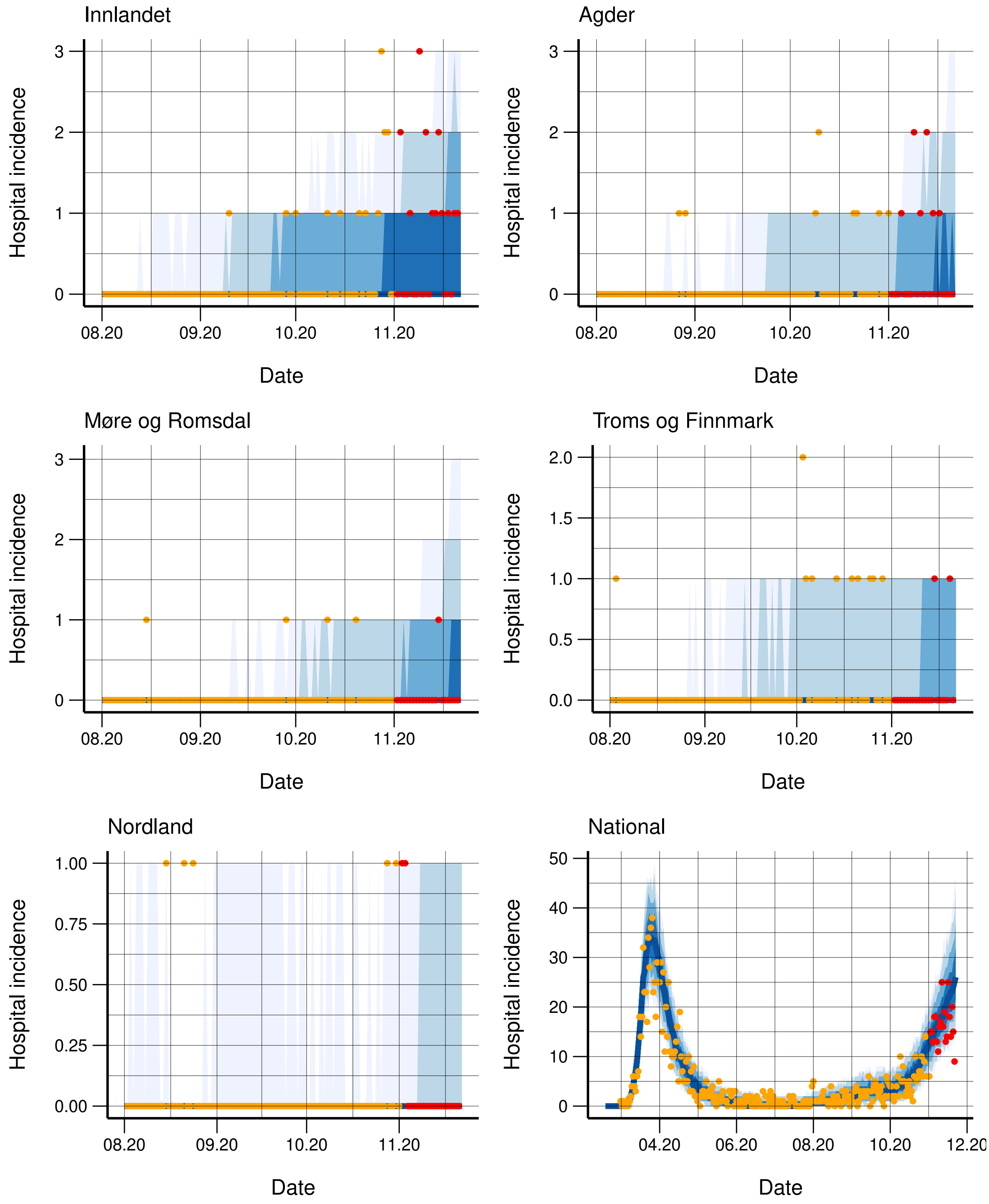


Figure S19: Observed (orange dots) and simulated hospitalisation incidence for each region, along with 95%, 90%, 75%, and 50% credibility intervals. The regions are ordered according to population. In the lower right panel, we have the national fit for the model assuming the same transmissibility in all counties. The incidence for the next three weeks not used in the calibration is provided in red.

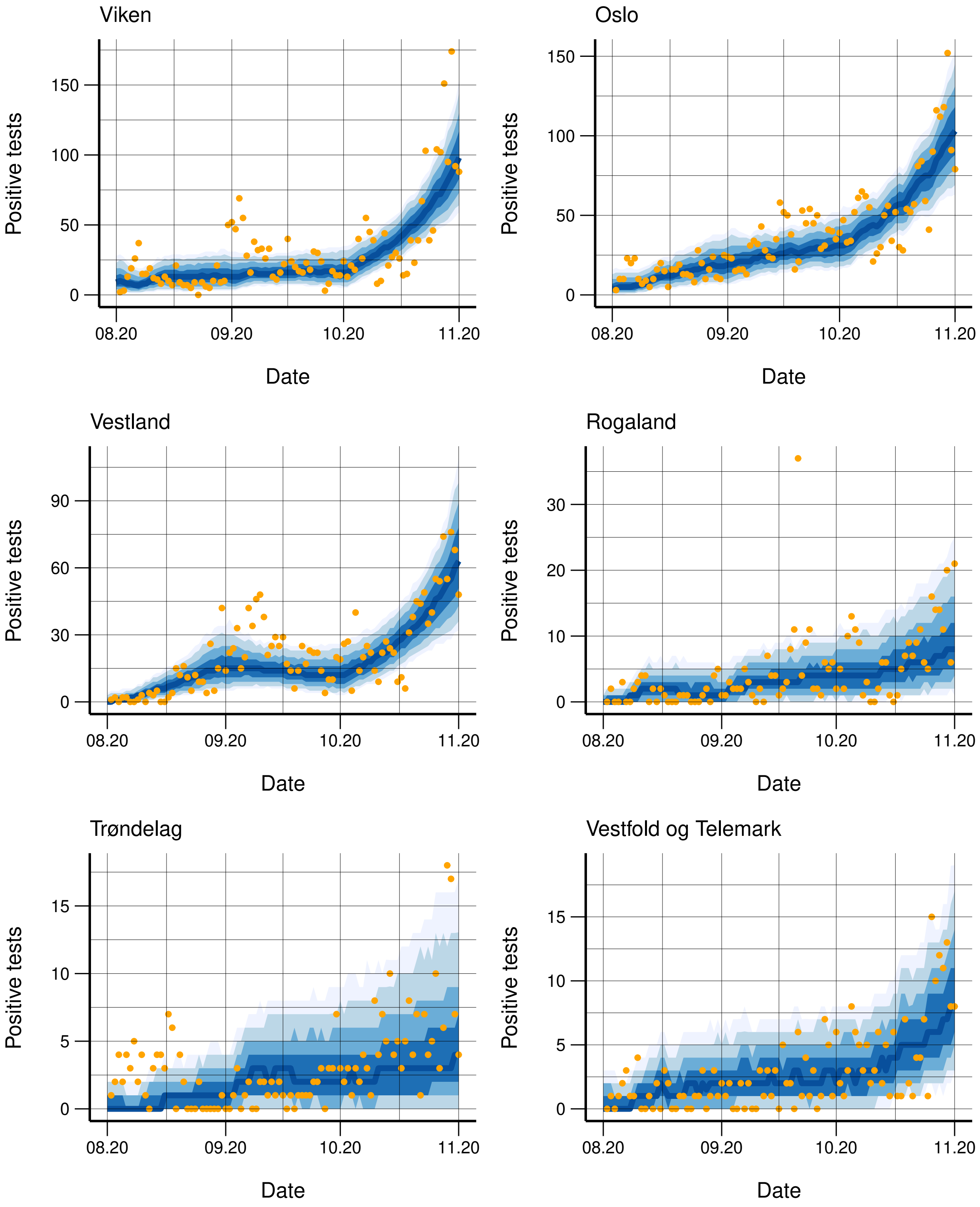


Figure S20: Observed (orange dots) and simulated positive tests for each region, along with 95%, 90%, 75%, and 50% credibility intervals. The regions are ordered according to population, with the capital Oslo as second.

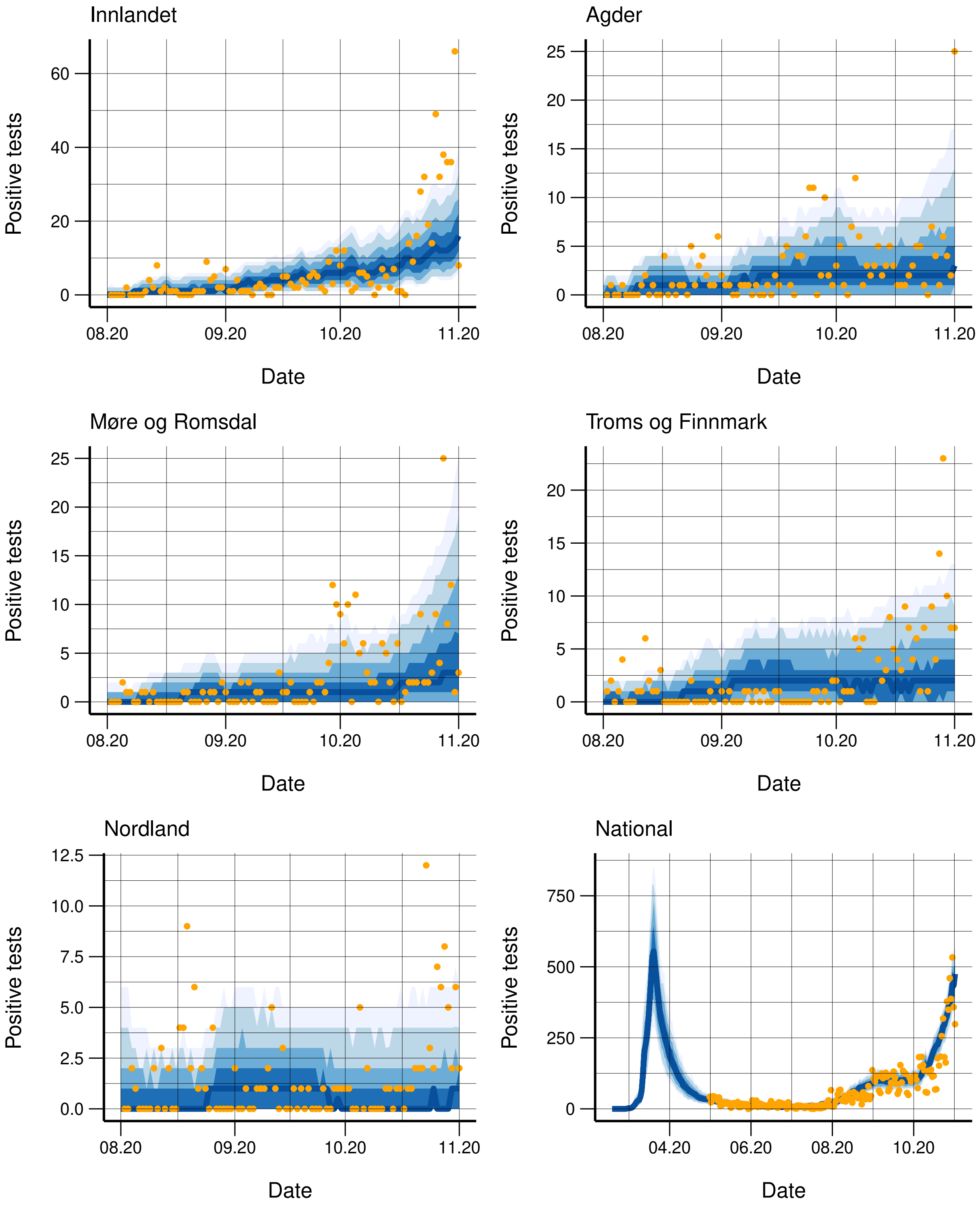


Figure S21: Observed (orange dots) and simulated positive tests for each region, along with 95%, 90%, 75%, and 50% credibility intervals. The regions are ordered according to population. In the lower right panel, we have the national fit for the model assuming the same transmissibility in all counties.

#### S3.5 Fit to data up to October 1 2020

We include also the fit to the test data and hospitalisation data for the calibration up to October 1 2020. Here, no changepoints later than September 1 2020 were used. The fit to the regional hospitalisation data until October 1 2020, along with three-weeks ahead predictions, is provided in figures S22-S23, for the model allowing region-specific transmissibilities. The fit to the test data is provided in figures S24-S25. We note the low level of hospitalisation incidence in Norway in October 2020.

#### S3.6 Fit to data up to September 1 2020

We include also the fit to the test data and hospitalisation data for the calibration up to September 1 2020. Here, no changepoints later than August 1 2020 were used. The fit to the regional hospitalisation data until September 1 2020, along with three-weeks ahead predictions, is provided in figures S26-S27, for the model allowing region-specific transmissibilities. The fit to the test data is provided in figures S28-S29. In September 2020, there were few COVID-19 cases in Norway.

#### S3.7 Fit to data up to August 1 2020

We include also the fit to the test data and hospitalisation data for the regional calibration up to August 1 2020. This is the calibration of the past which is considered fixed in the later regional calibrations. Here, no changepoints later than July 1 2020 were used. The fit to the regional hospitalisation data until August 1 2020 is provided in figures S30-S31, for the model allowing region-specific trans-

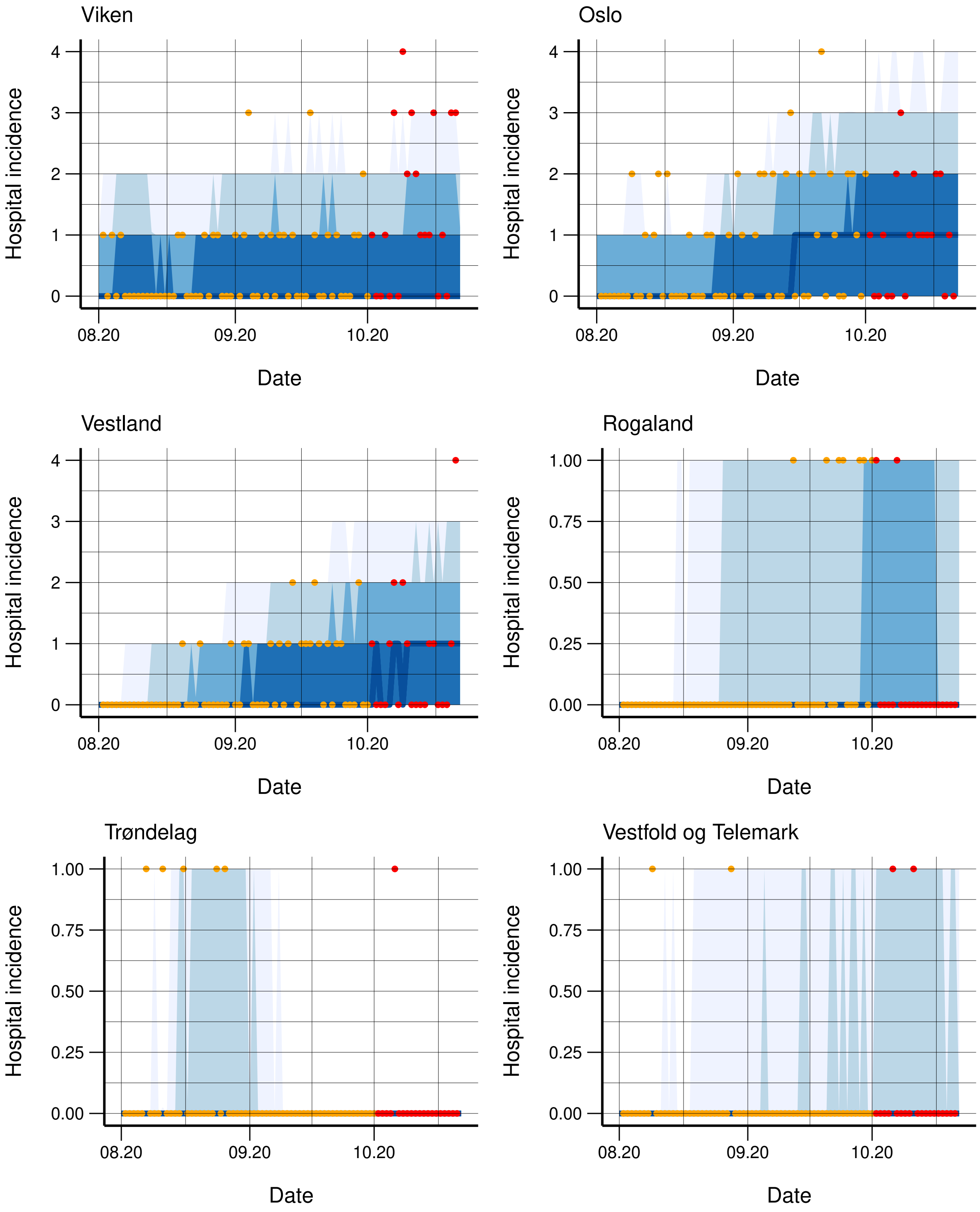


Figure S22: Observed (orange dots) and simulated hospitalisation incidence for each region, along with 95%, 90%, 75%, and 50% credibility intervals. The regions are ordered according to population, with the capital Oslo as second. The incidence for the next three weeks not used in the calibration is provided in red.

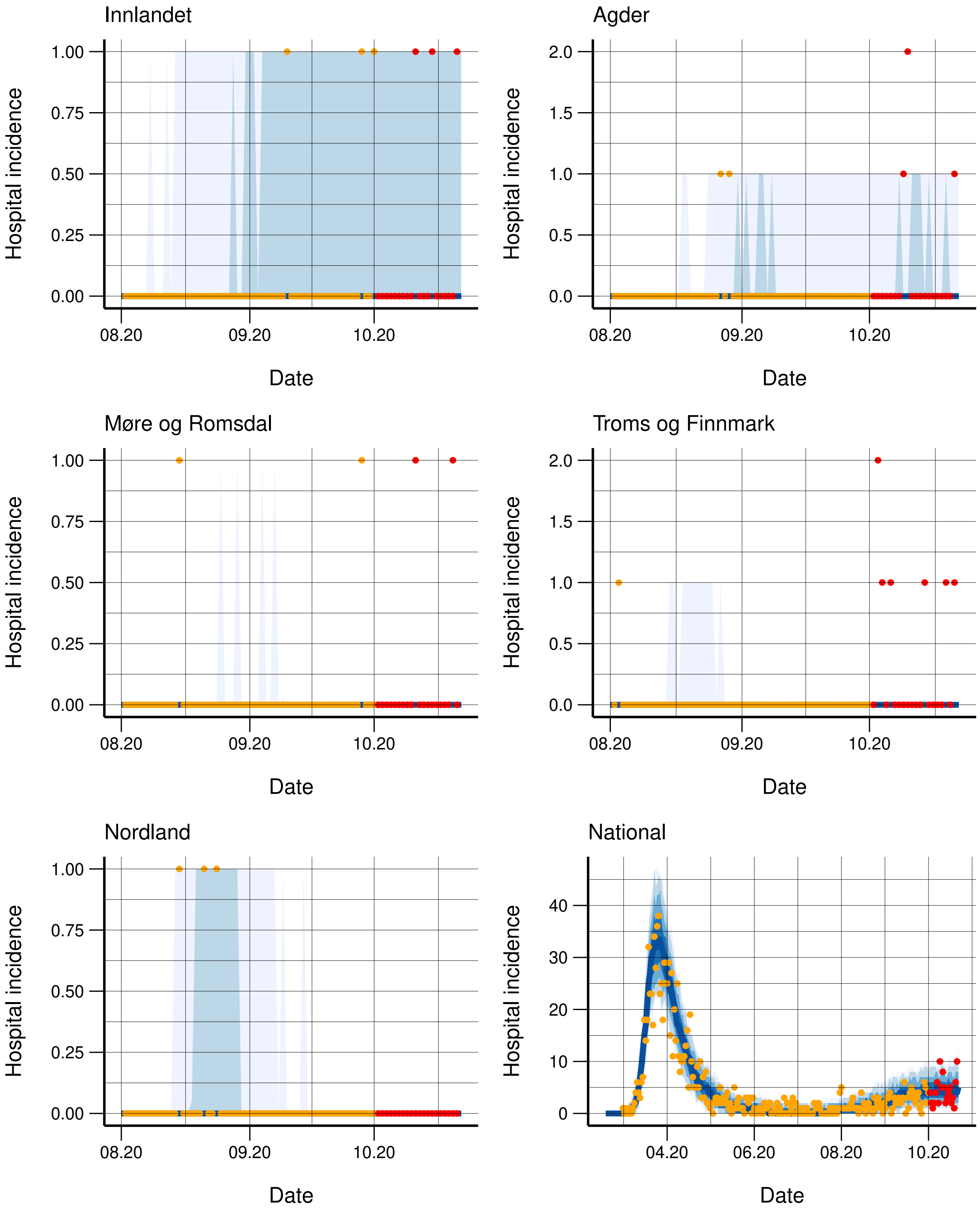


Figure S23: Observed (orange dots) and simulated hospitalisation incidence for each region, along with 95%, 90%, 75%, and 50% credibility intervals. The regions are ordered according to population. In the lower right panel, we have the national fit for the model assuming the same transmissibility in all counties. The incidence for the next three weeks not used in the calibration is provided in red.

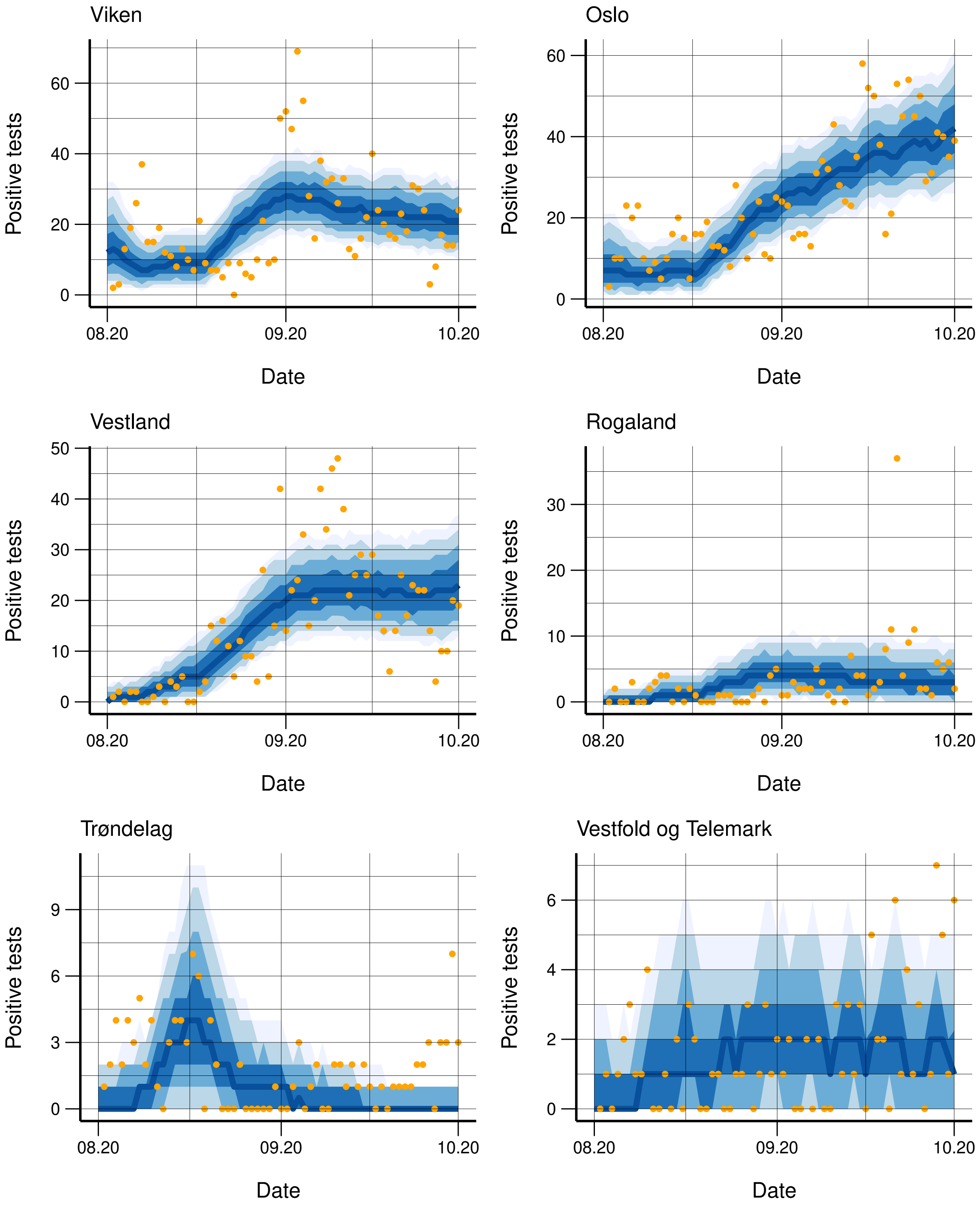


Figure S24: Observed (orange dots) and simulated positive tests for each region, along with 95%, 90%, 75%, and 50% credibility intervals. The regions are ordered according to population, with the capital Oslo as second.

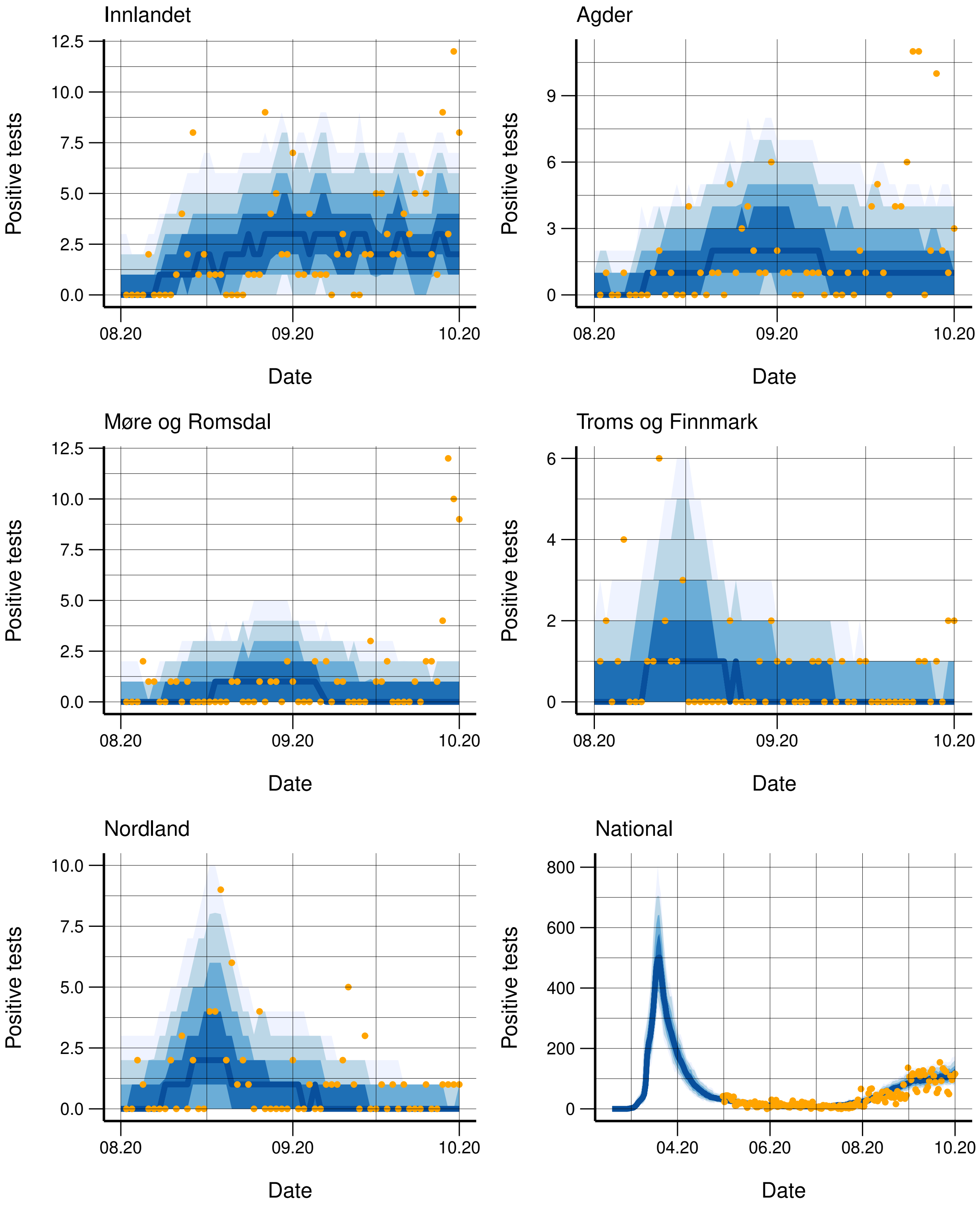


Figure S25: Observed (orange dots) and simulated positive tests for each region, along with 95%, 90%, 75%, and 50% credibility intervals. The regions are ordered according to population. In the lower right panel, we have the national fit for the model assuming the same transmissibility in all counties.

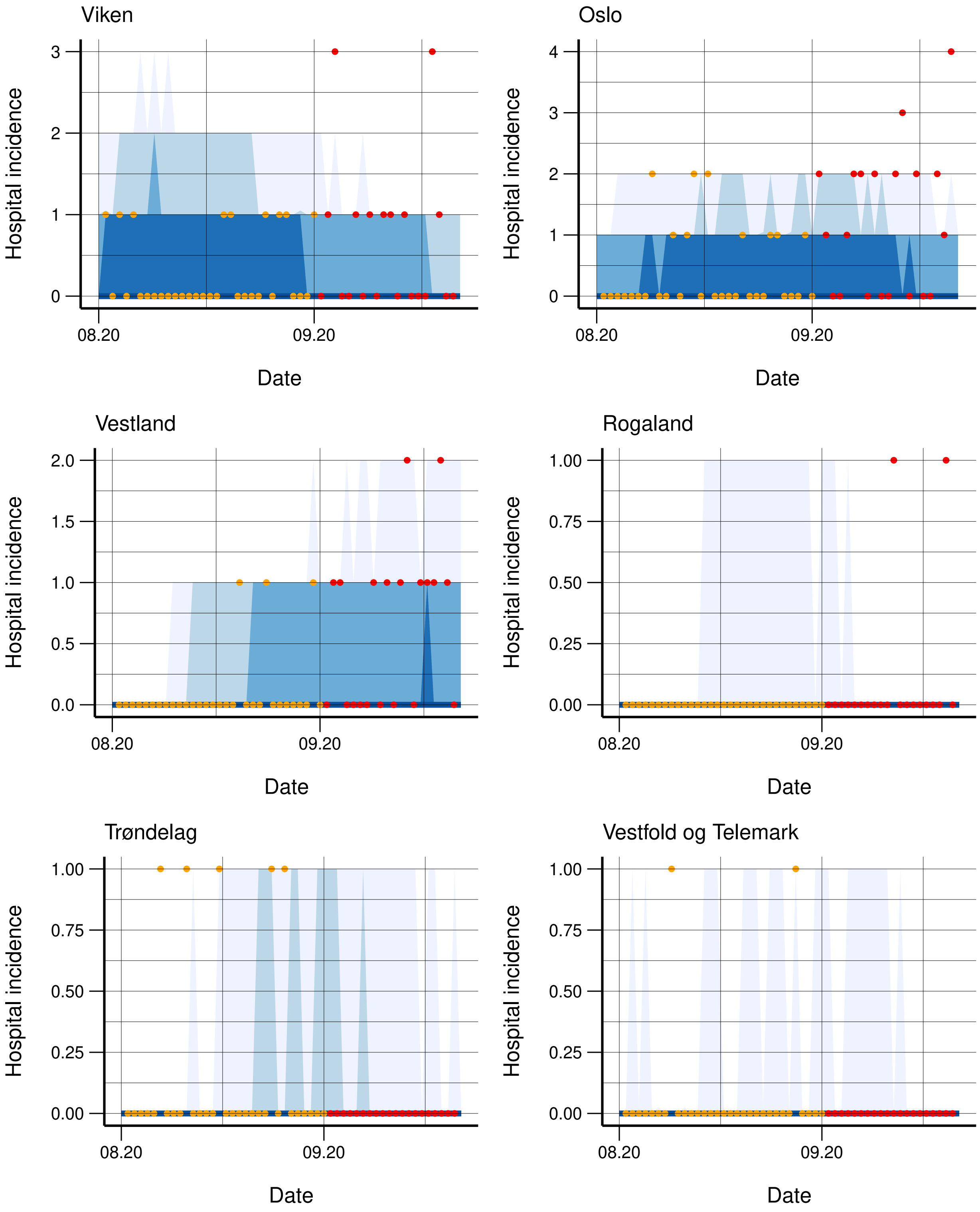


Figure S26: Observed (orange dots) and simulated hospitalisation incidence for each region, along with 95%, 90%, 75%, and 50% credibility intervals. The regions are ordered according to population, with the capital Oslo as second. The incidence for the next three weeks not used in the calibration is provided in red.

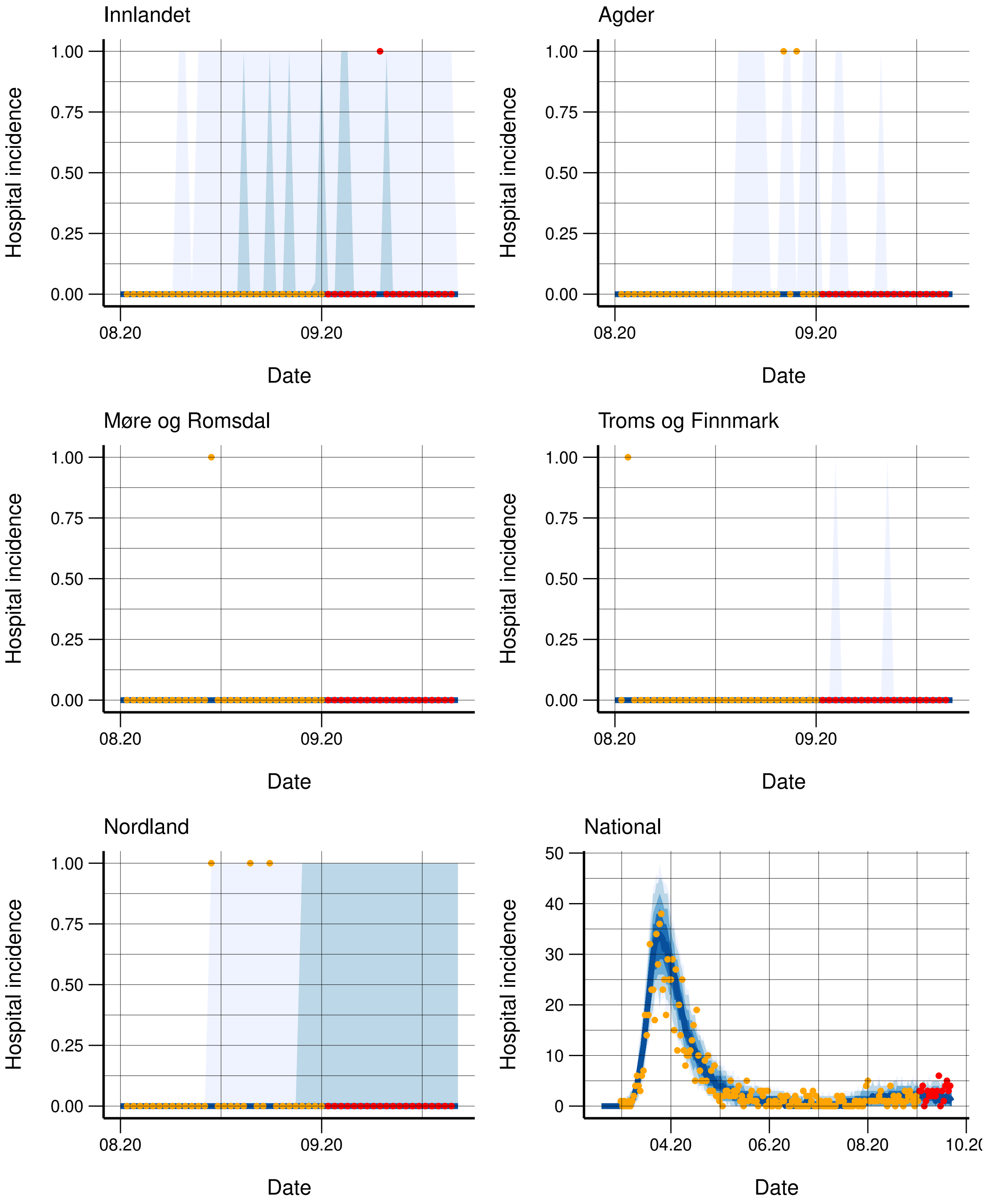


Figure S27: Observed (orange dots) and simulated hospitalisation incidence for each region, along with 95%, 90%, 75%, and 50% credibility intervals. The regions are ordered according to population. In the lower right panel, we have the national fit for the model assuming the same transmissibility in all counties. The incidence for the next three weeks not used in the calibration is provided in red.

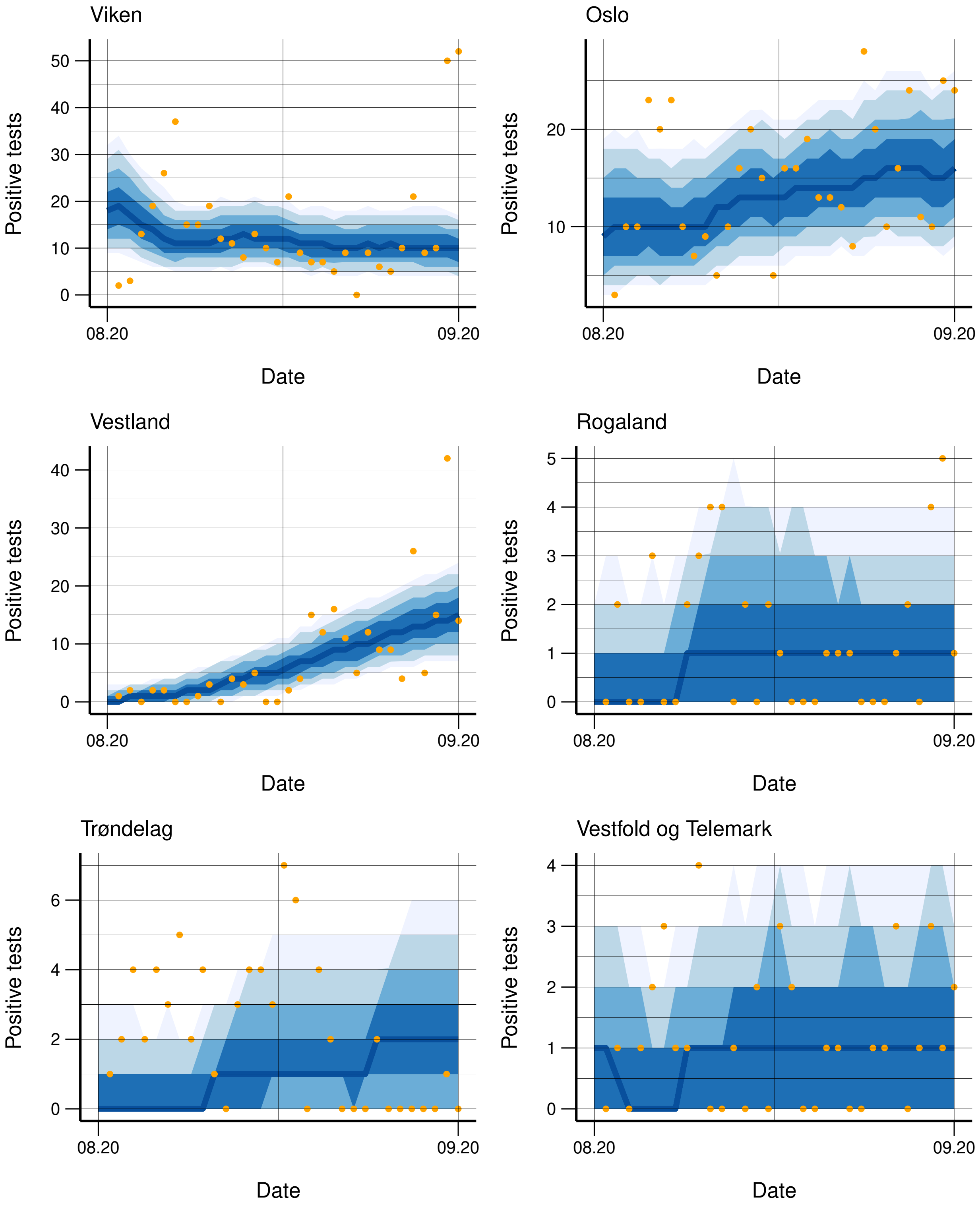


Figure S28: Observed (orange dots) and simulated positive tests for each region, along with 95%, 90%, 75%, and 50% credibility intervals. The regions are ordered according to population, with the capital Oslo as second.

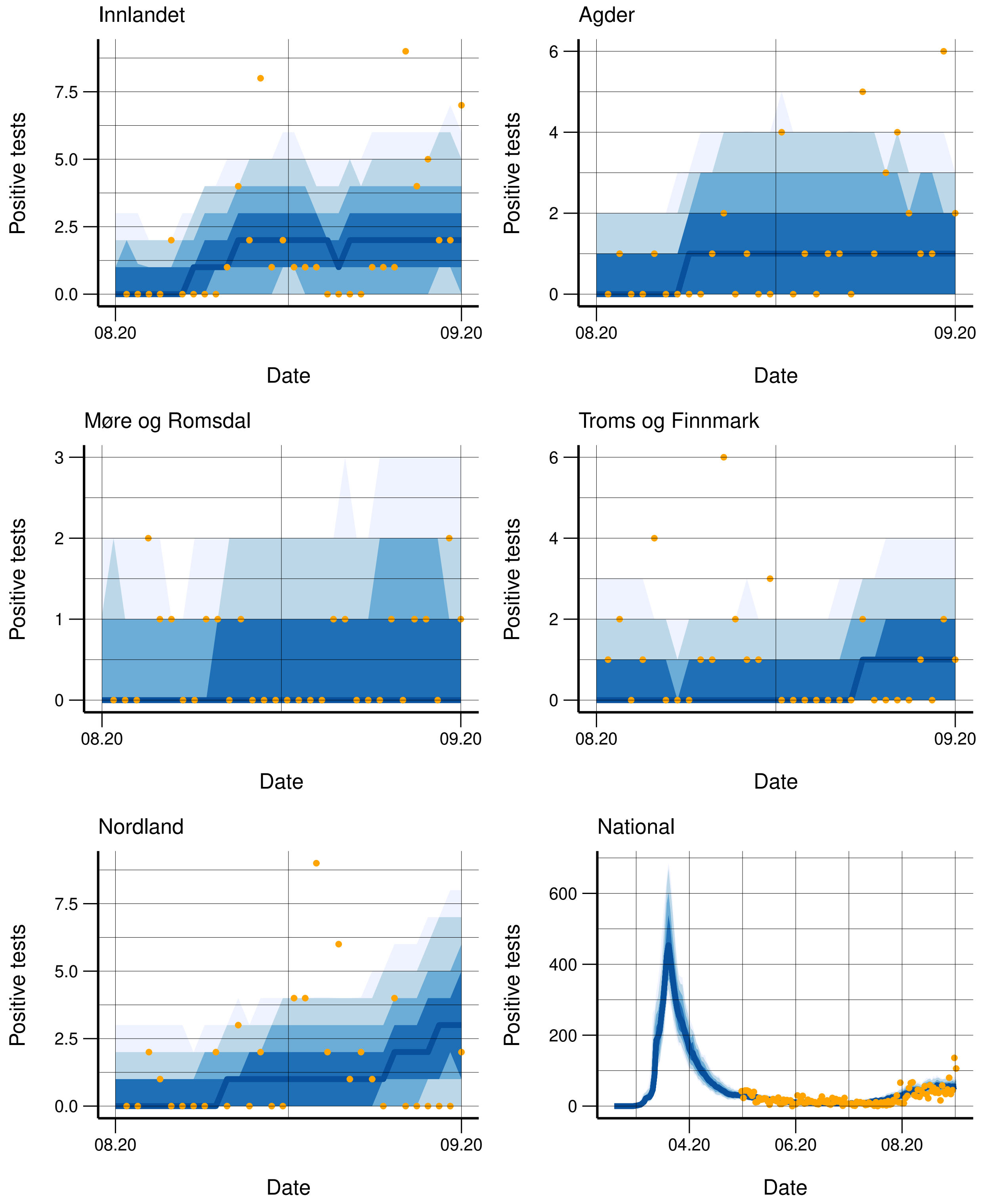


Figure S29: Observed (orange dots) and simulated positive tests for each region, along with 95%, 90%, 75%, and 50% credibility intervals. The regions are ordered according to population. In the lower right panel, we have the national fit for the model assuming the same transmissibility in all counties.

missibilities. The fit to the test data is provided in figures S32-S33. We note the low disease level during summer. The corresponding estimated reproduction numbers are provided in Figures S34-S36.

#### S3.8 Changepoint selection

We here present our calibration of the changepoint corresponding to the first lockdown in Norway. We calibrated the timing of the first changepoint by comparing the fit with a changepoint on March 11, 12, 13, 14, 15, and 16. This was done because the interventions were implemented gradually. The fit for the different changepoints is provided in figure S37. The fit was clearly best when using March 15 or March 16 as a changepoint. The differences between the two were minimal, but March 15 gave a slightly better fit, and we thus chose to use March 15 as the changepoint.

#### S3.9 Computational time and convergence

In particular for the regional calibration, the parameter space is very high-dimensional and correlated in both time and space, which makes the calibration exercise difficult. The calibration took one week running on 1400 cores.

Figure S38 shows the evolution of the estimated distributions of the parameters throughout the calibration rounds.

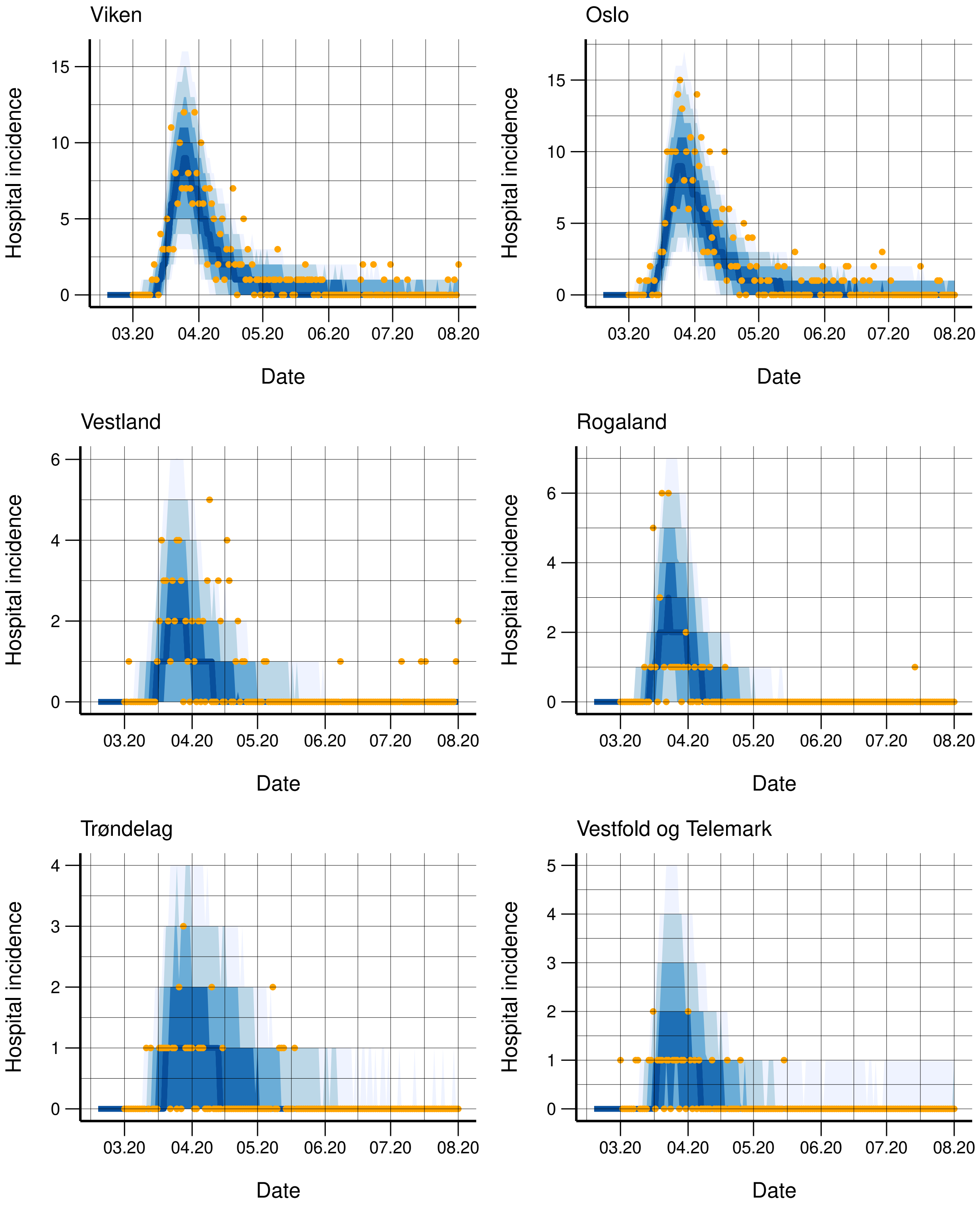


Figure S30: Observed (orange dots) and simulated hospitalisation incidence for each region, along with 95%, 90%, 75%, and 50% credibility intervals. The regions are ordered according to population.

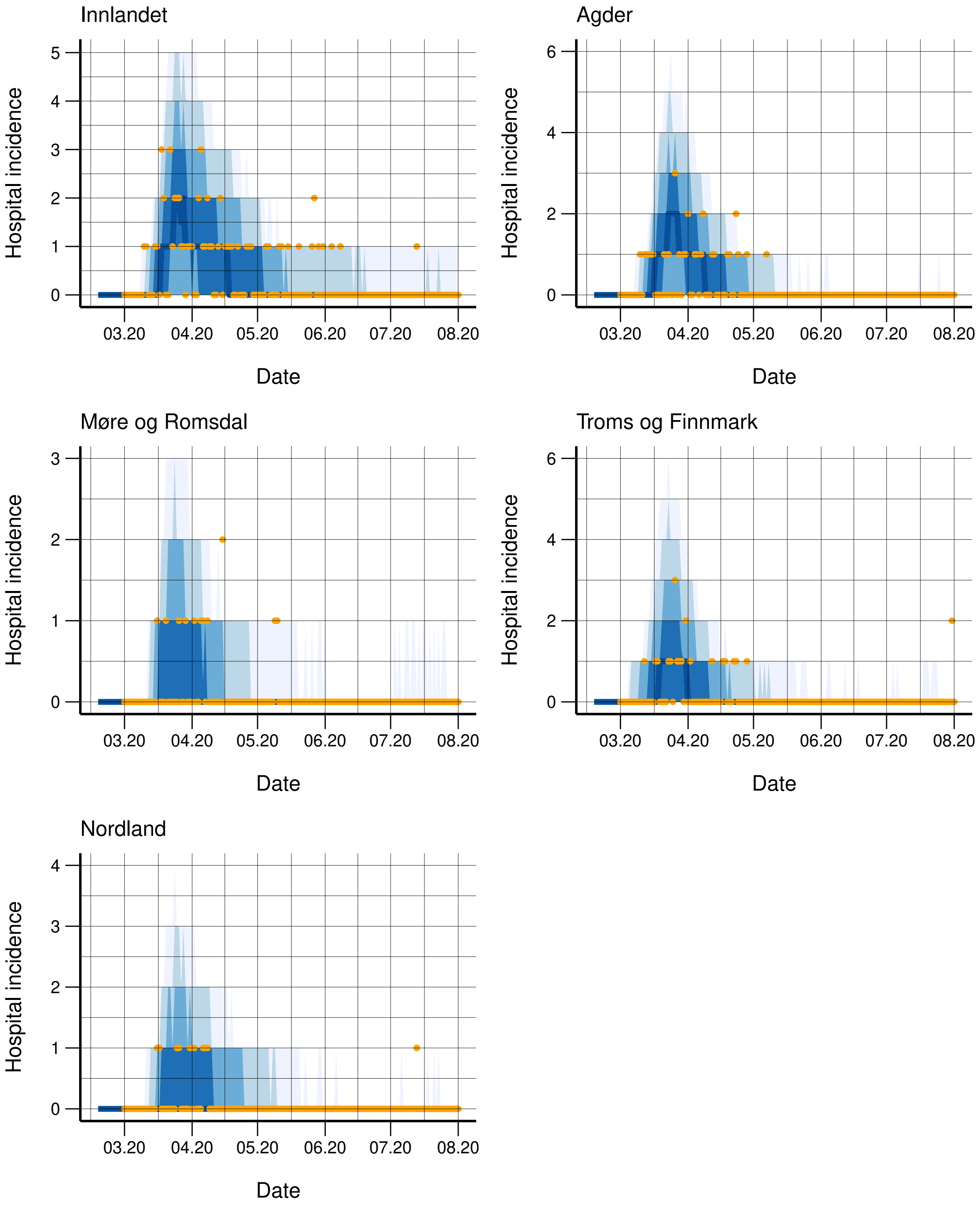


Figure S31: Observed (orange dots) and simulated hospitalisation incidence for each region, along with 95%, 90%, 75%, and 50% credibility intervals. The regions are ordered according to population.

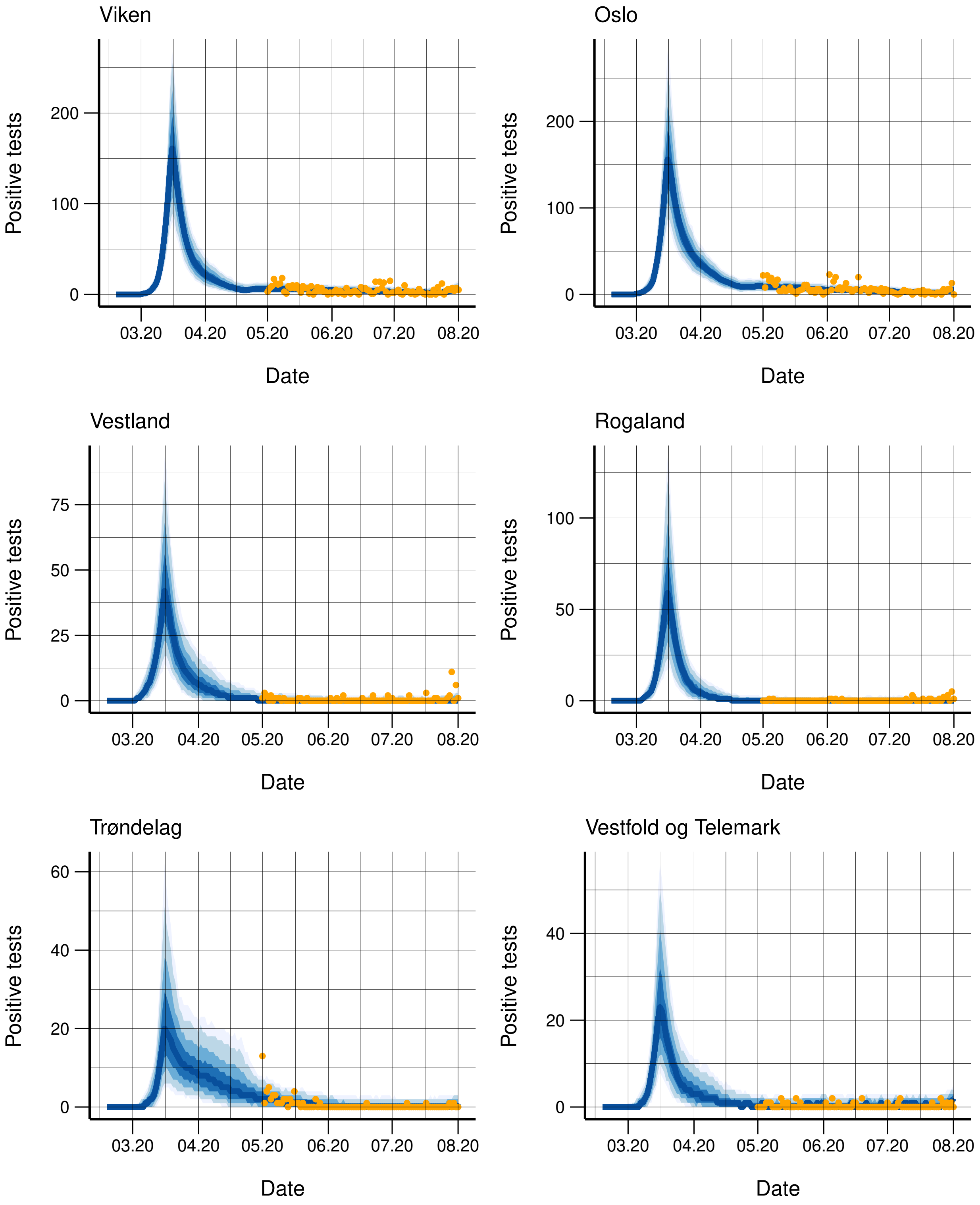


Figure S32: Observed (orange dots) and simulated positive tests for each region, along with 95%, 90%, 75%, and 50% credibility intervals. The regions are ordered according to population.

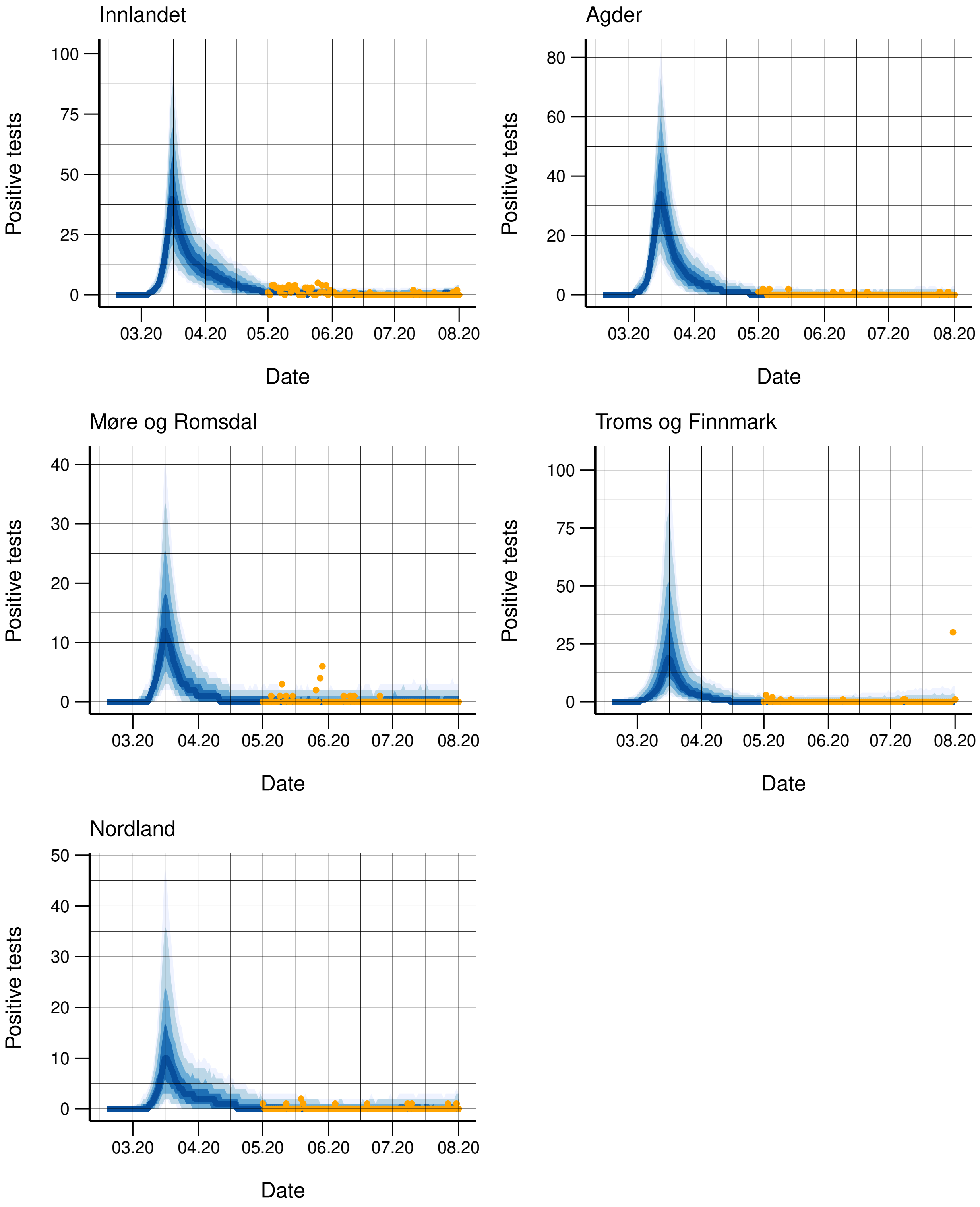


Figure S33: Observed (orange dots) and simulated positive tests for each region, along with 95%, 90%, 75%, and 50% credibility intervals. The regions are ordered according to population.

| Calibration | **Sep 1st-2020** | **Oct 1st-2020** | |
|---|---|---|---|
| Period | 08/01-08/31 | 08/01-08/16 | 08/17-09/30 |
| Oslo | 0.88 ( 0.73 - 1.03 ) | 0.58 ( 0.22 - 0.99 ) | 1.28 ( 1.16 - 1.39 ) |
| Rogaland | 0.19 ( 0.04 - 0.34 ) | 0.15 ( 0.01 - 0.38 ) | 0.7 ( 0.37 - 0.97 ) |
| Nordland | 0.16 ( 0.02 - 0.37 ) | 0.28 ( 0.03 - 0.67 ) | 0.29 ( 0.03 - 0.66 ) |
| Møre og Romsdal | 0.84 ( 0.42 - 1.23 ) | 1.4 ( 0.31 - 2.5 ) | 0.21 ( 0.01 - 0.5 ) |
| Viken | 0.39 ( 0.29 - 0.49 ) | 0.32 ( 0.09 - 0.53 ) | 0.88 ( 0.77 - 0.99 ) |
| Innlandet | 0.42 ( 0.17 - 0.64 ) | 0.36 ( 0.03 - 0.82 ) | 0.48 ( 0.1 - 0.82 ) |
| Vestfold og Telemark | 0.15 ( 0.01 - 0.37 ) | 0.32 ( 0.02 - 0.82 ) | 0.12 ( 0.01 - 0.35 ) |
| Agder | 0.27 ( 0.08 - 0.46 ) | 0.18 ( 0.01 - 0.46 ) | 0.55 ( 0.11 - 0.93 ) |
| Vestland | 0.95 ( 0.79 - 1.12 ) | 0.82 ( 0.29 - 1.28 ) | 1.1 ( 0.99 - 1.21 ) |
| Trøndelag | 0.36 ( 0.14 - 0.58 ) | 0.79 ( 0.21 - 1.43 ) | 0.11 ( 0.01 - 0.25 ) |
| Troms og Finmark | 0.21 ( 0.03 - 0.44 ) | 0.56 ( 0.07 - 1.15 ) | 0.09 ( 0.01 - 0.21 ) |
| Calibration | **Nov 1st-2020** | | |
| Period | 08/01-08/31 | 09/01-09/30 | 10/01-10/31 |
| Oslo | 0.97 ( 0.62 - 1.32 ) | 1.25 ( 0.97 - 1.52 ) | 1.46 ( 1.22 - 1.68 ) |
| Rogaland | 0.25 ( 0.02 - 0.51 ) | 1.03 ( 0.7 - 1.29 ) | |
| Nordland | 0.24 ( 0.02 - 0.61 ) | 0.75 ( 0.06 - 1.65 ) | 1.18 ( 0.15 - 2.08 ) |
| Møre og Romsdal | 0.4 ( 0.04 - 0.91 ) | 0.63 ( 0.1 - 1.17 ) | |
| Viken | 0.46 ( 0.11 - 0.74 ) | 0.88 ( 0.51 - 1.23 ) | 1.49 ( 1.18 - 1.83 ) |
| Innlandet | 0.23 ( 0.01 - 0.71 ) | 1.14 ( 0.79 - 1.46 ) | |
| Vestfold og Telemark | 0.25 ( 0.02 - 0.7 ) | 0.62 ( 0.14 - 1.13 ) | |
| Agder | 0.16 ( 0.01 - 0.45 ) | 1.04 ( 0.35 - 1.69 ) | 0.82 ( 0.11 - 1.68 ) |
| Vestland | 0.92 ( 0.6 - 1.27 ) | 0.97 ( 0.65 - 1.25 ) | 1.51 ( 1.18 - 1.88 ) |
| Trøndelag | 0.19 ( 0.02 - 0.43 ) | 0.93 ( 0.52 - 1.24 ) | |
| Troms og Finmark | 0.25 ( 0.02 - 0.58 ) | 0.87 ( 0.45 - 1.22 ) | |

Figure S34: Estimated regional reproductive numbers. Calibration from August 1st (2020). Median and 95 % CIs.

| Calibration | **March 1st-2021 (test and hospitalizations)** | | **March 1st-2021 (hospitalizations)** | |
|---|---|---|---|---|
| Period | 01/04-02/04 | 02/05-03/01 | 01/04-02/04 | 02/05-03/01 |
| Oslo | 0.78 ( 0.69 - 0.88 ) | 1.67 ( 1.44 - 1.91 ) | 0.71 ( 0.54 - 0.87 ) | 1.77 ( 1.22 - 2.33 ) |
| Rogaland | 0.3 ( 0.16 - 0.43 ) | 0.85 ( 0.13 - 1.65 ) | 0.14 ( 0.01 - 0.38 ) | 0.84 ( 0.06 - 2.15 ) |
| Nordland | 0.42 ( 0.12 - 0.68 ) | 0.73 ( 0.1 - 1.51 ) | 0.29 ( 0.02 - 0.76 ) | 0.78 ( 0.04 - 2 ) |
| Møre og Romsdal | 0.69 ( 0.35 - 1.01 ) | 1.3 ( 0.53 - 1.96 ) | 0.32 ( 0.02 - 0.85 ) | 0.88 ( 0.09 - 2.29 ) |
| Viken | 0.84 ( 0.76 - 0.92 ) | 1.22 ( 1 - 1.41 ) | 0.71 ( 0.55 - 0.86 ) | 1.57 ( 1.1 - 2.05 ) |
| Innlandet | 0.44 ( 0.18 - 0.66 ) | 0.4 ( 0.04 - 0.96 ) | 0.19 ( 0.01 - 0.55 ) | 0.65 ( 0.05 - 1.64 ) |
| Vestfold og Telemark | 0.73 ( 0.51 - 0.91 ) | 1.01 ( 0.57 - 1.46 ) | 0.32 ( 0.02 - 0.72 ) | 1.17 ( 0.12 - 2.5 ) |
| Agder | 0.88 ( 0.61 - 1.1 ) | 1.39 ( 0.96 - 1.85 ) | 0.31 ( 0.03 - 0.78 ) | 0.7 ( 0.05 - 1.86 ) |
| Vestland | 0.94 ( 0.78 - 1.11 ) | 0.5 ( 0.12 - 0.95 ) | 1.02 ( 0.6 - 1.38 ) | 0.39 ( 0.03 - 1.03 ) |
| Trøndelag | 0.33 ( 0.16 - 0.51 ) | 0.58 ( 0.09 - 1.17 ) | 0.2 ( 0.02 - 0.52 ) | 0.75 ( 0.04 - 1.92 ) |
| Troms og Finmark | 0.14 ( 0.01 - 0.37 ) | 0.77 ( 0.09 - 1.71 ) | 0.27 ( 0.02 - 0.76 ) | 0.73 ( 0.05 - 1.9 ) |

Figure S35: Estimated regional reproductive numbers. Calibration from January 4th (2021) up to March 1st. Median and 95 % CIs.

| | April 1st-2021 | | | |
|---|---|---|---|---|
| | $R_1$ | Period | $R_2$ | Period |
| Oslo | 0.88 ( 0.81 - 0.95 ) | 01/04-02/04 | 1.56 ( 1.49 - 1.64 ) | 02/05-03/13 |
| Rogaland | 0.25 ( 0.1 - 0.39 ) | 01/04-01/31 | 1.29 ( 0.63 - 1.8 ) | 02/01-02/19 |
| Nordland | 0.36 ( 0.13 - 0.59 ) | 01/04-02/04 | 0.36 ( 0.03 - 0.87 ) | 02/05-03/08 |
| Møre og Romsdal | 0.52 ( 0.18 - 0.87 ) | 01/04-02/04 | 1.16 ( 0.26 - 2.09 ) | 02/05-02/19 |
| Viken | 0.86 ( 0.78 - 0.94 ) | 01/04-02/04 | 1.13 ( 0.93 - 1.34 ) | 02/05-02/21 |
| Innlandet | 0.23 ( 0.03 - 0.48 ) | 01/04-02/21 | 0.81 ( 0.29 - 1.38 ) | 02/22-03/08 |
| Vestfold og Telemark | 0.57 ( 0.32 - 0.83 ) | 01/04-02/04 | 1.6 ( 1 - 2.22 ) | 02/05-02/21 |
| Agder | 0.67 ( 0.45 - 0.91 ) | 01/04-02/19 | 0.85 ( 0.23 - 1.45 ) | 02/20-03/08 |
| Vestland | 0.65 ( 0.34 - 0.98 ) | 01/04-02/04 | 0.83 ( 0.41 - 1.21 ) | 02/05-03/08 |
| Trøndelag | 0.45 ( 0.23 - 0.65 ) | 01/04-02/04 | 0.2 ( 0.01 - 0.59 ) | 02/05-03/08 |
| Troms og Finmark | 0.33 ( 0.09 - 0.57 ) | 01/04-02/04 | 0.55 ( 0.04 - 1.5 ) | 02/05-02/19 |
| | $R_3$ | Period | $R_4$ | Period |
| Oslo | 0.84 ( 0.72 - 0.99 ) | 03/14-04/01 | - | |
| Rogaland | 1.84 ( 1.19 - 2.7 ) | 02/20-03/08 | 1.18 ( 0.79 - 1.56 ) | 03/09-04/01 |
| Nordland | 1.72 ( 0.49 - 3.02 ) | 03/09-04/01 | - | |
| Møre og Romsdal | 1.22 ( 0.51 - 1.89 ) | 02/20-03/08 | 0.65 ( 0.26 - 1.02 ) | 03/09-04/01 |
| Viken | 1.61 ( 1.43 - 1.79 ) | 02/22-03/08 | 1.19 ( 1.08 - 1.29 ) | 03/09-04/01 |
| Innlandet | 0.57 ( 0.16 - 1.01 ) | 03/09-04/01 | - | |
| Vestfold og Telemark | 1.73 ( 1.26 - 2.27 ) | 02/22-03/08 | 0.55 ( 0.26 - 0.85 ) | 03/09-04/01 |
| Agder | 0.29 ( 0.03 - 0.84 ) | 03/09-04/01 | - | |
| Vestland | 0.94 ( 0.28 - 1.68 ) | 03/09-04/01 | - | |
| Trøndelag | 0.53 ( 0.05 - 1.34 ) | 03/09-04/01 | - | |
| Troms og Finmark | 1.07 ( 0.25 - 1.83 ) | 02/20-03/08 | 0.55 ( 0.09 - 1.05 ) | 03/09-04/01 |

Figure S36: Estimated regional reproductive numbers. Calibration from January 4th (2021) up to April 1st. Median and 95 % CIs.

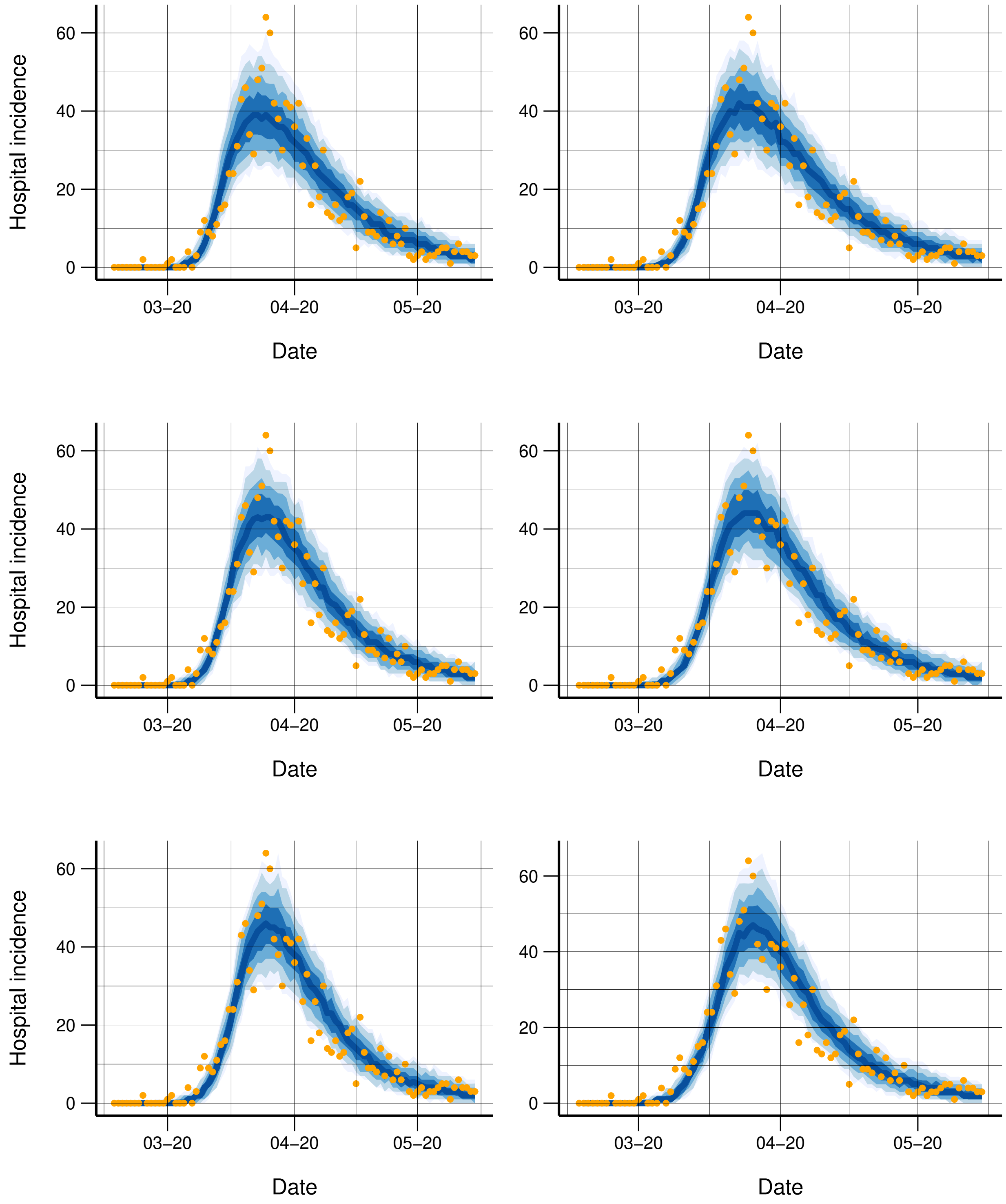


Figure S37: Observed (orange dots) and simulated hospitalisation incidence when varying the changepoint as March 11 (upper left), March 12 (upper right), March 13 (mid left), March 14 (mid right), March 15 (bottom left), and March 16 (bottom right).

### S4 Supplementary discussion

Our predictions and estimates rely on several parameters which are uncertain. There are uncertainties related to the time from symptom onset to hospitalisation, the age-specific hospitalisation risks, and the natural history parameters of COVID-19. We have based our estimates on local Norwegian data where available, and otherwise relied on international studies. To obtain information about the true risk of hospitalisation in Norway, it will be essential to get data on seroprevalence in the population. This will significantly improve the accuracy of the model results. Our model is intrinsically dynamic, in the sense that we update the parameters as new data points arrive daily, and when more information becomes available. This also means that the results have changed over the last months, sometimes significantly. In fact, our estimates of the hospitalisation parameters change regularly, as we have access to anonymous, individual-level data on all hospitalised patients, and we update the parameters when more patients are added.

In our real-time surveillance reports, we need to correct reporting delay in the most recent days of the case data. This was not necessary in this paper, as we worked with historical data. In operation however, we estimate reporting delays by modelling the progressive correction of data, estimated using a binomial model of the proportion of cases that have been reported the last one, two, three, and four days, corresponding to the maximally observed reporting delay. A different way to estimate reporting delay has been suggested in[32].

The metapopulation model is appropriate for incorporating mobility estimates on an aggregated level, like in our setting. It allows us to build a spatial prediction model, which can provide local estimates of infection level. Intraregion details are not included in the model. This means that the model is for example not the most appropriate to estimate the effect of different isolated interventions, like closing of schools. For this, the more detailed, data-hungry, and computationally demanding agent-based models should be used. However, without individual data, it would be difficult to incorporate the real-time mobility in such a model.

### S5 Implementation

The main model is implemented in the R-package spread

`https://github.com/folkehelseinstituttet/spread`.

### References


[1] World Health Organization. WHO Timeline - COVID-19, 2020. [Online;accessed 25-May-2020].

[2] European Centre for Disease Control and Prevention. Outbreak of novel coronavirus disease 2019 (COVID-19): increased transmission globally–fifth update, 2020. [Online;accessed 27-May-2020].

[3] Folkehelseinstituttet. Covid-19-epidemien: risikovurdering og respons i Norge – andre versjon, 2020. [Online; accessed 27-April-2020].

[4] Folkehelseinstituttet. COVID-19-EPIDEMIEN: Risikovurdering og respons i Norge Versjon 3, 2020. [Online; accessed 27-April-2020].

[5] Helsedirektoratet har vedtatt omfattende tiltak for å hindre spredning av Covid-19 - Helsedirektoratet. Library Catalog: www.helsedirektoratet.no.

[6] Government.no. Stricter border controls being introduced – Norwegian airports not closing, 2020. [Online; accessed 18-June-2020].

[7] Government.no. Innfører hytteforbud, 2020. [Online; accessed 18-June-2020].

[8] Government.no. Hold dere hjemme, ha minst mulig sosial kontakt, 2020. [Online; accessed 11-May-2021].

[9] Government.no. Innfører flere nasjonale smitteverntiltak, 2021. [Online; accessed 11-May-2021].

[10] Government.no. Regjeringen innfører strengere nasjonale tiltak, 2021. [Online; accessed 11-May-2021].

[11] Bentzrød, Sveinung B and Stolt-Nielsen, Harald and Ask, Alf O. Innfører flere nasjonale smitteverntiltak, 2021. [Online; accessed 11-May-2021].

[12] Oslo kommune. 9. november: Byrådet har vedtatt sosial nedstenging av Oslo, 2020.

[13] Ann-Kristin Loodtz and Endre Hovland. Byrådet presenterte nye smitteverntiltak, 2020.

[14] Nasjonal kommunikasjonsmyndighet. Ekomstatistikken. [Online; accessed 18-June-2020].

[15] Solveig Engebretsen, Kenth Engø-Monsen, Mohammad Abdul Aleem, Emily Suzanne Gurley, Arnoldo Frigessi, and Birgitte Freiesleben de Blasio. Time-aggregated mobile phone mobility data are sufficient for modelling influenza spread: the case of bangladesh. *Journal of the Royal Society Interface*, 17(167), 2020.

[16] Statistics Norway. 12871: Befolkning, etter kommunestørrelse, alder og kjønn 2017 - 2020. 1-Jan-2020.

[17] M. J. Keeling, L. Danon, M. C. Vernon, and T. A. House. Individual identity and movement networks for disease metapopulations. *Proceedings of the National Academy of Sciences of the United States of America*, 107(19):8866–8870, 2010.

[18] Luca Ferretti, Chris Wymant, Michelle Kendall, Lele Zhao, Anel Nurtay, David G Bonsall, and Christophe Fraser. Quantifying dynamics of sars-cov-2 transmission suggests that epidemic control and avoidance is feasible through instantaneous digital contact tracing. *medRxiv*, 2020.

[19] Yang Liu, Sebastian Funk, and Stefan Flasche. The contribution of pre-symptomatic transmission to the covid-19 outbreak. *CMMID Repository*, 2020.

[20] European Centre for Disease Prevention and Control. Coronavirus disease 2019 (COVID-19) in the EU/EEA and the UK–eighth update, 2020. [Online; accessed 11-May-2020].

[21] Daniel P Oran and Eric J Topol. Prevalence of asymptomatic sars-cov-2 infection: A narrative review. *Annals of Internal Medicine*, 2020.

[22] Odo Diekmann, Johan Andre Peter Heesterbeek, and Johan AJ Metz. On the definition and the computation of the basic reproduction ratio r 0 in models for infectious diseases in heterogeneous populations. *Journal of mathematical biology*, 28(4):365–382, 1990.

[23] Henrik Salje, Cécile Tran Kiem, Noémie Lefrancq, Noémie Courtejoie, Paolo Bosetti, Juliette Paireau, Alessio Andronico, Nathanaël Hozé, Jehanne Richet, Claire-Lise Dubost, Yann Le Strat, Justin Lessler, Daniel Levy-Bruhl, Arnaud Fontanet, Lulla Opatowski, Pierre-Yves Boelle, and Simon Cauchemez. Estimating the burden of SARS-CoV-2 in France. *Science*, (eabc3517), 2020.

[24] Helse Bergen–Haukeland Universitetssjukehus. The Norwegian Pandemic Registry, 2020. [Online; accessed 12-May-2021].

[25] Robert Whittaker, Anja Bråthen Kristofferson, Elina Seppälä, Beatriz Valcarcel Salamanca, Lamprini Veneti, Margrethe Larsdatter Storm, Håkon Bøås, Nina Aasand, Umaer Naseer, Karoline Bragstad, Olav Hungnes, Reidar Kvåle, Karan Golestani, Siri Feruglio, Line Vold, Karin Nygård, and Eirik Alnes Buanes. Trajectories of hospitalisation for patients infected with sars-cov-2 variant b. 1.1. 7 in norway, december 2020–april 2021. *medRxiv*, 2021.

[26] Jonas Christoffer Lindstrøm, Solveig Engebretsen, Anja Bråthen Kristoffersen, Gunnar Øivind Isaksson Rø, Alfonso Diz-Lois Palomares, Kenth Engø-Monsen, Elisabeth Henie Madslien, Frode Forland, Karin Maria Nygård, Frode Hagen, Gunnar Gantzel, Ottar Wiklund, Arnoldo Frigessi, and Birgitte Freiesleben de Blasio. Increased transmissibility of the b. 1.1. 7 sars-cov-2 variant: Evidence from contact tracing data in oslo, january to february 2021. *medRxiv*, 2021.

[27] Folkehelseinstituttet. COVID-19-EPIDEMIEN: Nye varianter av SARS-CoV-2: kunnskap, risiko og respons, 2020. [Online; accessed 08-March-2021].

[28] Peter Bager, Jan Wohlfahrt, Jannik Fonager, Morten Rasmussen, Mads Albertsen, Thomas Yssing Michaelsen, Camilla Holten Møller, Steen Ethelberg, Rebecca Legarth, Mia Sarah Fischer Button, Sophie Madeleine Gubbels, Marianne Voldstedlund, Kåre Mølbak, Robert Leo Skov, Anders Fomsgaard, Tyra Grove Krause, and The Danish Covid-19 Genome Consortium. Risk of hospitalisation associated with infection with sars-cov-2 lineage b. 1.1. 7 in denmark: an observational cohort study. *The Lancet Infectious Diseases*, 2021.

[29] Robert Challen, Ellen Brooks-Pollock, Jonathan M Read, Louise Dyson, Krasimira Tsaneva-Atanasova, and Leon Danon. Risk of mortality in patients infected with sars-cov-2 variant of concern 202012/1: matched cohort study. *bmj*, 372, 2021.

[30] Stephen A Lauer, Kyra H Grantz, Qifang Bi, Forrest K Jones, Qulu Zheng, Hannah R Meredith, Andrew S Azman, Nicholas G Reich, and Justin Lessler. The Incubation Period of Coronavirus Disease 2019 (COVID-19) From Publicly Reported Confirmed Cases: Estimation and Application. *Annals of internal medicine*, 172(9):577–582, 2020.

[31] Scott A Sisson, Yanan Fan, and Mark M Tanaka. Sequential Monte Carlo without likelihoods. *Proceedings of the National Academy of Sciences of the United States of America*, 104(6):1760–1765, 2007.

[32] Radka Jersakova, James Lomax, James Hetherington, Brieuc Lehmann, George Nicholson, Mark Briers, and Chris Holmes. Bayesian imputation of covid-19 positive test counts for nowcasting under reporting lag. *arXiv preprint arXiv:2103.12661*, 2021.

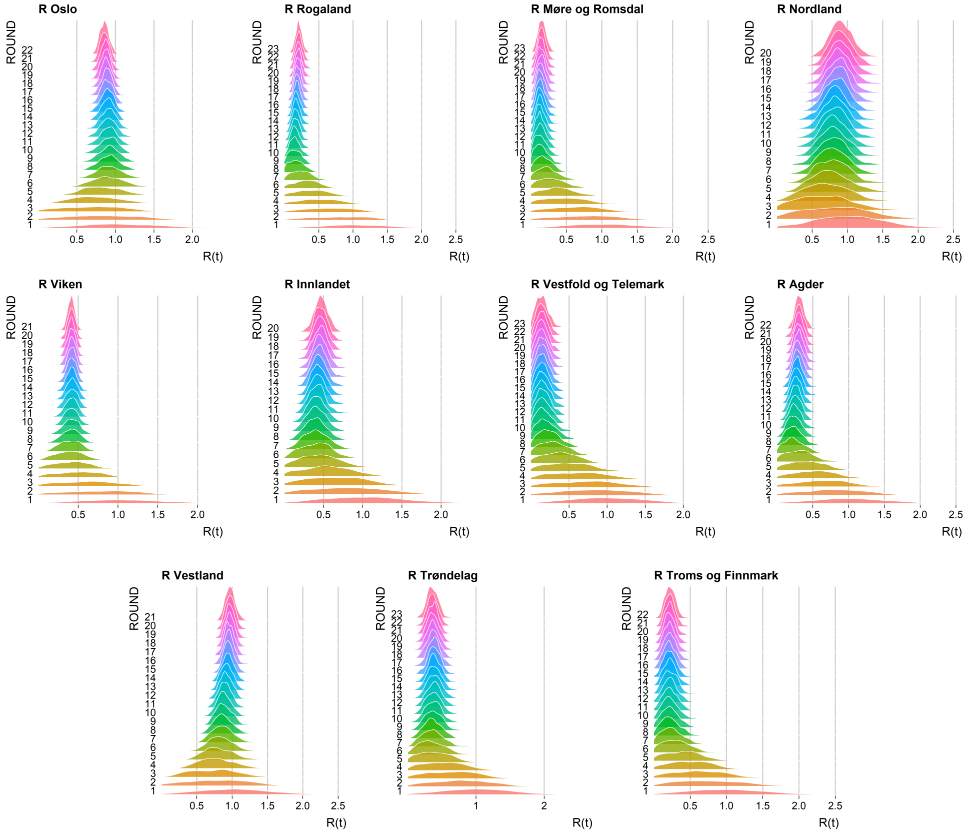


Figure S38: Example of ABC convergence sequences of the distributions of the estimated reproductive numbers throughout the calibration rounds (with data up until September 2020).
